# Information Leakage and Performance Overestimation in EEG-Based Schizophrenia Detection: Evidence from Literature and Empirical Analyses

**DOI:** 10.64898/2025.12.04.25341680

**Authors:** Frigyes Samuel Racz, Gabor Csukly

**Author notes:** Shared correspondence: Frigyes Samuel Racz, Gabor Csukly.

## Abstract

Detecting schizophrenia (SZ) from electroencephalography (EEG) signals using machine- and deep learning models gained traction lately due to potential utility in early disease detection and differential diagnosis. Classification performance reports in the range of 95% accuracy and above are common; however, review of state-of-the-art literature indicates that ∼65% of published works involve erroneous practices in the evaluation pipeline such as epoch- instead of subject-based data splitting or ranking and selecting features before data partitioning. The consequent information leakage can result in an overestimation of SZ detection performance. Here we explicitly test this on three open SZ-EEG datasets using gold standard classification approaches in leaky and leakage-free implementations. Results indicate that information leakage can inflate SZ classification accuracy by up to ∼30%. Accordingly, best practices regarding EEG-based SZ detection must be established and promoted before this technology can be further developed into a clinical decision-making tool.

## Introduction

Schizophrenia (SZ) is a severe, disabling mental disorder affecting about 1% of the population [1]; however, its exact etiology and pathological mechanisms are yet unknown [2]. Since no clinically valid biomarkers are established for SZ, the disorder is currently diagnosed using extensive psychiatric interviews (DSM-5), which can be susceptible to variability arising from heterogeneity in clinical presentation and physician subjectivity. Consequently, there is immense research effort for translating SZ diagnosis, monitoring and management from perception-based practices to those grounded in biology and pathophysiology [3]. Techniques to image or monitor neural activity such as functional magnetic resonance imaging (fMRI) or electroencephalography (EEG) appear as intuitive choices for potential biomarkers in psychiatry; however, as Etkin and Mathalon [4] state in their recent review, the “*low reliability and difficulty standardizing collection are the principal barriers to fMRI, along with the need to demonstrate that its superior spatial resolution over EEG and ability to image subcortical regions directly provide unique clinical value*”. On the other hand, resting-state EEG markers, such as individual alpha peak frequency and power, or microstates exhibit excellent test-retest reliability [5]. Furthermore, with the emergence and increased accessibility of machine learning (ML) and especially its specialized subset deep learning (DL), automated detection of SZ and its characteristic patterns from EEG garnered particular interest lately [6, 7]. Many of these works report stellar SZ detection accuracies ranging 95%, with some even surpassing 99% [8–13]. This performance is well over current estimates of SZ diagnostic accuracy via standard clinical procedures [14], and thus could imply that computer-aided diagnostics (CAD) for SZ are within reach in the near or foreseeable future, as suggested by some reports [15, 16]. In this present work we argue that such cautious optimism is yet uncalled for, as most reported performance metrics are likely massively overestimated due to *information leakage*.

Leakage refers to the scenario when information from outside the training set (and in particular, from the prospective test set) is involved in the model training procedure [17, 18]. This generally results in performance overestimation and poor subsequent generalization. The latter, however, remains unconfirmed if the model is never tested on previously unseen data, which is most often the case for EEG-based classification studies working with relatively small datasets or confined data repositories. While the employed ML/DL methodologies among related works shows great variability, approaches can be sorted into two main categories: *i*) DL architectures utilizing native EEG or that transformed into ‘image-like’ representations (such as spectrograms or scalograms) as input, which predominantly learn group-characteristic patterns from abundant training data and *ii*) DL/ML frameworks utilizing ‘handcrafted’ features (such as spectral, non-linear dynamic or connectivity metrics) that attempt to capture relevant patterns of EEG activity based on a priori hypotheses [6]. Both approaches, but especially approach *i*) relies on vast amounts of data, which is commonly achieved by segmenting continuous EEG into smaller epochs, substantially augmenting the available sample size. On the other hand, feature extraction in *ii*) often results in hundreds of feature candidates and necessitates selection to reduce dimensionality and eliminate noise (i.e., irrelevant features). Consequently, two forms of leakage is especially common when working with EEG, stemming from strong inter-individual variability and high dimensionality: leakage related to *data partitioning* [19] and *feature selection* [20] (DP and FS, respectively, see **Figure 1**). The first case manifests when the DP is not subject-based (i.e., where different samples from the same subject appear in both training and evaluation sets) as the model can pick up on subject-specific patterns (e.g., biometrics), while the second when the complete dataset is used to select most-discriminative features before partitioning. Despite being the leading source of error in ML and DL applications [21], leakage is only gaining traction lately in the neuroscience literature, as demonstrated in neuroimaging-based detection of post-traumatic stress disorder (PTSD) [22], Parkinson’s Disease [23] or Alzheimer’s Disease [24], or in connectome-based ML applications [25]. It was indicated that DP- and FS-related leakage has the most impact on classification performance when working with neuroimaging data [25]. Furthermore, Brookshire and coworkers [26] also reported that over 70% of translational EEG-based DL studies contained some form of leakage, likely inflating performance reports in detecting various clinical conditions. Recognizing these widely present issues prompted a recent urge for establishing best practices in similar approaches aiming for CAD using EEG [19].

**Figure 1.**
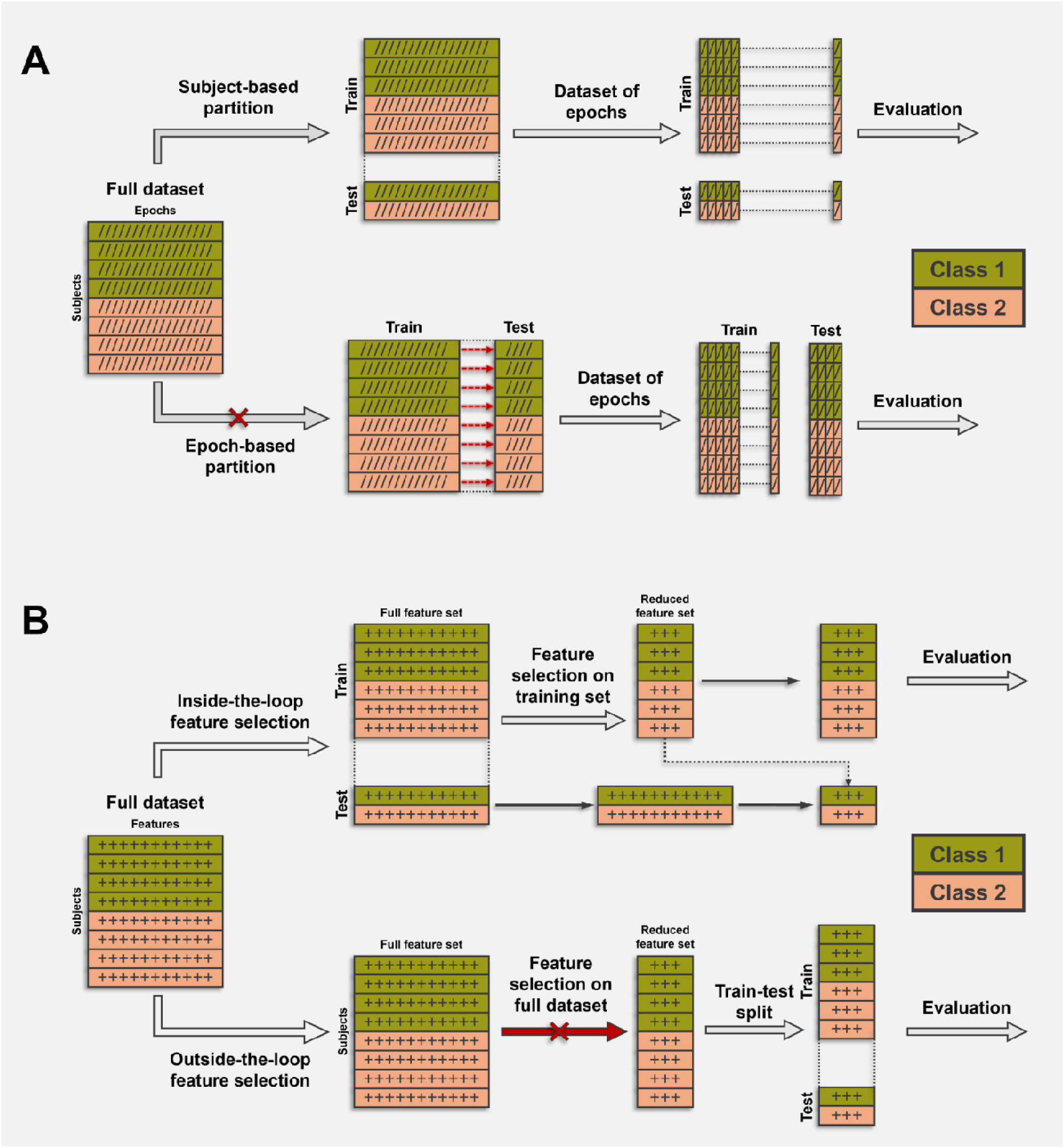
Illustration of various sources of information leakage. Panel **A** contrasts subject-based (good practice/no leakage) vs. epoch-based (bad practice/leakage) data partitioning (DP). In the first case, while each subject contributes multi-epoch data (as indicated by dash symbols), the dataset is first partitioned into train and test subjects, before epochs are considered as individual samples by the model, thus ensuring that all epochs of eac subject are exclusively present in either the train or the test sets. In contrast, in the epoch-based partitioning scheme, the multi-epoch data of each participant is sorted first into train and test samples, which are then used for model training and evaluation. Red arrows indicate that subject-specific patterns will be available in both the train and test sets, causing potential information leakage. Panel **B** illustrates inside-the-loop (good practice/no leakage) an outside-the-loop (bad practice/leakage) feature selection (FS) processes. In the former, while each subject contributes a feature vector with multiple features (as indicated by plus symbols), the dataset is first partitioned into train and test sets. Then, feature selection is only performed using the training set but not considering the test set, and the identified feature subset is extracted from the test samples before training and evaluation. In contrast, in the outside-the-loop setting, the complete dataset is used first to establish a ranking of features based on their discriminating power (leading to information leakage indicated by the boldened red arrow), and then DP, training and evaluation are performed on this previously identified, fixed feature subset. Note that these principles equivalently apply to singular train-test splits and those employed in k-fold cross-validation, and the two scenarios are not exclusive i.e., an evaluation pipeline can contain information leakage related to both DP and FS.

Addressing the potential effect of leakage in EEG-based CAD approaches for SZ is not only relevant but pressing as well. Latest systematic reviews on the topic, [6, 7, 27, 28] indicate that the number of studies using ML or DL techniques to detect SZ from EEG grows exponentially. While over 45 articles report classification accuracies of 95% or higher for SZ (against healthy controls), none of these contemporary review articles address or consider potential leakage effects, nor has this issue been systematically investigated in the case of SZ detection. Overall, developing CAD systems for SZ is indeed important as these efforts can help identifying characteristic biomarkers of the disease, support early diagnosis and enhance differential diagnosis, all being current therapeutic challenges regarding SZ [29]. However, a potential abundance of inadequate ML/DL practices coupled with corresponding inflated performance metrics poses a real risk of misdirecting future research efforts and resources towards unfeasible or unviable solutions [18]. Therefore, addressing the issue of information leakage in state-of-the-art literature on EEG-based CADs for SZ is of utmost importance. To this end, we first screened current literature to assess the presence of DP- and FS-related leakage in previously published studies, then performed empirical testing of ‘leaky’ vs. ‘leakage-free’ classification pipelines on three open EEG datasets. We carried out these evaluations using a variety of approaches including conventional ML, DL, and state-of-the-art, transformer-based architectures. Our findings explicitly demonstrate that bias introduced by various leakage scenarios can inflate SZ classification performance by up to >30%. To the best of our knowledge, this work presents the first synthesis of leakage in relevant recent literature, as well as the systematic assessment of leakage effects on EEG-based SZ detection evaluated via multiple approaches and on multiple real-world datasets. Furthermore, at the end of this article we propose best practices based on conclusions we derive from previous literature and empirical analyses, including a checklist provided as downloadable content to facilitate transparency and consistency among future studies.

## Results

### Leakage trends in current schizophrenia literature

First, we synthesized all articles collected by recent systematic reviews and broad surveys from 2023-2025 on EEG- and ML/DL-based SZ detection [6, 7, 27, 28, 30] to gain initial insight on the presence of leakage in current literature (see **Supplementary Material** for details). As a result, a total of 95 research articles were collected and screened for partition- and FS-based leakage (**Supplementary Table S1**). Leakage related to DP and/or FS was identified in 41 and 34 of these, respectively (with 14 containing both), while potential leakage could not be reliably confirmed in a total of 13 and 10 cases given the provided description (i.e., unclear or unspecified in methods). Out of the total number of 119 evaluations, 25 and 74 contained no or with at least one form of leakage, respectively, not considering 20 cases with incomplete description. Collecting published classification accuracies, the median highest reported performance of 97.00% (/QR: [91.30, 98.99]) from leaky- in contrast to that of 86.00% (/QR: [75.77, 92.2]) from leakage-free pipelines supported our concern that performance metrics are likely overestimated due to leakage effects (p < 4.6098 X 10^-7^, z = 5.0419, Wilcoxon rank sum test, r = 0.5067). Notably, there were 6 papers (reporting 7 pipelines) that explicitly contrasted SZ detection performance in subject-based and epoch-based DP schemes (disregarding FS) on the same dataset [13, 15, 31–34]; synthesizing outcomes showed a statistically significant drop in performance (p = 9.5559 X 10^-4^, t_6_ = 6.0107, paired t-test) with an effect size of d = 2.9515 (C/: [1.4025, 4.4505]) when switching from epoch-based (99.04 +0.78%) to subject-based (88.06 +4.86%) evaluation (**Supplementary Table S2**). Finally, among the 95 screened studies, 28 analyzed private dataset, while 82 did so with one or more of three open EEG datasets ([35–37]).

### Leaky vs leakage-free classification analyses

Accordingly, the observed trends warrant experimental validation. To this end, we implemented leaky and leakage-free realizations for various gold standard ML/DL pipelines employing both data-driven ‘black box’ and handcrafted EEG feature-based approaches. Performance of these models was evaluated on three openly available databases [35, 36, 38] (denoted MSU, RepOD and SU-SZ) containing resting-state EEG recordings from SZ patients and matched healthy control (HC) individuals (see **Table 1** for dataset characteristics). Please see the Supplementary Material (ML-NICAD checklist) for details on hardware/software configurations used throughout this study.

**Table 1.** Basic demographic and EEG data collection characteristics of the analyzed datasets. For age, brackets and square brackets indicate standard deviation and range, respectively. For number of channels, values in brackets indicate the final number of channels included in the analysis. For sampling frequency, values in brackets indicate final sampling rate after downsampling. N/A indicates information not available. HC: healthy control; SZ: schizophrenia; N: sample size; #ch: number of channels; *sf*: sampling frequency; *L*: maximum available data length in all subjects.

| Dataset |  | N | Mean Age | Sex | EEG (#ch) | sf (Hz) | L (s) |
| --- | --- | --- | --- | --- | --- | --- | --- |
| MSU [35] | HC | 39 | 12.25 [11-13.75] | N/A | 16 | 128 | 60 |
|  | SZ | 45 | 12.25 [10.67-14] | N/A |  |  |  |
| RepOD [36] | HC | 14 | 27.75 (3.16) | 7F/7M | 19 | 250 | 720 |
|  | SZ | 14 | 28.10 (3.72) | 7F/7M |  |  |  |
| SU-SZ [38] | HC | 39 | 32.8 (9.5) | 15F/24M | 64 (19) | 1000<br>(250) | 100 |
|  | SZ | 38 | 34.3 (10.7) | 17F/21M |  |  |  |

### The effect of partition-based leakage on classification performance in data-driven pipeline using deep convolutional neural networks

In the first pipeline we utilized a pre-trained ResNet-18 [39] convolutional neural network (CNN) architecture fine-tuned via transfer learning [40]. Continuous EEG segments were spliced into 5-second epochs to augment sample size and epochs were transformed into channel-wise time-frequency features organized into a tri-color image-like structures reflecting scalp topology (see **Methods** and **Figure 4**). HC vs. SZ classification performance was evaluated in a stratified 10-fold cross-validation (CV) pipeline. Random DP was epoch-based (i.e., irrespective of subject ID) in the leaky implementation, while subject-based in the leakage-free scenario. SZ classification performances on the three datasets are summarized in **Table 2**. In the RepOD and SU-SZ databases accuracy dropped significantly from leaky to leakage-free scenarios (confirmed via permutation testing), while the performance drop was marginally significant (*p*=0.0990) in the MSU dataset. The largest leaky vs. leakage free difference was observed in the RepOD dataset (28.27%), followed by the SU-SZ (12.79%) and MSU (4.03%) datasets.

**Figure 2.**
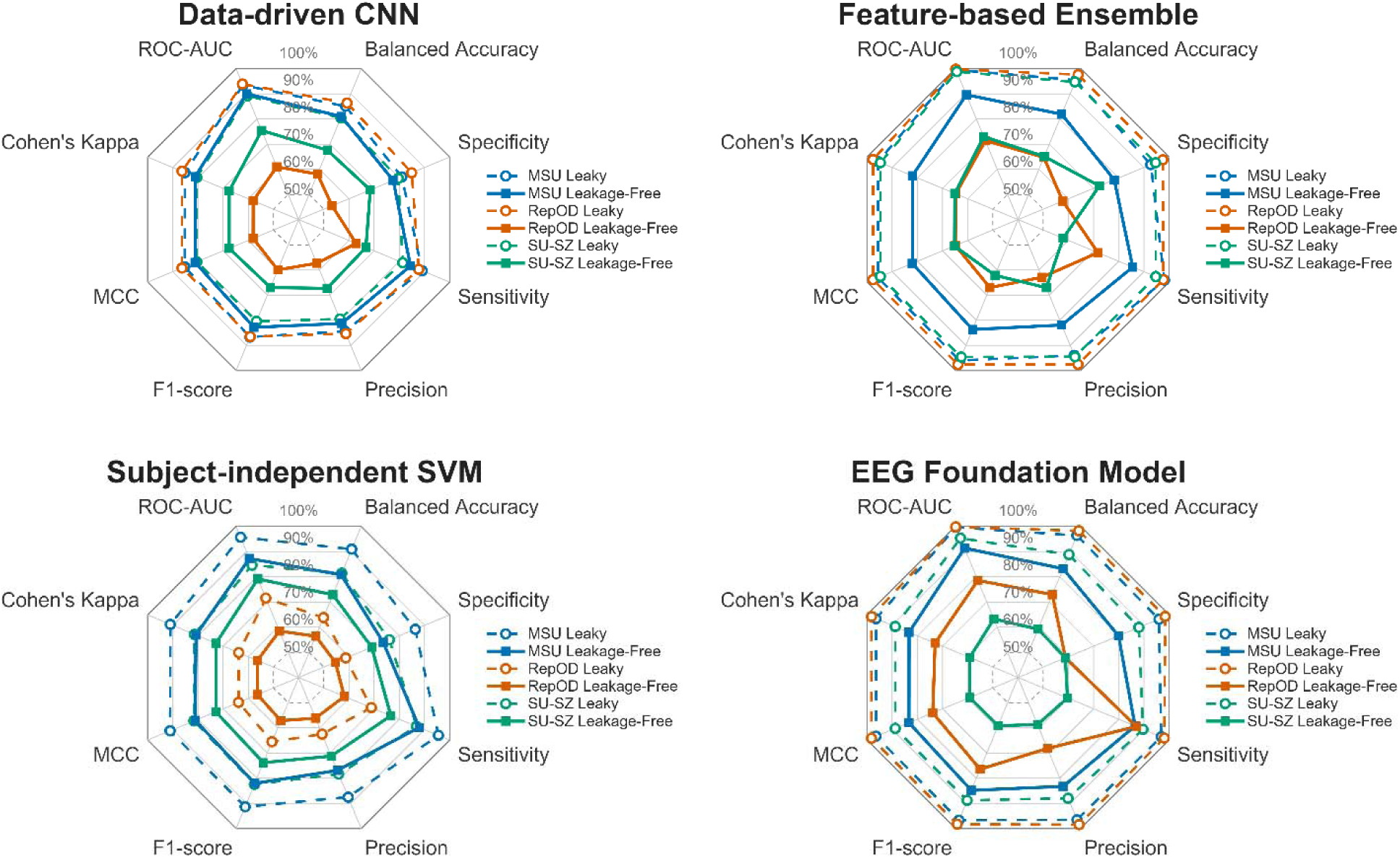
Performance comparison between the four considered classification approaches. Radar plots illustrate model performance on the three datasets in leaky (dashed lines) and leakage-free (solid lines) implementations through eight, standard performance metrics (for definitions, see **Methods**). In all four panels it can be observed that leaky model implementations outperform their leakage-free counterparts in every single performance metric, often by a substantial margin (i.e., >10%), indicating that all four classification approaches are susceptible to leakage-related performance overestimation. CNN: convolutional neural network; SVM: support vector machine.

**Figure 3.**
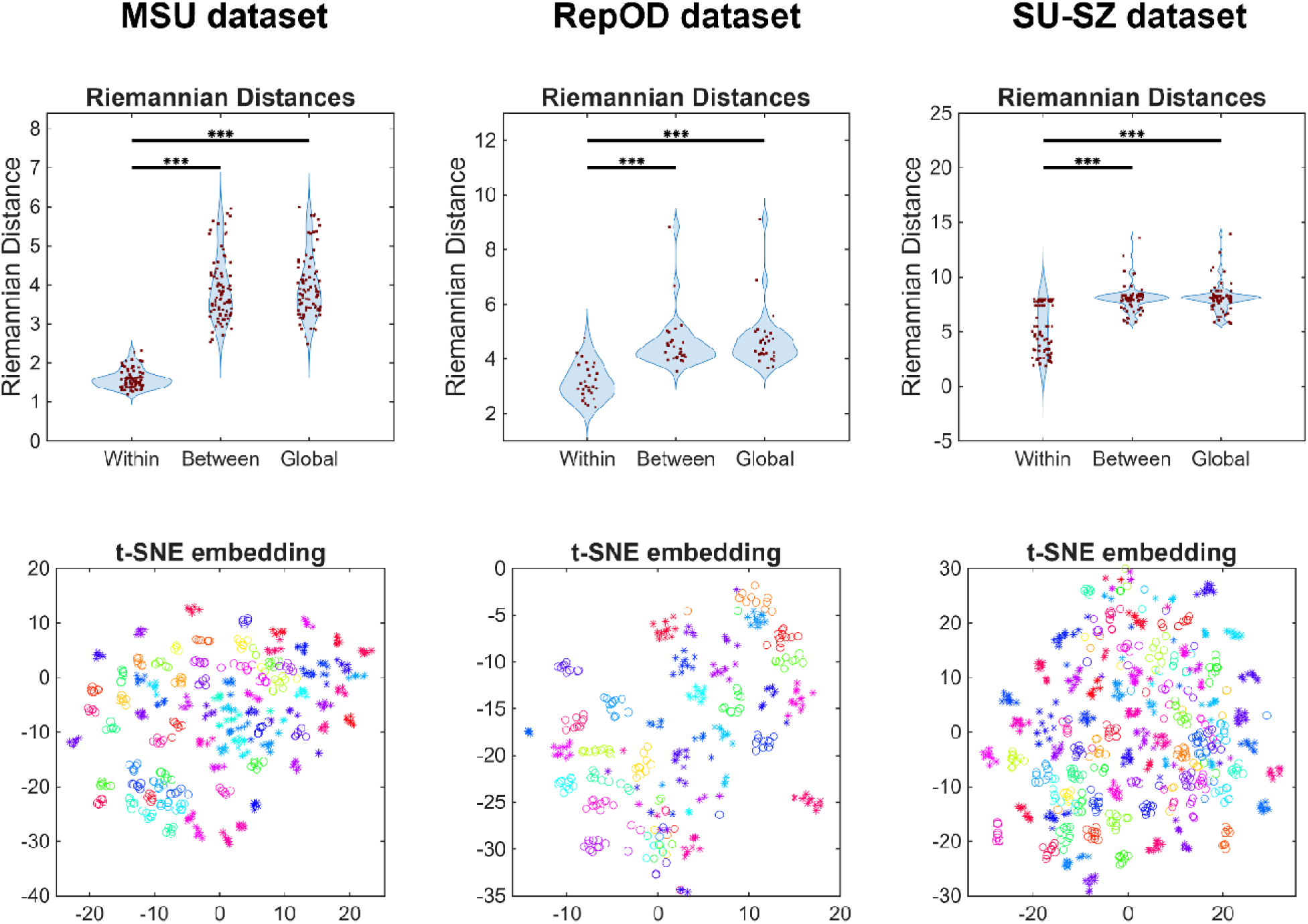
Within- versus between-subject variability of EEG properties. In the upper panels, violin plots show the average Riemannian distance between EEG samples of a given subject and *i*) its own prototype (labeled “Within”), *ii*) the class prototype of the given subject (labeled “Between”) and *iii*) the global prototype computed from all subjects regardless of class label (denoted “Global”). Prototype refers to the Riemannian mean obtained from corresponding covariance matrices (i.e., those from an individual subject, those from all subjects belonging to a given class, and those from all subjects in the complete dataset). Red dots show individual sample values while blu areas illustrate their distribution. Horizontal bars denote significant differences, with *** indicating. Lower panels illustrate a two-dimensional embeddings of multi-dimensional feature vectors using t-SNE. Different subjects are indicated by different colors, while class label is indicated by the symbol ( denoting HC and denoting SZ). It is apparent that samples from the same subject form tight, non- or minimally overlapping clusters, indicating close within-subject proximity while larger between-subject distance of individual samples. Notably, the HC and SZ classes do not separate clearly, suggesting difficult class-based separation.

**Figure 4.**
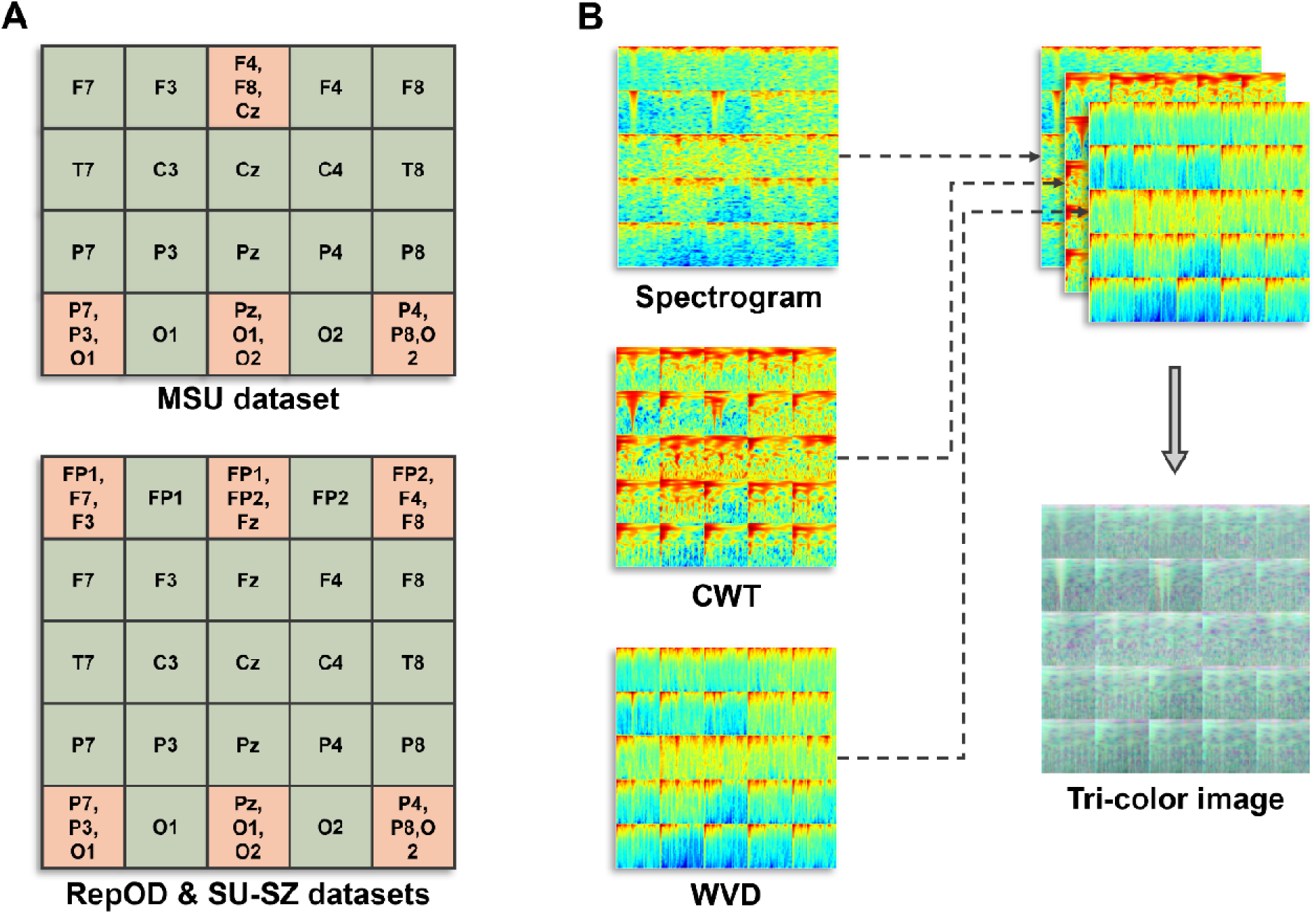
Transforming EEG data into image-like features. Panel **A** illustrates the organizing principle of channel-wise time-frequency maps for the 16- (MSU) and 19 (RepOD and SU-SZ)-channel datasets. Panel **B** depicts the three time-frequency maps obtained from a single epoch, and the resulting tri-color image, as obtained from an exemplary healthy subject from the SU-SZ dataset. It can be seen that the three distinct methods capture varyin aspects of EEG dynamics, providing complementary information, yet their similar nature results in consistent patterns in tri-color image format, ideal for ResNet-18 classification.

**Table 2.** Classification performance reports from leaky and leakage-free implementations of the CNN-based models on the three analyzed datasets. CNN: convolutional neural network; DP: data partition; CI: confidence interval.

| Dataset | Leaky DP | Leakage-free DP | Difference (CI) | p-value |
| --- | --- | --- | --- | --- |
| MSU | 85.32 ± 3.65% | 81.30 ± 8.72% | 4.03% (-0.69; 8.97) | $p=0.0990$ |
| RepOD | 86.33 ± 1.53% | 58.06 ± 7.45% | 28.27% (23.31; 33.57) | **$p=0.0001$** |
| SU-SZ | 80.45 ± 4.76% | 67.66 ± 10.95% | 12.79% (6.51; 19.81) | **$p=0.0009$** |

### The effect of partition-based leakage on classification performance in ensemble-based pipeline using handcrafted features

While the data-driven approach confirmed the effect of partition-based leakage on classification performance, the purpose of the second pipeline was two-fold: *i*) to investigate the extent of leakage introduced by selecting features outside-the-loop and *ii*) to confirm that leakage is also prevalent even when features are manually engineered from EEG data instead statistically learned by the model. A boosted ensemble of decision trees was used as classifier, considered efficient in identifying complex interactions among diverse feature sets [41]. Continuous EEG was first segmented into 10-second epochs (to allow for better estimation of dynamic EEG properties) and five common feature types (frequency domain, time domain, non-linear, complexity and connectivity features, see **Methods**) were computed for each channel, resulting in a total of 386-458 features depending on the number of channels in the dataset. Model performance was evaluated in a 10-fold CV framework similarly as described above with DP-related leakage assessed as described previously. Additionally, we also employed the ReliefF algorithm [42] to select features that efficiently discriminate between HC and SZ. In leaky FS, top 50% features were first selected using the full dataset before 10-fold CV evaluation, while FS was performed within every CV iteration using only the training set in the leakage-free implementation. Since outcomes with these parameters indicated negligible effect of FS leakage in this pipeline, below we report the effect of leaky DP with leakage-free FS (**Table 3**), while more details are reported in the **Supplementary Material** and **Supplementary Table S4**.

**Table 3.** Classification performance reports from leaky and leakage-free implementations of the decision tree-based ensemble models on the three analyzed datasets. DP: data partition, CI: confidence interval.

| Dataset | Leaky DP | Leakage-free DP | Diff (CI) | *p*-value |
| --- | --- | --- | --- | --- |
| MSU | 95.43 ± 4.31% | 82.27 ± 7.91% | 13.16% (6.36; 18.37) | ***p*=0.0001** |
| RepOD | 97.47 ± 1.07% | 64.73 ± 13.58% | 32.74% (24.36; 42.66) | ***p*=0.0002** |
| SU-SZ | 94.42 ± 3.24% | 64.91 ± 8.96% | 29.50% (24.30; 34.94) | ***p*=0.0001** |

Overestimation of classification performance was even more apparent than in the CNN-based pipeline. Namely, classification accuracy neared or surpassed 95% performance dropped significantly in all three datasets (average 25.13%) when switching to leakage-free implementations. Interestingly, while leaky performance was the highest on the RepOD dataset (97.47%), the most substantial drop in classification accuracy was also observed here (32.74%), indicating severe overfitting on subject-specific patterns.

### Realistic subject-based evaluation: the effect of leaky feature-selection on classification performance

While these analyses clearly demonstrate inflation of SZ detection accuracy due to DP-based leakage, they do not yield realistic estimates on clinical performance, as model loss is minimized on epochs rather than aggregated subject-level assignments e.g., via majority vote [43].

Therefore, to mimic a realistic scenario, we re-used the previously obtained hand crafted feature set and characterized each subject with a single feature vector by averaging features over all epochs. Since this scenario eliminates the potential leakage from non-subject-based DP, here we only focused on addressing the effect of leaky FS. Furthermore, given the substantially smaller datasets, we employed a more radical FS pipeline, where among the features selected by ReliefF, only the top 10 were retained using the maximum relevance minimum redundancy (MRMR) technique [44]. Support vector machine (SVM) with radial basis function (RBF) kernel was used for classification. To make maximal use of available data, performance was evaluated in an exhaustive leave-one-subject-out (eLOSO) CV framework and leaky and leakage-free FS was implemented as described previously. As shown in **Table 4**, leaky FS resulted in significant overestimation of performance in all three datasets, with an average 8.72% improvement over leakage-free implementations.

**Table 4.** Classification performance reports from leaky and leakage-free implementations of the SVM-based models on the three analyzed datasets. SVM: support vector machine; FS: feature selection; CI: confidence interval.

| Dataset | Leaky FS | Leakage-free FS | Diff (CI) | *p*-value |
| --- | --- | --- | --- | --- |
| MSU | 90.94 ± 20.06% | 80.77 ± 27.78% | 10.17% (9.12; 11.23) | ***p*=0.0001** |
| RepOD | 63.78 ± 33.04% | 56.38 ± 35.77% | 7.40% (2.30; 12.50) | ***p*=0.0206** |
| SU-SZ | 81.44 ± 27.00% | 72.84 ± 31.01% | 8.60% (7.05; 10.17) | ***p*=0.0001** |

### The cutting-edge approach: are transformer-based EEG foundation models immune to leakage-related performance overestimation?

Beyond conventional ML techniques, recent advances in artificial intelligence led to the emergence of so-called EEG foundation models, which use transformer-based architectures and arguably represent the current frontier of decoding EEG activity [45]. These models are trained using vast amounts (e.g., 10,000+ hours) of diverse EEG data and appear to outperform former approaches on benchmark classification tasks; however, their performance under leaky vs. leakage-free scenarios has not been assessed yet, especially in the case of detecting SZ. For this purpose, we utilized the recently introduced REVE model [46], which provides state-of-the-art performance. Since EEG foundation models are data driven, we devised a similar evaluation scheme as in our CNN-based pipeline probing for DP-based leakage, using 5-second epochs as model input (see **Methods**). Embedding vectors produced by the REVE encoder were mean-aggregated to provide a single feature vector per EEG epoch, and an SVM with a linear kernel was used for classification. As presented in **Table 5**, leakage in DP resulted in significantly overestimated classification performance in case of all three datasets by an average 22.53%.

**Table 5.** Classification performance reports from leaky and leakage-free implementations of EEG foundation model-based approaches on the three analyzed datasets. DP: data partition; CI: confidence interval.

| Dataset | Leaky DP | Leakage-free DP | Difference (CI) | *p*-value |
| --- | --- | --- | --- | --- |
| MSU | 96.43 ± 1.70% | 83.47 ± 9.48% | 12.96% (7.92; 18.31) | ***p*=0.0003** |
| RepOD | 98.26 ± 0.31% | 73.07 ± 20.00% | 25.19% (13.74; 40.57) | **p=0.0003** |
| SU-SZ | 88.80 ± 2.35% | 59.36 ± 13.03% | 29.44% (21.62; 36.47) | **p=0.0001** |

### Performance comparison summary

As a summary, **Figure 2** illustrates model performances in all evaluation schemes through eight different performance metrics commonly employed in similar analyses (see **Methods**). Corresponding numerical values are provided in **Supplementary Table S5** for completeness, while confusion matrices for the four evaluation schemes are illustrated on **Supplementary Figures S2-S5**. Model performances in each scenario are also illustrated via receiver operator characteristic curves on **Supplementary Figure S6**. It can be ubiquitously observed on **Figure 2** that for all four classification techniques that leaky evaluation pipelines (indicated via dashed lines) outperform leakage-free implementations (denoted via straight lines) in all aspects. In most cases, performance in leakage-free cases is balanced albeit poor, indicating low discriminative power. Notably, for the RepOD dataset all leakage-free techniques except the subject-based SVM have stronger sensitivity but low specificity and precision, indicating a bias towards the SZ class and high Type I. error.

While none of the four pipelines stand out as superior in leakage-free scenarios, it is worth noting that performances strongly depend on the dataset at hand. Specifically, while in leaky DP evaluations (i.e., not considering the subject-based SVM pipeline) performances are comparable among the three datasets, a great variability emerges when DP-related leakage is eliminated. The MSU dataset stands out as the most separable with balanced accuracy at or above 80% in all leakage-free evaluations. On the other hand, leaky performances surpassing 90% drop sharply to the 60-70% range (or even below 60%, depending on model architecture) for the RepOD and SU-SZ datasets. This performance range is somewhat lower than most outcomes reported in the literature where the risk of leakage is low (median of 86% accuracy from 25 studies). The most likely reason for this is that we deliberately did not employ hyperparameter tuning (HT), in part to gain insight on between-dataset generalization, but also as HT can be another source of information leakage (see Discussion). Therefore, while leakage-free evaluation performance in the 80-90% range appears achievable for single datasets, ‘realistic’ performance of current models is most likely below that due to poor between-dataset generalizability.

### How partition-based leakage inflates performance: within- vs. between-subject variability in EEG

While the erroneous nature of utilizing the complete dataset for selecting most discriminative features is straightforward, the issue is less obvious with regards to partitioning. Specifically, epoch-based partitioning of data can cause leakage if unique, distinguishing patterns are present in the data that are characteristic of the individual instead of their group. On the level of data this can manifest as smaller within-subject compared to between-subject variability. We performed auxiliary analyses to confirm and illustrate this both for native EEG signals as well as extracted signal features (see **Methods**). Riemannian geometry-based analysis of covariance-transformed EEG epochs confirmed that within-subject spread of samples was significantly smaller (*p*<10^-5^ in all cases) than between-subject (group-level or global) variability in the data for all three datasets (**Figure 3** upper panels, see also **Methods**). Additionally, 2-dimensional projection of multi-dimensional feature vectors via t-distributed stochastic neighborhood embedding (t-SNE) [47] indicated that same-subject samples form tight, non-overlapping clusters, while HC and SZ groups cannot be clearly delineated (**Figure 3** lower panels). Both analyses support that the dependence of EEG samples obtained from the same subject is stronger than group association, likely due to subject-specific characteristics. Therefore, there is real risk that an ML/DL model will learn to recognize these patterns and use them for decision making – instead of less potent group-characteristic patterns – if this dependency is not considered during DP.

## Discussion

The two cases of information leakage considered in this study – namely, DP- and FS-related leakage – are both categorized as explicit cases of “test-to-train” leakage in the recent survey of Sasse and colleagues [18]. Specifically, ranking and selecting features using the complete dataset (i.e., outside-the-loop) directly exploits information from prospective test samples in the model training, leading to falsely inflated performance. This effect was addressed formerly by Shim and coworkers [20] on simulated data and EEG from HC and PTSD cohorts, showing that outside-the-loop FS overestimated classification accuracy on average by ∼9% compared to inside-the-loop FS. This aligns well with our findings (8.72 +1.39%) in the case of SZ detection (**Table 4**). Our analyses indicate that effects stemming from DP can be even more severe (15.03 + 12.27% and 25.20 +10.38%). This is somewhat smaller than those reported in previous SZ literature [13, 15, 31–34] (10.98 +4.83% on average), although none of these studies attributed this effect to information leakage. On the other hand, Del Pup and colleagues [19] recently compared epoch- vs. subject-based pipelines to classify Parkinson’s and Alzheimer’s patients against HC and reported substantial drops in balanced accuracy (ranging ∼15-45%). A fundamental theoretical assumption of supervised learning approaches is that different examples in the dataset are independently and identically distributed (I.I.D.) and sampled from a fixed probability distribution [18]. The I.I.D. assumption is violated by multiple EEG samples (epochs) from the same individual, as they likely contain the same characteristic traits specific to the individual. This was supported by our auxiliary analyses, even when attempting to match data distributions among subjects (**Figure 3**). As a result, it is possible that an improperly trained model will recognize the *individual* (i.e., Jane Doe/John Smith) as their characteristic patterns were encountered during the training phase and simply predict the already associated group label instead of making this decision based on group-specific (i.e., HC/SZ) traits. Another important consequence of this phenomenon is that a model relying on subject-characteristic traits will likely categorize new, previously unseen subjects poorly, whose data were not encountered during the training procedure. In that regard, none of the reviewed articles reported cross-dataset classification performances. Overall, our findings emphasize an issue verifiably present in current literature, and while we can only conjecture, it is likely that performance metrics in the 42 other sampled studies employing only epoch-based DP are overestimated to a similar extent.

There is yet another aspect beyond DP and FS that we did not consider here, namely tuning hyperparameters of the classifiers. Training most DL models involve the heuristic selection of numerous hyperparameters (e.g., learning rate, optimization algorithm, hidden layer architecture, or regularization [48]) and optimal hyperparameter settings most often yield substantially better performances [49]. While HT can be performed both outside- and inside-the-loop similarly to FS, here we decided not to consider nor employ it for reasons outlined below. As emphasized by Del Pup and colleagues recently [19], training DL models for EEG classification should utilize a nested CV pipeline, where in each CV iteration (outer loop), an internal CV (inner loop) is performed by further partitioning the training set for HT. To facilitate an exact comparison between all our cases, we omitted this step in our CNN-based pipeline and instead model hyperparameters were either set to defaults or according to common ML/DL practices. This is likely the reason why our models – even with leaky implementations – underperformed compared to similar approaches in the literature (e.g., [50]), where HT was employed. It should be emphasized, however, that i) hyperparameter tuning, if not employed in a nested CV framework, is expected to have a similar effect on performance as leaky FS, and ii) excessive hyperparameter tuning on limited small datasets even with best practices can result in overfitting to dataset characteristics, resulting in poor generalization to new data [51].

Considering the excessive computational burden associated with HT, as well as to maintain clarity and conciseness of this paper, we eventually decided not to explore the effects of HT here (similar to previous works, see e.g., [25]). Nevertheless, we acknowledge this as a current limitation and an important research question that we intend to pursue in future work.

Further considerations should be made when working with openly available datasets. Recent work indicates that optimal parameter settings yielding the highest performance are often specific to the dataset at hand [49]. Therefore, it is probable that performance metrics obtained on these relatively small, open EEG databases (80 evaluations from the literature sample, as well as our analyses) would not hold when final models were tested on newly obtained data. Second, excessive use of open datasets invokes the issue of “dataset decay” [18]: when multiple hypotheses are tested on the same dataset, the chance of Type I error and thus false discoveries increases. As a reference, 43 out of 95 papers in our sample analyzed the RepOD dataset. Furthermore, given available reports on the same dataset, current/future research is incentivized to push for even higher performances; this can potentially result in models that overfit on dataset idiosyncrasies besides disease-specific traits, or employing features based on a priori knowledge of the same given dataset. Consequently, there is increased likelihood that some of the outstanding performances are results of chance or influenced by double dipping, also diminishing generalizability of new findings due to dataset over-usage [18]; however, we must note that this is not explicitly supported by data here. Finally, our analyses show substantial variability in leakage-free same-model performances for the three datasets, likely stemming from differences in the data collection environment and subject cohort characteristics. Importantly, this variability is much smaller when leakage is present, indicating that a single, outstanding performance report obtained from a singular dataset is no guarantee for true, clinically relevant generalizability. These notions all emphasize the need for open research efforts and especially data sharing. In this regard, the SU-SZ dataset, which was published in 2025 and at the time of writing this manuscript have not yet used in an EEG-based SZ detection study [38], might provide a new opportunity to test the generalizability of previously developed classification approaches.

We would also like to briefly address data reporting standards in relevant literature. According to our sampling, 22 out of 95 articles provided insufficient information in their methods description to confirm if e.g., if DP was epoch- or subject-based, or if FS was performed inside or outside the CV loop. This proportion is much higher when one aims for an exact replication of the analytical and evaluation pipeline e.g., the full parameter space scanned during HT is incompletely reported. Access to code implementations is rarely provided (only 3 out of 95 sampled cases), preventing verification of the outcomes.

To propel EEG-based CAD systems in SZ towards true clinical utility, there appears to be a need for establishing best reporting practices considering both dataset, analytical and evaluation pipeline aspects. To this end, below we summarize the most relevant takeaways substantiated by our analyses in **Table 6**. Furthermore, to facilitate consistency among independent studies and transparency in reporting methods and results, we propose a checklist template, which we termed the Checklist for Transparent Reporting of Machine Learning Pipelines in Neuroimaging-based Computer-Aided Diagnostics (**ML-NICAD Checklist**). This checklist is composed of two sections. The first allows for a concise summary of the article where the outcomes are reported, including core details of the analyzed dataset(s). The second concerns common building blocks of classification pipelines, including *i*) model specifics and hardware/software environment, *ii*) data preprocessing, *iii*) feature extraction/engineering, *iv*) model architecture and training, *v*) hyperparameter tuning *vi*) data partitioning and cross-validation, *vii*) feature selection, and *viii*) performance evaluation. The blank document can be accessed as **Supplementary Document 1** of this submission. It is important to acknowledge that not all classification approaches might fit the same rigid framework and thus we intend the ML-NICAD checklist to be customizable; as an illustration, we provide the completed checklists detailing all evaluation pipelines presented in this study in **Supplementary Document 2**.

**Table 6.** Best practices and recommendations for developing EEG-based CAD systems.

| Aspect | Recommendation |
| --- | --- |
| Data Partitioning | In case of data augmentation via EEG segmentation, data should always be partitioned along subjects instead of treating epochs as independent samples. |
| Feature Selection | Feature selection should always be performed excluding test data and using only the training set (inside-the-loop for CV). |
| Hyperparameter tuning | Hyperparameter tuning should always be performed along the same lines as FS. Since HT also requires validation data, employ nested, subject-based CV as recommended by Del Pup and colleagues [19]. |
| Generalization | Verify classification pipelines on multiple, independent datasets to better assess the generalizability of the proposed approach across data collection environments and patient cohorts. |
| Transparency | Provide a detailed, structured summary of the evaluation scheme, with particular emphasis on how DP, FS and HT were handled (e.g., via the ML-NICAD checklist), if applicable. |
| Reproducibility | Whenever possible, share analysis codes for validation and reproducibility. |
| Dataset Decay | Whenever possible, share anonymized EEG data to facilitate development and cross-validation of EEG-based CAD systems for SZ detection. |

Lastly, we address the limitations and broader implications of this present study. Foremost, the central aim for this work was not to propose a benchmark model for EEG-based SZ detection, but to emphasize the potential effects of leaky evaluation protocols. As such, the reported leakage-free performances should not be treated as ‘state-of-the-art’, especially that certain steps (most particularly, HT) were omitted. We only focused on leakage stemming from DP and FS as we believed these being to most pertinent in literature; there are, however, numerous other sources of leakage to attend to, as summarized in [18]. Performances in the CNN-based pipeline could be further improved via nested CV scheme involving HT [19]. Here we only analyzed previously shared, confined EEG datasets; none of the models were evaluated on external samples, and cross-dataset or pooled-dataset performances were not explored. Small sample sizes also limit the generalizability of our findings. We only evaluated four gold standard ML/DL approaches here, while other techniques (e.g., recurrent neural networks [52] or other transformer-based architectures [53] besides EEG foundation models) also show promise. Note, however, that the leakage mechanisms considered here (also including HT) are model agnostic; therefore, they are expected to have similar effects in a broad range of approaches where these notions are applicable. Evaluations only considered resting-state EEG databases for the sake of comparability, although task-specific EEG patterns can provide additional opportunities for SZ detection such as exploiting evoked response differences [37]. We also did not account for demographic differences such as the MSU dataset [35] consisting of adolescents instead of adults, or medication effects such as in the SU-SZ dataset. Our literature screening was based on recent systematic reviews [6, 7, 27, 28], discarding works that were published outside the inclusion period of these papers. Therefore, we likely omit a substantial amount of recent research, given the increasing trend in the number of related publications (see also **Supplementary Figure S1**) and considered time frame. As an exploration, we performed a screening of literature published after January 1^st^, 2024, using the Rayyan [54] tool (see **Supplementary Material**). After removing duplicates, screening of 219 titles and abstracts indicated 125 articles for full-text consideration. While performing an updated systematic or scoping review to synthesize these works is beyond our current scope, these trends clearly indicate the heightened interest in EEG-based SZ detection and warrant frequent and structured synthesis of relevant literature. Finally, we should like to emphasize that while we investigated the issue of leakage in the case of SZ detection, the implications of our findings are not limited to this particular problem, and similar concerns are likely to arise across a wide range of EEG-based psychiatric or neurological applications, as already indicated by the literature [22, 26]. Furthermore, these issues are not unique to EEG as a modality, and can emerge whenever limited sample sizes are countered with segmentation-based data augmentation or excessive feature engineering e.g., in fMRI-based studies [25].

## Data availability

No new physiological data was generated throughout this study as only openly available datasets were analyzed. The MSU dataset [35] can be accessed at the following hyperlink: http://brain.bio.msu.ru/eeg_schizophrenia.htm. The RepOD dataset [36] can be accessed at the following hyperlink: https://doi.org/10.18150/repod.0107441. The SU-SZ dataset [38] can be accessed at the following hyperlink: https://doi.org/10.5281/zenodo.14808295.

## Code availability

All code, including but not limited to scripts and corresponding functions performing data pre-processing, feature extraction, classification and results presentation will be made available without restriction upon publication of this article at the corresponding author’s GitHub repository (https://github.com/samuelracz/schizophrenia_EEG_detection_leakage).

## Author contributions

FSR and GC conceptualized the study. FSR performed all data analyses, literature review and drafted the manuscript. GC contributed to manuscript development and results interpretation.

## Methods

### Literature review

As there are multiple recent systematic reviews on EEG-based detection of SZ [6, 7, 27, 28], we synthesized all the articles considered by these works instead of performing another systematic review. While the work of Khare and colleagues [30] is not a systematic review, it contains a survey of over 40 papers (see Table 1 in [30]) and thus we decided to also include it in the synthesis. After eliminating duplicates and inadmissible works (see **Supplementary Material**, section **Notes** for details), this procedure yielded 95 original publications (**Supplementary Table S1**), including a total of 119 evaluation pipelines (some works analyzed multiple datasets). The full text from all these publications was assessed. We followed the screening guidelines and categorization recently proposed by Young and colleagues [24] to assess if leakage was i) present, ii) absent, or iii) it could not be determined with reasonable certainty. Please note that these categories are denoted as high, low or moderate risk for leakage, respectively, in the original work [24]. Classification accuracy (or in cases of multiple evaluations, the highest reported performance) was extracted from each evaluation pipeline as the characteristic performance metric. For more details on the assessment of leakage risk, please see the **Supplementary Material**.

### Experimental datasets

We analyzed recordings from three, open EEG repositories, detailed below. In all datasets, individuals in the SZ cohorts had a diagnosis of SZ according to international ICD-10 (F-20, F-21 or F-25) established by a psychiatrist. We also report medication information where available.

#### Dataset 1: MSU dataset

The first dataset was collected at the M.V. Lomonosov Moscow State University in Moscow, Russia, and shared by Borisov and colleagues [35, 55], containing one minute of eyes-closed resting-state EEG data from *n*=39 HC (mean: 12.25 years, range: 11-13.75 years) and *n*=45 SZ (mean: 12.25 years, range: 10.67-14 years) adolescents. EEG was collected at 128 Hz sampling from 16 locations (standard 10-20 locations excluding Fp1, Fp2 and Fz) referenced to linked earlobes. The original study was approved by the ethical committee of Moscow Mental Health Scientific Center of Russian Academy of medical Sciences, and written informed consent was collected from parents of the participants [56]. No information was available on the distribution of females/males in the study groups, or if SZ participants were on medication at the time of recording. The dataset is available at http://brain.bio.msu.ru/eeg_schizophrenia.htm.

#### Dataset 2: RepOD dataset

The second dataset was collected at the Institute of Psychiatry and Neurology in Warsaw, Poland, and shared by Olejarczyk and Jernajczyk [36], containing ∼15 minutes of eyes-closed resting-state data from *n*=14 HC (7 females aged 28.7±3.4 years and 7 males aged 26.8±2.9 years) individuals and *n*=14 patients with paranoid SZ (7 females aged 28.3 ± 4.1 years and 7 males aged 27.9 ± 3.3 years). Data was sampled at 250 Hz from 19 standard locations of the international 10-20 montage [57], with the reference electrode positioned over FCz. The Ethics Committee of the Institute of Psychiatry and Neurology in Warsaw reviewed and approved the study protocol and all participants provided written informed consent before recordings. Individuals in the SZ cohort were subjected to a medication washout period with a minimum of seven days before EEG collection. The dataset can be accessed at https://doi.org/10.18150/repod.0107441.

#### Dataset 3: SU-SZ dataset

The third dataset was collected at the Department of Psychiatry and Psychotherapy, Semmelweis University in Budapest, Hungary, and shared by Racz and colleagues [38]. The full dataset contained 2 minutes of eyes-closed resting-state data from *n*=39 HC individuals (age: 32.8±9.5 years, 15 females) and *n*=38 patients with SZ (age: 34.3±10.7 years, 17 females). Original data was collected from 64 scalp locations at 1000 Hz sampling; however, for dataset comparability, only the subset of 19 channels according to the 10-20 montage was considered in this study and the data was downsampled to 250 Hz (see below). The Semmelweis University Regional and Institutional Committee of Science and Research Ethics reviewed and approved the study protocol, and all participants provided written informed consent before enrollment. Participants were ON medication at the time of EEG collection. The dataset is available at https://doi.org/10.5281/zenodo.14808295.

### Ethics statement

No original data from human participants was recorded for this manuscript; all analyzed data was obtained from repositories available in the public domain. All original studies were conducted in line with the Declaration of Helsinki, approved by their respective regional ethics committees (see details above), and written informed consent was obtained from all participants. The current study was approved by the Semmelweis University Regional and Institutional Committee of Science and Research Ethics (registration number: 197/2015) and was conducted in line with the Declaration of Helsinki.

### EEG preprocessing

Pre-processing of raw EEG data was carried out in MATLAB 2025b (The MathWorks, Natick, MA) using the EEGLAB toolbox [58] version 2024.0. The same pre-processing steps were applied in all three datasets, with exceptions noted when applicable. From the MSU dataset, all available 1-minute EEG was considered, while from the RepOD and SU-SZ datasets the first 720 seconds and 100 seconds of EEG recordings were selected, respectively, being the longest available segment for all participants in the corresponding datasets. Raw, continuous EEG data was first band-pass filtered between 1-45 Hz using a finite impulse-response filter using default EEGLAB settings. Additionally, the CleanLine algorithm [59] was used to eliminate line noise at 50 and 100 Hz. The 1000 Hz data of the SU-SZ dataset was downsampled to 250 Hz for computational efficiency and comparability with the other datasets. Then, independent component analysis (ICA) [60] was utilized to remove artifact signal components such as those related to eye movements, muscle or cardiac activity. The multiple artifact rejection algorithm [61] was used to identify artifact-related independent components based on six temporal, spatial and spectral features, which were removed before regaining the signal via reverse-ICA. Finally, data was re-referenced to the common average electrode, and signal for each channel was standardized to have zero mean and unit variance, thus eliminating spatial variability due to electrode impedances.

### Deep learning pipeline using image-transformed EEG features

The advantage of DL classification is that it does not require extensive feature engineering; instead, complex models, such as CNNs learn features from data directly. In particular, CNNs are most commonly used in computer vision applications [62], as early convolutional filters are effective in learning simple spatial patterns (such as edges and primitive shapes), while those in later layers of the neural network synthesize them to encode more complex spatial structures. In general, CNNs are effective on any type of data where information can be encoded as spatial or spatiotemporal patterns, and they are widely used in EEG classification based on time-frequency features [63]. On the downside, since CNNs usually involve training of millions of model parameters, they require substantial amounts of training data. To circumvent this, data augmentation techniques are commonly employed, such as segmenting continuous EEG into shorter epochs and thus multiplying sample size [63]. Additionally, when training data is scarce, transfer learning [40] is often employed, only retraining the final, fully connected layer(s) of a deep CNN already trained on millions of samples, thus preserving the model’s capability to extract and recognize low-level features. Accordingly, in this pipeline we first segmented continuous EEG into short epochs, transformed EEG them into ‘image-like’ time-frequency features encoding spatiotemporal information, and fine-tuned a pre-trained CNN architecture for EEG epoch classification. Our analytical pipeline is a modified version of that proposed by Khare and colleagues [50].

### EEG transformation

**C**ontinuous, pre-processed EEG was segmented into 5-second, non-overlapping epochs. From these, time-frequency information was extracted for every channel in three separate ways, much like in [50]: spectrogram using the Welch periodogram method, scalogram using continuous wavelet transformation (CWT), and smoothed pseudo Wigner-Ville Distribution (WVD). The spectrogram was estimated using the `spectrogram()` Matlab function with a window size of 1-second, overlap of 90% (0.1-second step size), Hanning-windowing and a frequency resolution of 0.25 Hz. Complex-valued spectrograms S were converted into dB according to dB = 20 * log_10_(|S|). CWT transformation was performed using the `cwt()` Matlab function with default Morse wavelet, and WVD was performed using the `wvd()` Matlab function with “smoothedPseudo” option. Both CWT and WVD outputs were log-transformed for better scaling. From all three time-frequency decompositions, data from outside the 1-45 Hz regime were excluded. The obtained channel-wise spectrogram, CWT and WVD matrices were then arranged into a tri-color image the following way. For the MSU dataset, a 4X5 grid was created, while a 5X 5 for the RepOD and SU-SZ datasets, due to the slightly different channel arrangement (16 vs. 19 channels). For all three methods, time-frequency matrices obtained from each channel were assigned to grid positions corresponding to their approximate arrangement on the scalp, as illustrated on **Figure 4A**, with grid cells not corresponding to an existing scalp location filled using average time-frequency matrices of nearby channels. These composite images were then rescaled to 224 X 224 using the `imresize()` Matlab function and scaled between [0, 255] to fit input requirements for the ResNet-18 architecture (see below). Finally, these processed matrices from spectrogram, CWT and WVD were organized into a tri-color image of size 224 X 224 X 3 (**Figure 4B**), capturing short-term spatiotemporal dynamics in EEG. The procedure was carried out for all non-overlapping epochs in all subject, resulting in 12 samples per subject (1008 total) for the MSU dataset, while 144 (4032 total) samples and 20 (1540 total) samples for the RepOD and SU-SZ datasets, respectively.

### Evaluation pipeline

We employed the widely utilized ResNet-18 [39] architecture, pre-trained on over one million images of the ImageNet dataset [64], as implemented in the MATLAB Deep Learning Toolbox `imagePretrainedNetwork` library. Since ResNet-18 has about 11.7 million parameters, we employed a transfer learning framework to prevent overfitting. In that, we froze weights in the first ten convolutional layers of the model, added a DropOut layer with probability 0.4 before the final fully connected layer, and replaced the classification layer to predict two outputs after applying SoftMax on the preceding fully connected layer outputs. The resulting architecture was then evaluated in two pipelines on all three datasets independently, using a k-fold CV framework, where in each of k folds, 1/k proportion of the data was set aside for evaluation and the model is trained on the remainder (1-1/k) of the data. In the leaky implementation, we employed a stratified k-fold CV evaluation in which data epochs were assigned to train and evaluation sets regardless of their subject ID but preserving HC-SZ proportions in both sets. In the leakage-free implementation, during each CV iteration the subjects were first partitioned into training and evaluation subjects, then all samples from a given subject were sorted exclusively into the training or evaluation sets. The MSU and SU-SZ datasets allowed for an approximate 10-fold CV framework in the leakage-free, subject-dependent implementation (e.g., 3-4 HC and SZ in the evaluation set) while preserving the approximate proportion of HC and SZ in each evaluation folds, however for the much smaller RepOD dataset we utilized 7-fold CV instead, yielding completely balanced partitions. In both leaky and leakage-free CV schemes, all samples of the dataset were utilized as an evaluation sample at least once, and exactly once.

### Machine learning pipeline using handcrafted EEG features

Small datasets might prevent the full utilization of DL capabilities in learning features from data without risking overfitting. Instead, one might employ certain feature engineering techniques to extract relevant information from data guided by a priori knowledge [65]. This approach is also commonly utilized in EEG classification studies [66–69], complementing the previously described data-driven approach. In most cases, a wide range of features is obtained initially along different principles, in order to capture various aspects of complex EEG signals.

### Feature extraction

In our approach, we computed five different types of EEG features commonly utilized in the SZ literature (see e.g., [70]): *i*) power spectral features, *ii*) time-frequency features, *iii*) time-domain features, *iv*) non-linear dynamic features and *v*) connectivity features. Pre-processed EEG wa first segmented into non-overlapping epochs of 10 seconds to allow for a better characterization of dynamic properties. Then, various EEG features for each channel were computed as follows (for details and formulas, see **Supplementary Material**):

- *i*)-*ii*) power spectral and time-frequency features: in every epoch, for every channel a spectrogram was obtained using 1-second window size, Hanning-windowing, 75% overlap (0.25-second step size) and frequency resolution of 0.25 Hz. Spectral power was integrated over five canonical frequency bands (delta: 2-4 Hz, theta: 4-7 Hz, alpha: 7-12 Hz, beta: 12-30 Hz and gamma: 30-45 Hz) to obtain time-resolved band-limited power (BLP) estimates. Then, each frequency band was characterized by its mean and variance of BLP over time. This procedure yielded 10 features per channel, per epoch.
- *iii*) time domain features: temporal complexity of broadband (1-45 Hz) EEG in each 10-second epoch was characterized via Hjorth parameters [71] activity, mobility and complexity, and the detrended fluctuation analysis (DFA) scaling exponent [72]. Since Hjorth activity is the variance of native EEG, this feature was computed from pre-processed EEG before standardization, while mobility and complexity were obtained from standardized EEG data. DFA scaling exponents were obtained using the efficient multi-channel method proposed in [73]. These procedures yielded 4 features per channel, per epoch.
- *iv*) non-linear dynamic features: non-linearity was characterized in the five canonical frequency regimes (as above) using Permutation Entropy (PE) [74]. For this analysis, pre-processed continuous EEG data was first band-pass filtered to the corresponding frequency regime using a 4^th^ order zero-phase Butterworth filter. Then, normalized PE was computed using the full 10-second EEG of each epoch, for every channel. Parameters of PE were set to match the frequency regime characteristics as outlined in [75] (**Supplementary Table S3**). This analysis produced 5 features per channel, per epoch.
- *v*) connectivity features: we utilized the Phase-Lag Index (PLI) as our connectivity estimator [76]. In that, weighted adjacency matrices were constructed for each frequency band by computing pairwise PLI between all channel pairs. Following the approach in [67], matrices were density-thresholded [77] using a set of threshold values ranging from 15% to 50% to eliminate spurious connections, and matrices were collapsed over various threshold values to yield robust estimates. Finally, in every frequency band the node degree was estimated for each EEG channel, yielding 5 node degree estimates for each channel, per epoch.

The above-described procedure produced feature vectors F^t^ E lffi.^1x24^ for each channel ch and epoch t. Feature vectors were concatenated according to all channels, leading to training samples of size F^t^ E lffi.^1x3B4^ for the MSU dataset and F^t^ E lffi.^1x456^ for the RepOD and SU-SZ datasets. Every subject in the MSU dataset provided 6 samples (504 total), while 72 samples (2016 total) and 10 samples (770 total) for the RepOD and SU-SZ datasets.

### Feature selection

The feature extraction procedure yielded 384 initial features per epoch in the MSU dataset, while 456 initial features in the RepOD and SU-SZ datasets. To reduce dimensionality and eliminate irrelevant features from our initial set, we performed feature selection using the ReliefF algorithm [42]. ReliefF is a supervised feature selection algorithm that iteratively samples and evaluates feature relevance based on the feature’s ability to differentiate nearest neighbors from the same class (hits) and different classes (misses). ReliefF assigns positive weights to features that consistently show small differences in values among hits but large differences among misses, while negative weights arise when differences are larger among hits than misses. Therefore, a negative weight indicates that a feature is irrelevant for the classification problem. Here we utilized ReliefF in a simple manner with the number of nearest neighbors set uniformly to 10. As we have found that ReliefF assigned positive weights to over 50% of initial features in every case, we decided to uniformly retain the top 50% of features for classification in every evaluation scenario and CV iteration. This resulted in a final feature set of 192 for the MSU, while 228 for the RepOD and SU-SZ datasets. Feature selection was implemented in leaky and leakage-free ways. In the leaky scenario, first all features were standardized across the complete dataset. Then, ReliefF-based feature selection was performed on the full dataset, and CV evaluation was executed using this reduced, fixed feature set. In the leakage-free pipeline, feature selection (including standardization of features) was performed in every CV iteration separately using only data from the training set, and this subset was then utilized to classify examples in the evaluation set.

### Evaluation pipeline

In this pipeline we utilized an ensemble classifier of decision trees enhanced with “GentleBoost” for classifying examples obtained by the above-described procedure. Specifically, in every CV iteration an ensemble classifier of simple decision trees (with maximum number of splits set to 5) was trained using the `fitensemble()` method in MATLAB, with setting the learning method to GentleBoost, the number of learning cycles to 100, and the learning rate to 0.1. Similarly to the data-driven pipeline, k-fold CV was implemented in leaky and leakage-free (i.e., subject-based) manners. Therefore, this pipeline was ultimately executed in four different ways: *i*) Leaky DP and leaky feature selection, *ii*) leaky DP but leakage-free feature selection, *iii*) leakage-free DP but leaky feature selection, and *iv*) completely leakage-free. In the main text we report results from cases *ii*) and *iv*) for comparability with the CNN-based pipeline, while details from all analysis scenarios are provided in the **Supplementary Material**.

### Single feature vector-based pipeline

This pipeline utilized the same feature set as obtained previously; however, for each subject to be characterized by a single feature vector, the obtained estimates were averaged over all epochs per subject. Accordingly, for this analysis the MSU dataset consisted of 84 examples (F^t^ E lffi.^1x3B4^), the RepOD dataset of 28 examples (F^t^ E lffi.^1x456^), and the SU-SZ dataset of 77 examples (F^t^ E lffi.^1x456^). Given the disproportionally large number of initial features compared to the drastically reduced sample size, we employed a more stringent feature selection and CV framework. However, since each subject contributed only one sample, DP-based leakage was not considered in this framework.

### Feature selection

As an initial step, features were standardized. Then, the ReliefF algorithm was utilized to eliminate features with negative weight and thus likely irrelevant for classification. Finally, the number of features were further reduced by employing the maximum relevance minimum redundancy (mRMR) algorithm [44]. The mRMR is an iterative algorithm that aims to select a subset of features that are maximally relevant to the target variable to be predicted, while at the same time minimally redundant amongst themselves. Here, we set mRMR to identify the top 10 most relevant features in all cases, which were then used for training and classification.

### Evaluation pipeline

Given the small samples, in this pipeline a support vector machine with radial basis function kernel was utilized for classification via the `fitcsvm()` Matlab function. To maximally utilize the available datasets, we employed an extensive leave-one-subject-out (eLOSO) CV scheme. In that, the evaluation set was always defined as a single pair of HC-SZ individuals with all remaining data used for training, and all possible pair combinations – i.e., N_Hc_ X N_sz_ where N_Hc_ and N_sz_ are the number of HC and SZ individuals in the dataset, respectively – were utilized as evaluation cases. Consequently, this pipeline only considered leakage related to feature selection.

In the leaky implementation, standardization and ReliefF+mRMR was performed first (i.e., outside-the-loop) to obtain the top 10 most relevant features, and eLOSO-CV was executed using this fixed feature subset in every iteration. In the leakage-free implementation, all feature selection steps – including standardization – was performed within every CV iteration (i.e., inside-the-loop) using only training set data, and the obtained feature subset was then selected from the evaluation set. Thus, the top 10 features varied for the different evaluation HC-SZ pairs in the CV evaluation. In both cases, classifier performance was characterized via classification accuracy averaged over CV iterations.

### EEG foundation model pipeline

EEG foundation models (FMs) employ a similar label-free, self-supervised learning strategy as modern large language and multimodal models [45] in an attempt to identify meaningful patterns and salient features in EEG data. The end-to-end training procedure of these models (often referred to as *pretraining*) generally involves a transforemer-based autoencoder architecture: an encoder projects native, unlabeled EEG data into a latent, high-dimensional embedding space, while a decoder attempts to reconstruct masked EEG segments (or their specific properties e.g., spectral characteristics) from these embeddings. Given sufficient amount of diverse pretraining data, a product of this procedure is an encoder that compresses EEG into meaningful embeddings, which meaningfully capture the intrinsic properties of brain activity. These embeddings then can be used in various supervised classification tasks (often termed *downstream* tasks) such as brain-computer interface applications or clinical decision making support, and thus FMs can be considered as “*off-the-shelf feature extractors*” [45]. While the pretraining is expensive and relies on thousands of hours of unlabeled EEG data, the downstream classifiers are usually simple and lightweight compared to the encoder itself. Overall, state-of-the-art FMs are built to recognize and extract inherent properties of EEG collected from a diverse set of conditions and experimental setups, to generalize well to unique or custom electrode montages, and to circumvent the bottleneck of manual data annotation, and therefore they hold great promise for future research dealing with EEG-based applications [45].

Conceptually, the embedding procedure performed by the pretrained encoder is feature extraction itself, and therefore downstream tasks do not commonly involve explicit FS. However, since the downstream classifier is ultimately trained to separate embedding vectors, it can be subject to DP-related effects same as any other classification approach. To the best of our knowledge, no previous study has investigated this issue yet on downstream classification tasks. For this purpose, we opted to utilize REVE [46], a recently proposed FM pretrained on over 60,000 hours of both clinical and other EEG data. Note that according to the current documentation of REVE, none of the three SZ-EEG datasets considered in our study were part of the REVE pretraining dataset (see Appendix B in [46]).

### EEG preparation

Preprocessed EEG was further prepared to align with data used for pre-training REVE [46]. In that, EEG was first resampled to have 200 Hz sampling rate (upsampling for MSU, downsampling for RepOD and SU-SZ) using the `resample()` Matlab function, and continuous recordings for each channel were standardized to have zero mean and unit variance (subject-level). Then, recordings were segmented into 5 second-long, non-overlapping epochs. Finally, epochs with values outside the ±15 standard deviation regime were excluded. This resulted in a final sample size of 1008, 3909 and 1500 EEG epochs for the MSU, RepOD and SU-SZ datasets, respectively, which were subsequently used as inputs to the REVE encoder.

### Feature extraction

We used the REVE base model (accessible at https://huggingface.co/brain-bzh/reve-base) to transform the 5-second EEG epochs into 512-dimensional embedding vectors. Parameters of the REVE model were kept frozen during the procedure. Since REVE internally segments EEG into 1-second non-overlapping vectors, this procedure yielded 5 embedding vectors per EEG epoch, which were then aggregated into a single feature vector of size 1 X 512 via mean-pooling. Accordingly, the final samples for the MSU, RepOD and SU-SZ datasets were of size 1008 X 512, 3909 X 512 and 1500 X 512, respectively.

### Evaluation pipeline

To remain consistent with standard practices in FM performance evaluation [45], we performed *linear probing* by using SVMs with linear kernels for classification. For this, we used the `fitcsvm()` Matlab function with ‘`KernelFunction`‘ set to ‘`linear`‘ and ‘`Standardize`‘ set to `true`. The full, 512-dimensional embedding vectors were used as model input and no FS was performed. Similar to the CNN-based evaluation pipeline, stratified 10-fold CV was utilized for the MSU and SU-SZ datasets, while 7-fold CV for the RepOD dataset in both leaky DP (random split) or leakage-free (subject-based split) implementations.

### Performance evaluation

In the primary analyses, model performances in every CV iteration were characterized via classification accuracy i.e., the proportion of correctly predicted evaluation samples to the total number of evaluation samples. For a more comprehensive assessment, ground truth labels and predictions were pooled from all CV iterations and performance was characterized through eight standard metrics [78]. With TP, TN, FP and FN denoting true positives, true negatives, false positives and false negatives, respectively, the following measures were computed:

1. Balanced Accuracy (%): 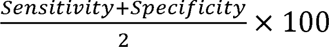
2. Specificity (%): 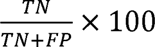
3. Sensitivity (%): 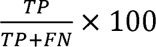
4. Precision or Positive Predictive Value (%): 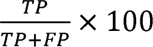
5. F1 Score (%): 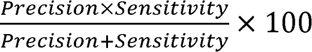
6. Matthew’s Correlation Coefficient (%): 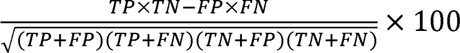
7. Cohen’s Kappa [79]
8. receiver operator characteristic area under the curve (ROC-AUC) [80].

### Riemannian geometry and assessment of data variability

The toolset of Riemannian geometry provides a suitable way to address variability in multi-channel EEG data [81]. In the Riemannian framework, multivariate data is first transformed into covariance matrix representation (being symmetric positive definite) and then the distance between different matrices can be captured via the Riemannian distance (or geodesic) that is invariant under affine transformations [82]. We analyzed the EEG epochs utilized in the CNN-based evaluation, and each epoch was transformed into trace-normalized covariance matrix structure using the Ledoit-Wolf method [83]. In each dataset, individual covariance matrices were re-centered using a reference matrix obtained as the Riemannian (Karcher) mean of all covariance matrices in the dataset [84]. This step is equivalent to standardization for univariate signals, and its goal was to match distributions among different subjects and thus minimize between-subject variability. Then, the following prototype (i.e., center of mass) matrices were computed from corresponding epochs as in [84]: *i*) subject centroids, obtained as the Riemannian mean of covariance matrices from that subject, *ii*) group centroids, obtained as the Riemannian mean of all covariance matrices from HC/SZ groups, and *iii*) global centroids, obtained as the Riemannian mean of all covariances in the dataset, irrespective of group label. Within-subject variability was defined as the average Riemannian distance of a subject’s samples to their own centroid, while between-subject and global variability for each subject was defined as the average Riemannian distance of the subjects’ samples to their respective group centroid and the global dataset centroid, respectively. Note that this approach is analogous to within- and between-class variability assessment in traditional clustering analysis, however the Riemannian approach allows for exploiting multivariate interactions in the data via the covariance representation and thus is more suitable to address variability in multi-channel EEG data. For more details on the application of Riemannian geometry on EEG data and related concepts, please see [81] or [85].

For variability in extracted EEG features, the t-SNE [47] method was utilized, which is a common visualization and analysis technique used in data science. In essence, t-SNE employs a non-linear approach to map high-dimensional data to a lower-dimensional (e.g., 2D) space while preserving local similarities, and thus it is ideal in intuitive exploration of internal data structures such as clustering of samples or separability of classes [47]. We analyzed the EEG feature vectors derived for the ensemble-based evaluation for this analysis. All features were standardized using all data in the given dataset to reduce distribution shifts across subjects and ensure similar scaling of all features. The `tsne()` Matlab function was utilized with default perplexity value of 30 to obtain two-dimensional embeddings of full feature vectors.

Note that in both analyses, data processing steps (re-centering, standardization) were employed to reduce between-subject distribution shifts in the data and therefore the resulting outcomes should be considered as conservative estimates on data variability.

### Statistical testing

The performance drops from leaky to leakage-free implementations were characterized as µ(ACC_L_) - µ(ACC_LF_) where µ(·) denotes the average taken over CV iterations, and ACC_L_ and ACC_LF_ refers to classification accuracy in the leaky and leakage-free implementation, respectively. Confidence intervals for this difference were obtained via bootstrapping. In that, for both the leaky and leakage-free fold accuracies, an equal number of samples to the number of fold iterations (i.e., 10 in 10-fold CV, N_Hc_ X N_sz_ in eLOSO-CV) were chosen randomly with replacement, and the difference between average bootstrap accuracies were computed. This procedure was repeated *K*=10,000 times. Then, the 2.5% and 97.5% percentiles of the obtained distribution were chosen as confidence interval boundaries. The statistical significance of the difference was estimated via permutation testing. In that, for each evaluation scheme, the leaky and leakage-free fold accuracies were first concatenated, then the complete set was shuffled before re-partitioning into leaky and leakage-free outcomes, with finally the difference between average leaky and leakage-free CV accuracies computed from these permuted sets. The procedure was repeated for *K*=10,000 times, and the non-parametric *p*-value was defined as the proportion of permuted differences larger than the observed true difference. In the analysis of data variability, within-subject, between-subject and global variability values were contrasted using paired t tests or Wilcoxon signed rank tests, depending on data normality. Outcomes were adjusted for multiple comparisons (*n*=3) using Bonferroni’s method at a = 0.05.

## Data Availability

Data availability
No new physiological data was generated throughout this study as only openly available datasets were analyzed. The MSU dataset can be accessed at the following hyperlink: http://brain.bio.msu.ru/eeg_schizophrenia.htm. The RepOD dataset can be accessed at the following hyperlink: https://doi.org/10.18150/repod.0107441. The SU-SZ dataset can be accessed at the following hyperlink: https://doi.org/10.5281/zenodo.14808295.
Code availability
All code, including but not limited to scripts and corresponding functions performing data pre-processing, feature extraction, classification and results presentation will be made available without restriction upon publication of this article at the following GitHub repository: https://github.com/samuelracz/schizophrenia_EEG_detection_leakage.

https://doi.org/10.18150/repod.0107441

https://doi.org/10.5281/zenodo.14808295

## References

1. Solmi, M., G. Seitidis, D. Mavridis, C.U. Correll, E. Dragioti, S. Guimond, et al., Incidence, prevalence, and global burden of schizophrenia - data, with critical appraisal, from the Global Burden of Disease (GBD) 2019. Mol Psychiatry, 2023.

2. Misiak, B., J. Samochowiec, K. Kowalski, W. Gaebel, C.L. Bassetti, A. Chan, et al., The future of diagnosis in clinical neurosciences: Comparing multiple sclerosis and schizophrenia. European Psychiatry, 2023. 66(1): p. e58.

3. Oso, T.A., O.J. Okesanya, U.O. Adebayo, K.B. Obadeyi, S.S. Musa, M.M. Ahmed, et al., Advancing our understanding of schizophrenia: insights from recent research, emerging therapies, and future directions. Exploration of Neuroscience, 2025. 4: p. 1006112.

4. Etkin, A. and D.H. Mathalon, Bringing imaging biomarkers into clinical reality in psychiatry. JAMA psychiatry, 2024. 81(11): p. 1142–1147.

5. Popov, T., M. Tröndle, Z. Baranczuk Turska, C. Pfeiffer, S. Haufe, and N. Langer, Test–retest reliability of resting state EEG in young and older adults. Psychophysiology, 2023. 60(7): p. e14268.

6. Rahul, J., D. Sharma, L.D. Sharma, U. Nanda, and A.K. Sarkar, A systematic review of EEG based automated schizophrenia classification through machine learning and deep learning. Frontiers in Human Neuroscience, 2024. 18: p. 1347082.

7. Saha, A., S. Park, Z.W. Geem, and P.K. Singh, Schizophrenia detection and classification: A systematic review of the last decade. Diagnostics, 2024. 14(23): p. 2698.

8. Sun, J., R. Cao, M. Zhou, W. Hussain, B. Wang, J. Xue, et al., A hybrid deep neural network for classification of schizophrenia using EEG Data. Scientific Reports, 2021. 11(1): p. 4706.

9. Shoeibi, A., D. Sadeghi, P. Moridian, N. Ghassemi, J. Heras, R. Alizadehsani, et al., Automatic diagnosis of schizophrenia in EEG signals using CNN-LSTM models. Frontiers in neuroinformatics, 2021. 15: p. 777977.

10. Aslan, Z. and M. Akin, A deep learning approach in automated detection of schizophrenia using scalogram images of EEG signals. Physical and Engineering Sciences in Medicine, 2022. 45(1): p. 83–96.

11. Bagherzadeh, S., M.S. Shahabi, and A. Shalbaf, Detection of schizophrenia using hybrid of deep learning and brain effective connectivity image from electroencephalogram signal. Computers in Biology and Medicine, 2022. 146: p. 105570.

12. Ilakiyaselvan, N., A.N. Khan, and A. Shahina, Reconstructed phase space portraits for detecting brain diseases using deep learning. Biomedical Signal Processing and Control, 2022. 71: p. 103278.

13. Sharma, G. and A.M. Joshi, SzHNN: A novel and scalable deep convolution hybrid neural network framework for schizophrenia detection using multichannel EEG. IEEE Transactions on Instrumentation and Measurement, 2022. 71: p. 1–9.

14. Urkin, B., J. Parnas, A. Raballo, and D. Koren, Schizophrenia spectrum disorders: an empirical benchmark study of real-world diagnostic accuracy and reliability among leading international psychiatrists. Schizophrenia Bulletin Open, 2024. 5(1): p. sgae012.

15. Sharma, G., A.M. Joshi, D. Yadav, and S.P. Mohanty, A smart healthcare framework for accurate detection of schizophrenia using multichannel EEG. IEEE Transactions on Instrumentation and Measurement, 2023. 72: p. 1–9.

16. Grover, N., A. Chharia, R. Upadhyay, and L. Longo, Schizo-Net: A novel Schizophrenia Diagnosis framework using late fusion multimodal deep learning on Electroencephalogram-based Brain connectivity indices. IEEE Transactions on Neural Systems and Rehabilitation Engineering, 2023. 31: p. 464–473.

17. Kapoor, S. and A. Narayanan, Leakage and the reproducibility crisis in machine-learning-based science. Patterns, 2023. 4(9).

18. Sasse, L., E. Nicolaisen-Sobesky, J. Dukart, S. Eickhoff, M. Götz, S. Hamdan, et al., Overview of leakage scenarios in supervised machine learning. Journal of Big Data, 2025. 12(1): p. 135.

19. Del Pup, F., A. Zanola, L.F. Tshimanga, A. Bertoldo, L. Finos, and M. Atzori, The role of data partitioning on the performance of EEG-based deep learning models in supervised cross-subject analysis: a preliminary study. Computers in Biology and Medicine, 2025. 196: p. 110608.

20. Shim, M., S.-H. Lee, and H.-J. Hwang, Inflated prediction accuracy of neuropsychiatric biomarkers caused by data leakage in feature selection. Scientific Reports, 2021. 11(1): p. 7980.

21. Kaufman, S., S. Rosset, C. Perlich, and O. Stitelman, Leakage in data mining: Formulation, detection, and avoidance. ACM Transactions on Knowledge Discovery from Data (TKDD), 2012. 6(4): p. 1–21.

22. Lee, H.-T., H.-R. Cheon, S.-H. Lee, M. Shim, and H.-J. Hwang, Risk of data leakage in estimating the diagnostic performance of a deep-learning-based computer-aided system for psychiatric disorders. Scientific Reports, 2023. 13(1): p. 16633.

23. Starcke, J., J. Spadafora, J. Spadafora, P. Spadafora, and M. Toma, The Effect of Data Leakage and Feature Selection on Machine Learning Performance for Early Parkinson’s Disease Detection. Bioengineering, 2025. 12(8): p. 845.

24. Young, V.M., S. Gates, L.Y. Garcia, and A. Salardini, Data Leakage in Deep Learning for Alzheimer’s Disease Diagnosis: A Scoping Review of Methodological Rigor and Performance Inflation. Diagnostics, 2025. 15(18): p. 2348.

25. Rosenblatt, M., L. Tejavibulya, R. Jiang, S. Noble, and D. Scheinost, Data leakage inflates prediction performance in connectome-based machine learning models. Nature Communications, 2024. 15(1): p. 1829.

26. Brookshire, G., J. Kasper, N.M. Blauch, Y.C. Wu, R. Glatt, D.A. Merrill, et al., Data leakage in deep learning studies of translational EEG. Frontiers in Neuroscience, 2024. 18: p. 1373515.

27. Ranjan, R., B.C. Sahana, and A.K. Bhandari, Deep learning models for diagnosis of schizophrenia using EEG signals: emerging trends, challenges, and prospects. Archives of Computational Methods in Engineering, 2024. 31(4): p. 2345–2384.

28. Uyanik, H., A. Sengur, M. Salvi, R.S. Tan, J.H. Tan, and U.R. Acharya, Automated Detection of Neurological and Mental Health Disorders Using EEG Signals and Artificial Intelligence: A Systematic Review. Wiley Interdisciplinary Reviews: Data Mining and Knowledge Discovery, 2025. 15(1): p. e70002.

29. Jiang, S., Q. Jia, Z. Peng, Q. Zhou, Z. An, J. Chen, et al., Can artificial intelligence be the future solution to the enormous challenges and suffering caused by Schizophrenia? Schizophrenia, 2025. 11(1): p. 32.

30. Khare, S.K., V. Bajaj, and U.R. Acharya, SchizoNET: a robust and accurate Margenau–Hill time-frequency distribution based deep neural network model for schizophrenia detection using EEG signals. Physiological Measurement, 2023. 44(3): p. 035005.

31. Oh, S.L., J. Vicnesh, E.J. Ciaccio, R. Yuvaraj, and U.R. Acharya, Deep Convolutional Neural Network Model for Automated Diagnosis of Schizophrenia Using EEG Signals. Applied Sciences-Basel, 2019. 9(14).

32. Aydemir, E., S. Dogan, M. Baygin, C.P. Ooi, P.D. Barua, T. Tuncer, et al. CGP17Pat: automated schizophrenia detection based on a cyclic group of prime order patterns using EEG signals. in Healthcare. 2022. MDPI.

33. Hassan, F., S.F. Hussain, and S.M. Qaisar, Fusion of multivariate EEG signals for schizophrenia detection using CNN and machine learning techniques. Information Fusion, 2023. 92: p. 466–478.

34. Li, B., J. Wang, Z. Guo, and Y. Li, Automatic detection of schizophrenia based on spatial–temporal feature mapping and LeViT with EEG signals. Expert Systems with Applications, 2023. 224: p. 119969.

35. Borisov, S., A.Y. Kaplan, N. Gorbachevskaya, and I. Kozlova, Analysis of EEG structural synchrony in adolescents with schizophrenic disorders. Human Physiology, 2005. 31(3): p. 255–261.

36. Olejarczyk, E. and W. Jernajczyk, Graph-based analysis of brain connectivity in schizophrenia. PLoS One, 2017. 12(11): p. e0188629.

37. Ford, J.M., V.A. Palzes, B.J. Roach, and D.H. Mathalon, Did I do that? Abnormal predictive processes in schizophrenia when button pressing to deliver a tone. Schizophrenia bulletin, 2014. 40(4): p. 804–812.

38. Racz, F.S., K. Farkas, M. Becske, H. Molnar, Z. Fodor, P. Mukli, et al., Reduced temporal variability of cortical excitation/inhibition ratio in schizophrenia. Schizophrenia (Heidelb), 2025. 11(1): p. 20.

39. He, K., X. Zhang, S. Ren, and J. Sun. Deep residual learning for image recognition. in Proceedings of the IEEE conference on computer vision and pattern recognition. 2016.

40. Shin, H.-C., H.R. Roth, M. Gao, L. Lu, Z. Xu, I. Nogues, et al., Deep convolutional neural networks for computer-aided detection: CNN architectures, dataset characteristics and transfer learning. IEEE transactions on medical imaging, 2016. 35(5): p. 1285–1298.

41. Chen, T. and C. Guestrin. Xgboost: A scalable tree boosting system. in Proceedings of the 22nd acm sigkdd international conference on knowledge discovery and data mining. 2016.

42. Urbanowicz, R.J., M. Meeker, W. La Cava, R.S. Olson, and J.H. Moore, Relief-based feature selection: Introduction and review. Journal of biomedical informatics, 2018. 85: p. 189–203.

43. Sugden, R.J. and P. Diamandis, Generalizable electroencephalographic classification of Parkinson’s disease using deep learning. Informatics in Medicine Unlocked, 2023. 42: p. 101352.

44. Peng, H.C., F.H. Long, and C. Ding, Feature selection based on mutual information: Criteria of max-dependency, max-relevance, and min-redundancy. Ieee Transactions on Pattern Analysis and Machine Intelligence, 2005. 27(8): p. 1226–1238.

45. Kuruppu, G., N. Wagh, V. Kremen, and Y. Varatharajah, EEG foundation models: a critical review of current progress and future directions. Journal of neural engineering, 2026. 23(2): p. 021001.

46. Ouahidi, Y.E., J. Lys, P. Thölke, N. Farrugia, B. Pasdeloup, V. Gripon, et al., REVE: A Foundation Model for EEG--Adapting to Any Setup with Large-Scale Pretraining on 25,000 Subjects. arXiv preprint arXiv:2510.21585, 2025.

47. van der Maaten, L. and G. Hinton, Visualizing Data using t-SNE. Journal of Machine Learning Research, 2008. 9: p. 2579–2605.

48. Raiaan, M.A.K., S. Sakib, N.M. Fahad, A. Al Mamun, M.A. Rahman, S. Shatabda, et al., A systematic review of hyperparameter optimization techniques in Convolutional Neural Networks. Decision Analytics Journal, 2024. 11: p. 100470.

49. Wojciuk, M., Z. Swiderska-Chadaj, K. Siwek, and A. Gertych, Improving classification accuracy of fine-tuned CNN models: Impact of hyperparameter optimization. Heliyon, 2024. 10(5).

50. Khare, S.K., V. Bajaj, and U.R. Acharya, SPWVD-CNN for automated detection of schizophrenia patients using EEG signals. IEEE Transactions on Instrumentation and Measurement, 2021. 70: p. 1–9.

51. Hosseini, M., M. Powell, J. Collins, C. Callahan-Flintoft, W. Jones, H. Bowman, et al., I tried a bunch of things: The dangers of unexpected overfitting in classification of brain data. Neuroscience & Biobehavioral Reviews, 2020. 119: p. 456–467.

52. Supakar, R., P. Satvaya, and P. Chakrabarti, A deep learning based model using RNN-LSTM for the detection of schizophrenia from EEG data. Computers in Biology and Medicine, 2022. 151: p. 106225.

53. Shoeibi, A., M. Jafari, D. Sadeghi, R. Alizadehsani, H. Alinejad-Rokny, A. Beheshti, et al. Early diagnosis of schizophrenia in EEG signals using one dimensional transformer model. in International work-conference on the interplay between natural and artificial computation. 2024. Springer.

54. Ouzzani, M., H. Hammady, Z. Fedorowicz, and A. Elmagarmid, Rayyan—a web and mobile app for systematic reviews. Systematic reviews, 2016. 5(1): p. 210.

55. Gorbachevskaya, N. and S.V. Borisov, EEG of healthy adolescents and adolescents with symptoms of schizophrenia. 2019.

56. Kulaichev, A.P. and N.L. Gorbachevskaya, Differentiation of norm and disorders of schizophrenic spectrum by analysis of EEG correlation synchrony. Journal of Experimental & Integrative Medicine, 2013. 3(4).

57. Jasper, H.H., The ten-twenty electrode system of the International Federation. Electroencephalogr. Clin. Neurophysiol., 1958. 10: p. 370–375.

58. Delorme, A. and S. Makeig, EEGLAB: an open source toolbox for analysis of single-trial EEG dynamics including independent component analysis. Journal of Neuroscience Methods, 2004. 134(1): p. 9–21.

59. Bigdely-Shamlo, N., T. Mullen, C. Kothe, K.-M. Su, and K.A. Robbins, The PREP pipeline: standardized preprocessing for large-scale EEG analysis. Frontiers in neuroinformatics, 2015. 9: p. 16.

60. Hyvarinen, A. and E. Oja, Independent component analysis: algorithms and applications. Neural Netw, 2000. 13(4-5): p. 411–430.

61. Winkler, I., S. Brandl, F. Horn, E. Waldburger, C. Allefeld, and M. Tangermann, Robust artifactual independent component classification for BCI practitioners. Journal of Neural Engineering, 2014. 11(3).

62. Bhatt, D., C. Patel, H. Talsania, J. Patel, R. Vaghela, S. Pandya, et al., CNN variants for computer vision: History, architecture, application, challenges and future scope. Electronics, 2021. 10(20): p. 2470.

63. Rajwal, S. and S. Aggarwal, Convolutional neural network-based EEG signal analysis: A systematic review. Archives of Computational Methods in Engineering, 2023. 30(6): p. 3585–3615.

64. Deng, J., W. Dong, R. Socher, L.-J. Li, K. Li, and L. Fei-Fei. Imagenet: A large-scale hierarchical image database. in 2009 IEEE conference on computer vision and pattern recognition. 2009. Ieee.

65. Hosseini, M.-P., A. Hosseini, and K. Ahi, A review on machine learning for EEG signal processing in bioengineering. IEEE reviews in biomedical engineering, 2020. 14: p. 204–218.

66. Santos-Mayo, L., L.M. San-José-Revuelta, and J.I. Arribas, A computer-aided diagnosis system with EEG based on the P3b wave during an auditory odd-ball task in schizophrenia. IEEE transactions on biomedical engineering, 2016. 64(2): p. 395–407.

67. Racz, F.S., O. Stylianou, P. Mukli, and A. Eke, Multifractal and Entropy-Based Analysis of Delta Band Neural Activity Reveals Altered Functional Connectivity Dynamics in Schizophrenia. Frontiers in Systems Neuroscience, 2020. 14.

68. Guo, Z., L. Wu, Y. Li, and B. Li. Deep neural network classification of EEG data in schizophrenia. in 2021 IEEE 10th Data Driven Control and Learning Systems Conference (DDCLS). 2021. IEEE.

69. Dimitriadis, S.I., *Reconfiguration of* α*mplitude driven dominant coupling modes (DoCM) mediated by* α*-band in adolescents with schizophrenia spectrum disorders*. Progress in Neuro-Psychopharmacology and Biological Psychiatry, 2021. 108: p. 110073.

70. De Miras, J.R., A.J. Ibáñez-Molina, M.F. Soriano, and S. Iglesias-Parro, Schizophrenia classification using machine learning on resting state EEG signal. Biomedical Signal Processing and Control, 2023. 79: p. 104233.

71. Hjorth, B., EEG analysis based on time domain properties. Electroencephalography and clinical neurophysiology, 1970. 29(3): p. 306–310.

72. Márton, L., S.T. Brassai, L. Bakó, and L. Losonczi, Detrended fluctuation analysis of EEG signals. Procedia Technology, 2014. 12: p. 125–132.

73. Kaposzta, Z., A. Czoch, O. Stylianou, K. Kim, P. Mukli, A. Eke, et al., Real-Time Algorithm for Detrended Cross-Correlation Analysis of Long-Range Coupled Processes. Frontiers in Physiology, 2022. 13.

74. Bandt, C. and B. Pompe, Permutation entropy: A natural complexity measure for time series. Physical Review Letters, 2002. 88(17).

75. Montez, T., K. Linkenkaer-Hansen, B.W. van Dijk, and C.J. Stam, Synchronization likelihood with explicit time-frequency priors. Neuroimage, 2006. 33(4): p. 1117–1125.

76. Stam, C.J., G. Nolte, and A. Daffertshofer, Phase lag index: Assessment of functional connectivity from multi channel EEG and MEG with diminished bias from common sources. Human Brain Mapping, 2007. 28(11): p. 1178–1193.

77. Rubinov, M. and O. Sporns, Complex network measures of brain connectivity: Uses and interpretations. NeuroImage, 2010. 52(3): p. 1059–1069.

78. Bretones, C.S., C.R. Parra, J. Cascón, A.L. Borja, and J.M. Sotos, Automatic identification of schizophrenia employing EEG records analyzed with deep learning algorithms. Schizophrenia Research, 2023. 261: p. 36–46.

79. Cohen, J., A coefficient of agreement for nominal scales. Educational and psychological measurement, 1960. 20(1): p. 37–46.

80. Hanley, J.A. and B.J. McNeil, The meaning and use of the area under a receiver operating characteristic (ROC) curve. Radiology, 1982. 143(1): p. 29–36.

81. Kumar, S., H. Alawieh, F.S. Racz, R. Fakhreddine, and J.D.R. Millán, Transfer learning promotes acquisition of individual BCI skills. PNAS Nexus, 2024. 3(2): p. pgae076.

82. Moakher, M., A differential geometric approach to the geometric mean of symmetric positive-definite matrices. SIAM journal on matrix analysis and applications, 2005. 26(3): p. 735–747.

83. Ledoit, O. and M. Wolf, A well-conditioned estimator for large-dimensional covariance matrices. Journal of multivariate analysis, 2004. 88(2): p. 365–411.

84. Kumar, S., F. Yger, and F. Lotte. Towards adaptive classification using Riemannian geometry approaches in brain-computer interfaces. in *2019 7th International Winter Conference on Brain-Computer Interface (BCI)*. 2019. IEEE.

85. Racz, F.S., S. Kumar, Z. Kaposzta, H. Alawieh, D.H. Liu, R. Liu, et al., Combining detrended cross-correlation analysis with Riemannian geometry-based classification for improved brain-computer interface performance. Front Neurosci, 2024. 18: p. 1271831.

