## Supplementary Material for "Information Leakage and Performance Overestimation in EEG-Based Schizophrenia Detection: Evidence from Literature and Empirical Analyses"

**Supplementary Table S1** summarizes previous literature aiming for automated classification of healthy controls (HC) against patients with schizophrenia (SZ) from EEG data using machine- and deep learning (ML/DL) methods. Our literature screening procedure (see below) yielded 95 publications with a total number of 119 evaluation pipelines, as some of these analyzed multiple datasets and/or implemented multiple evaluation pipelines. Performance is characterized as classification accuracy (i.e., the proportion of correctly classified samples to the total number of samples) in all cases, unless noted otherwise. Notably, many of these previous works explore more than one ML/DL technique for classification; in these cases, only the highest reported performance is included here. Multiple performance outcomes are included if *i*) datasets were evaluated in both leaky- and leakage-free schemes, *ii*) multiple datasets are evaluated, or *iii*) relevant otherwise (e.g., mean vs. highest accuracy).

**Supplementary Table S1.** Summary table of studies classifying HC vs. SZ using EEG data. Throughout this table, ‘N/A’ denotes if the field was not applicable for the corresponding study, while ‘UNK’ indicates if the information was not available or unclear given the provided description by the paper. For the FS and DP columns, ‘+’ and ‘-‘ denotes the presence or absence of leakage, respectively. For the CA column, ‘+’ indicates available code, while ‘-‘ otherwise. HC: healthy control; SZ: schizophrenia; FS: feature selection; DP: data partitioning; CA: code availability; RS: resting state; EO: eyes-open; EC: eyes-closed; ERP: event-related potential.

| **Paper** | **Dataset** | **Condition** | **Model** | **Features** | **Selection** | **FS** | **Evaluation** | **DP** | **Performance** | **CA** |
| --- | --- | --- | --- | --- | --- | --- | --- | --- | --- | --- |
| Sabeti et al., 2009 [1] | Private  HC: 20  SZ: 20 | RS (EO) | LDA, AdaBoost | Non-linear, time domain | Genetic Programming | +/- | LOSO-CV | - | 86%-89% (- FS)  89%-91% (+ FS) | - |
| Hiesh et al., 2013 [2] | Private  HC: 5  SZ: 5 | Auditory stimulation | SVM | Non-linear, time domain | Genetic Algorithm | + | UNKs | UNK | 88.24% | - |
| Neuhaus et al., 2013 [3] | Private  HC: 24  SZ: 24 | Visual and auditory oddball | 11 classifiers (KNN best) | ERP features | Feature ranking, 10-fold CV using all feature subsets | + | 90-10 train/test 50 times | - | 72.4% | - |
| Parvinnia et al., 2014 [4] | Private  HC: 18  SZ: 13 | RS (EO) | WDNN | Autoregressive, spectral power, fractal features | WDNN | - | LOSO-CV | - | 95.32% | - |
| Sui et al., 2014 [5] | Private  HC: 53  SZ: 48 | RS (EO) | SVM | Spectral power (also sMRI and fMRI) | SVM-RFE | + | 10-fold CV | - | 74%-80% | - |
| Dvey-Aharon et al., 2015 [6] | Private  HC: 25  SZ: 25 | Contextual processing task | KNN | Stockwell time-frequency features | Channel, time window, frequency band combinations | + | LOSO-CV | - | 91.5%-93.9% (using top 5 channels) | - |
| Ravan et al., 2015 [7] | Private  HC: 66  SZ: 47 | Auditory oddball | FCM | Auto- and cross-spectral density | Regularized feature selection with LOO-CV | + | LOO-CV | - | 85% | - |
| Santos-Mayo et al., 2016 [8] | Private  HC: 31  SZ: 16 | Auditory oddball | MLP, SVM | ERP features | 3 techniques (J5, MIFS, DISR) | + | 60-10-30 and 75-25 train/test split | - | 93.42% | - |
| Shim et al., 2016 [9] | Private  HC: 34  SZ: 34 | Auditory oddball | SVM | ERP features (sensor and source space) | Fisher score (1-20 features) | + | LOSO-CV | - | 88.24% (highest)  78.24% (mean) | - |
| Chu et al., 2017 [10] | Private  HC: 40  SZ: 40  CHR: 40 | RS (EO) | CNN, ANN, RNN + RF | Filtered EEG, Power spectral features | N/A | N/A | k-fold CV | UNK | 81.6%-99.2% | - |
| Liu et al., 2017 [11] | Private  HC: 40  SZ: 40  CHR: 40 | RS (EC) | SVM, DT, KNN, RF, NB | Sample covariance matrix, linear eigenvalue statistics | N/A | N/A | 5-fold CV | - | 91.16% | - |
| Piryatinska et al., 2017 [12] | MSU  HC: 39  SZ: 45 | RS (EC) | RF, SVM | ε-complexity coefficients, power spectral features | Feature ranking using Gini index | + | OOB, 10-fold CV | + | 81%-85.3% | - |
| Alimardani et al., 2018 [13] | Private  BD: 23  SZ: 23 | SSVEP | LDA, QDA, SVM, KNN, LRA | SSVEP features | Fisher score (1-10 features) | + | LOSO-CV | - | 91.3% best performance | - |
| Buettner et al., 2019 [14] | RepOD  HC: 14  SZ: 14 | RS (EC) | RF | Power spectral features | Power spectral features | N/A | 75-25 train/test split | - | 71.43%  80% (balanced accuracy) | - |
| Devia et al., 2019 [15] | Private  HC: 9  SZ: 11 | Free image exploration | LDA | ERP features | N/A | N/A | 80-20 train/test 300 times | - | 71% | - |
| Jamunah et al., 2019 [16] | RepOD  HC: 14  SZ: 14 | RS (EC) | DT, LDA, KNN, PNN, SVM | Non-linear features | t-test | + | UNK | UNK | 92.91% | - |
| Li et al., 2019 [17] | Private  HC: 25  SZ: 23 | RS (EC), P300 task | LDA, SVM | P300 and brain network features (SPN) | N/A | N/A | LOO-CV | - | 90.48% | - |
| Naira and Jos, 2019 [18] | MSU  HC: 39  SZ: 45 | RS (EC) | CNN | Pearson correlation matrices | N/A | N/A | 80-20 train/test split | + | 90% | - |
| Oh et al., 2019 [19] | RepOD  HC: 14  SZ: 14 | RS (EC) | CNN+ | Processed EEG signals | N/A | N/A | LOSO-CV  10-fold CV | +/- | 98.07% (EB)  81.26% (SB) | - |
| Phang et al., 2019 [20] | MSU  HC: 39  SZ: 45 | RS (EC) | DNN-DBN, SVM, LDA, QDA | Connectivity map, graph theoretical measures | N/A | N/A | 60-15-25 train/val/test split | - | 95% (highest) | - |
| Ahmedt-Aristizabal et al., 2020 [21] | Private  RSZ: 1287  TD: 127 | Auditory oddball | KNN, DT, SVM, CNN, RNN, R-CNN | ERP features, processed EEG | N/A | N/A | 5-fold CV  External validation (A3) | - | 89.98% (val)  69.80% (test) | - |
| Aslan and Akin, 2020 [22] | MSU  HC: 39  SZ: 45  RepOD  HC: 14  SZ: 14 | RS (EC)  RS (EC) | CNN (VGG-16) | Spectrogram features | N/A | N/A | 80-20 train/test split | +  + | 95% (MSU)  97% (RepOD) | - |
| Buettner et al., 2020 [23] | RepOD  HC: 14  SZ: 14 | RS (EC) | RF | Power spectral features | N/A | N/A | 10-fold CV | + | 96.77% | - |
| Calhas et al., 2020 [24] | MSU  HC: 39  SZ: 45 | RS (EC) | SVM, RF, XGB, NB, KNN | SNN trained on spectrogram features, output vectors used as features | SNN training | UNK | LOO-CV | - | 95% | + |
| Goshvarpour and Goshvarpour, 2020 [25] | RepOD  HC: 14  SZ: 14 | RS (EC) | PNN | Time domain, non-linear, fractal features | Feature fusion using ANOVA F-statistics | + | k-fold CV (k=2, 5, 10) | + | 100% | - |
| Khare et al., 2020 [26] | Kaggle  HC: 32  SZ: 49 | Auditory ERP task | KNN, LDA, ensemble, SVM, DT | Time domain features from EWT transformed EEG | Kruskal-Wallis test | + | 10-fold CV | + | 88.7% | - |
| Kim et al., 2020 [27] | Private  HC: 119  SZ: 119 | RS (EC) | LDA | Connectivity features from source-space EEG | Sequential forward selection | + | 10-fold CV 10 times | + | 80.66% (best feature set) | - |
| Krishnan et al., 2020 [28] | RepOD  HC: 14  SZ: 14 | RS (EC) | KNN, LDA, NB, SVM, RF, GB | Entropy measures from MEMD transformed EEG | RF-RFE  (full, but N/A as well) | + | k-fold CV (k=2, 4, 5, 10) | + | 93% (SVM)  Same for RFE or N/A | - |
| Nikhil et al., 2020 [29] | RepOD  HC: 14  SZ: 14 | RS (EC) | LSTM | Non-linear and time domain features | N/A | N/A | 89-11 train/test split | + | 99% | - |
| Phang et al., 2020 [30] | MSU  HC: 39  SZ: 45 | RS (EC) | SVM, CNN, RNN | Time domain, power spectral, connectivity features | HT on validation set | N/A | Single 60-20-20 train/val/test split | - | 91.69% (CNN) | - |
| Prabhakar et al., 2020a [31] | RepOD  HC: 14  SZ: 14 | RS (EC) | AdaBoost and Bayesian classifiers | PLS regression, EM-PCA, isomap features | 4 nature inspired algorithms | + | 10-fold CV | + | 98.77% | - |
| Prabhakar et al., 2020b [32] | RepOD  HC: 14  SZ: 14 | RS (EC) | ANN, QDA, SVM, LR, FLDA, KNN | Time domain, fractal, non-linear features | 4 nature inspired algorithms | + | 5-fold CV, 70%-30% split | + | 87.54%-92.17% | - |
| Racz et al., 2020 [33] | RepOD  HC: 14  SZ: 14 | RS (EC) | RF | Dynamic connectivity features | Hyperparameter tuning | + | LOSO-CV | - | 89.29% | - |
| Shalbaf et al., 2020 [34] | RepOD  HC: 14  SZ: 14 | RS (EC) | 4 pretrained CNN + SVM | Scalogram features (CWT) | N/A | N/A | 10-fold CV 10 times | + | 98.6% | - |
| Siuly et al., 2020 [35] | Kaggle  HC: 32  SZ: 49 | Auditory ERP task | DT, KNN, SVM, EBT | Time domain, non-linear features from EMD-EEG | Kruskal-Wallis | + | 10-fold CV | + | 89.59% | - |
| Zhang 2020 [36] | Kaggle  HC: 32  SZ: 49 | Auditory ERP task | ANN | ERP features | N/A | N/A | 70-30 train/test split, 10-fold CV | + | 86.1% | - |
| Azizi et al., 2021 [37] | RepOD HC: 14  SZ: 14 | RS (EC) | Logistic regression | Connectivity features, network thresholding | ANOVA ranking (top 10 features) | + | 10-fold CV | - | 97% (best frequency band) | - |
| Chang et al., 2021 [38] | Private  HC: 40  FSZ: 40  CSZ: 40 | Auditory MMN task | SVM, GCNN | Connectivity features | Significant connections via permutation testing | UNK | 5-fold CV 10 times | - | 93.33% | - |
| Ciprian et al., 2021 [39] | Private  HC: 70  SZ: 62  RepOD  HC: 14  SZ: 14 | RS (EC) | GNB, LDA, KNN, SVM, RF | Connectivity features | ReliefF algorithm with CN-CV | + | 5-fold CN-CV | - | Private  96.92%  RepOD  97.86% | - |
| Dimitriadis, 2021 [40] | MSU  HC: 39  SZ: 45 | RS (EC) | KNN | Spectral, dynamic connectivity, | Infinite feature selection | UNK | 5-fold CV 100 times | - | 100% if features selected | + |
| Guo et al., 2021 [41] | Kaggle  HC: 32  SZ: 49 | Auditory ERP task | CNN | Spectral, information theory, connectivity, ERP features | N/A | N/A | 80-20 train/test split | + | 92% | - |
| Khare and Bajaj, 2021 [42] | Kaggle  HC: 32  SZ: 49 | Auditory ERP task | F-LSSVM | time domain, non-linear features | Fisher score (channel selection), Kruskal-Wallis test (FS) | + | 10-fold CV | + | 91.39% | - |
| Khare et al., 2021 [43] | Kaggle  HC: 32  SZ: 49 | Auditory ERP task | CNN (AlexNet, VGG16, ResNet50) | Spectrogram, scalogram, WVD features | N/A | N/A | 10-fold CV | + | 93.36% | - |
| Khodabakhsh et al., 2021 [44] | MSU  HC: 39  SZ: 45 | RS (EC) | FC-UNET | Connectivity features | N/A | UNK | UNK | UNK | 94.11% | - |
| Kim et al., 2021 [45] | RepOD  HC: 14  SZ: 14 | RS (EC) | SVM, LDA, NB, RF, KNN | Microstate, time domain, spectral, non-linear features | t-test, RFE | - | 10-fold CV | - | 76.85% | - |
| Masychev et al., 2021 [46] | Private  HC: 66  SZ: 57 | Auditory oddball | SVM, RF, LDA, GNB | Source-space connectivity features | MRMR in LOO-CV loop | + | Stratified 10-fold CV | - | 92.68% | - |
| Najafzadeh et al., 2021 [47] | RepOD HC: 14  SZ: 14 | RS (EC) | ANFIS, SVM, ANN | Non-linear, autoregressive features | Top features selected | + | 60-40 train/test split x5 times | - | 98.89% | - |
| Rajesh and Kumar, 2021 [48] | MSU  HC: 39  SZ: 45 | RS (EC) | LogitBoost | SLBP histogram features | Correlation-based feature selection | +/- | 10-fold CV | - | 86.9% (- FS)  91.66% (+ FS) | - |
| Shoeibi et al., 2021 [49] | RepOD  HC: 14  SZ: 14 | RS (EC) | CNN-LSTM | Raw EEG | N/A | N/A | 5-fold CV | + | 99.25% | - |
| Singh et al., 2021 [50] | MSU  HC: 39  SZ: 45 RepOD  HC: 14  SZ: 14 | RS (EC) | CNN-LSTM | UNK | UNK | UNK | UNK | UNK | MSU  94.08%  RepOD  98.96% | - |
| Sun et al., 2021 [51] | Private  HC: 55  SZ: 54 | RS (EO) | CNN-LSTM | Fuzzy entropy, spectral features converted to image | N/A | N/A | 10-fold CV | + | 99.22% | - |
| Aksöz et al., 2022 [52] | Kaggle  HC: 32  SZ: 49 | Auditory ERP task | SVM, KNN, ANN | ERP, time domain, non-linear features | UNK | UNK | 10-fold CV | UNK | 93.90% | - |
| Aslan and Akin, 2022 [53] | MSU  HC: 39  SZ: 45 RepOD  HC: 14  SZ: 14 | RS (EC) | CNN (VGG16) | Scalogram images (CWT) | N/A | N/A | 80-20 train/test split | + | MSU: 98%  RepOD: 99.5% | - |
| Aydemir et al., 2022 [54] | RepOD  HC: 14  SZ: 14 | RS (EC) | CGP17Pat KNN | CGP17Pat features | INCA | + | LOSO-CV, 10-fold CV | +/- | 84.33% (SB)  99.91% (EB) | - |
| Bagherzadeh et al., 2022 [55] | RepOD  HC: 14  SZ: 14 | RS (EC) | CNN-LSTM | Connectivity features | N/A | N/A | 10-fold CV | + | 99.9% | - |
| Barros et al., 2022 [56] | Kaggle  HC: 32  SZ: 49  Private  HC: 31  SZ: 16 | Auditory ERP task | RF, CNN | ERP features, single-trial EEG as image-like features | N/A | N/A | 10-fold CV | - | RF: 73%  CNN: 78% | - |
| Ellis et al., 2022 [57] | Private  HC: 54  SZ: 47 | RS (?) | CNN, CNN-LSTM | Raw EEG | N/A | N/A | 10-fold CV  (8-1-1 train/val/test) | - | 75.9% | - |
| Ilakiyaselvan et al., 2022 [58] | RepOD  HC: 14  SZ: 14 | RS (EC) | CNN (AlexNet, GoogleNet, VGG19, Inception-v4, ResNeXt-50) | Reconstructed phase space features | N/A | N/A | 75-15-10 train/val/test split | + | 99.37% | - |
| Jindal et al., 2022 [59] | RepOD  HC: 14  SZ: 14 | RS (EC) | VGG16 + Bi-LSTM-CNN | MSST-based time-frequency images | N/A | N/A | 70-30 train/test split 10 times | + | 84.42% | - |
| Keihani et al., 2022 [60] | RepOD  HC: 14  SZ: 14 | RS (EC) | Bayesian model selection | Feature ranking + Bayesian model selection | Microstate features | + | 5-fold CV | + | 90.93% | - |
| Khare and Bajaj, 2022 [61] | Kaggle  HC: 32  SZ: 49 | Auditory ERP task | ELM-NN | RVMD + time domain and non-linear features | Hybrid decision support system | + | 10-fold CV | + | 92.93% | - |
| Ko and Yang, 2022 [62] | Kaggle  HC: 32  SZ: 49 | Auditory ERP task | CNN (VGGNet) | Recurrence plot + GAF image features | N/A | N/A | 10-fold CV | + | 90%-92.3% | - |
| Lillo et al., 2022 [63] | RepOD  HC: 14  SZ: 14 | RS (EC) | CNN | Microstates random walk | N/A | N/A | LOO-CV | - | 93% (best) | - |
| Luján et al., 2022 [64] | Private  HC: 320  SZ: 312 | UNK | RBF-NN, SVM, BLDA, GNB, KNN | UNK | Bayesian hyperparameter tuning | UNK | 70-30 CV | UNK | 93.40% (best hyperparameters) | - |
| Nsugbe et al., 2022 [65] | MSU  HC: 39  SZ: 45 | RS (EC) | CNN (AlexNet, SqueezeNet, ResNet-18) LR, DT, SVM | Spectrogram, scalogram, time domain, non-linear features | Only 20 participants included | N/A | 80-20 train/test split | + | 98.3% (best feature fusion model) | - |
| Prabhakar et al., 2022 [66] | RepOD  HC: 14  SZ: 14 | RS (EC) | Bi-LSTM | Fusion hybrid model | Hybrid differential particle bee | + | 10-fold CV  (8-1-1 train/val/test) | + | 98.35% | - |
| Saeedi et al., 2022 [67] | RepOD  HC: 14  SZ: 14 | RS (EC) | CNN-LSTM, CNN-FFT, CNN-CWT | GAF image features |  | N/A | 80-20 train/test split (10x) | + | 99.04% (CNN-FFT) | + |
| Santos Febles et al., 2022 [68] | Private  HC: 54  SZ: 54 | Auditory and visual ERP, MMN | MKL-SVM | ERP, time domain, spectral features | Boruta-RF in nested 10-fold CV | - | LOSO-CV | - | Without FS: 83%  With FS: 86% | - |
| Sharma and Joshi, 2022 [69] | RepOD  HC: 14  SZ: 14  MSU  HC: 39  SZ :45 | RS (EC) | SzHNN (CNN + LSTM), SVM | Band-pass filtered EEG, spectral features (for SVM) | N/A | N/A | 10-fold CV, both subject- and epoch-based | +/- | RepOD  99.9% (EB) 90.11% (SB)  MSU  99.5% (EB)  89.6% (SB) | - |
| Siuly et al., 2022 [70] | Kaggle  HC: 32  SZ: 49 | Auditory ERP task | GoogLeNet with SVM (Schizo-GoogLeNet) | Image-transformed pre-processed EEG | N/A | N/A | 10-fold CV  (7-1-2 train/val/test) | + | 98.84% | - |
| Sobahi et al., 2022 [71] | MSU  HC: 39  SZ: 45 | RS (EC) | ELM, ELM-AE, CNN (ResNet 50, 101) | Image-transformed 1D-LBP features | N/A | N/A | 75-25 train/test split | + | 97.7%  (ResNet-50) | - |
| Supakar et al., 2022 [72] | MSU  HC: 39  SZ: 45 | RS (EC) | RNN-LSTM | Random projection, raw EEG | N/A | N/A | 5-fold CV | + | 98% | - |
| Wang et al., 2022 [73] | Private  HC: 100  SZ: 100  DP: 100  RepOD  HC: 14  SZ: 14 | Private  RS  (EO+EC)  RepOD  RS (EC) | MUCHf-Net (CNN), RF | Spectral features | N/A | N/A | 5-fold CV for validation, separate test set (~15%) for generalization | - | Private:  70.15% (all)  64.87% (SZ  RepOD:  79.3% (all)  75.38% (SZ) | - |
| Wu et al., 2022 [74] | RepOD  HC: 14  SZ: 14 | RS (EC) | Recurrent auto-encoder | Raw EEG | N/A | N/A | LOSO-CV | - | 81.81% | - |
| Agarwal and Singhal, 2023 [75] | RepOD  HC: 14  SZ: 14  Kaggle  HC: 32  SZ: 49 | RepOD  RS (EC)  Kaggle  Auditory ERP task | SVM, KNN, BT, DT | Statistical, time domain, look ahead pattern features | Kruskal-Wallis | + | Holdout CV | UNK | RepOD  98.62%  Kaggle:  99.24% | - |
| Baygin et al., 2023 [76] | Kaggle  HC: 32  SZ: 49 | Auditory ERP task | KNN, CCPNet136 | CCP, ITQWT feature generation | INCA + majority voting | + | 10-fold CV | + | 99.20% | - |
| Bretones et al., 2023 [77] | Private  HC: 320  SZ: 312  SUD+: 128  SUD-: 184 | UNC | GNB, BLDA, SVM, KNN, Adaboost, RBF-ANN | Raw EEG | N/A | N/A | 10-fold CV, with 70-30 train/test split | - | 93.72% | - |
| De Miras et al., 2023 [78] | Private  HC: 20  SZ: 11 | RS (EO) | KNN, LR, DT, RF, SVM | Time domain, non-linear features | t-test, PCA | + | LOSO-CV | - | 89% (SVM, best) | - |
| Divya et al., 2023 [79] | Kaggle  HC: 32  SZ: 49 | Auditory ERP task | DT, LDA, KNN, SVM, ensemble, CNN, CNN-MA | Non-linear features | t-test, May Fly for CNN | + | UNK | UNK | 95.85% | - |
| Göker, 2023 [80] | RepOD  HC: 14  SZ: 14 | RS (EC) | CNN | Periodogram, Welch, Multitaper features | N/A | N/A | 67-33 train/test split | + | 98.75% | - |
| Grover et al., 2023 [81] | RepOD  HC: 14  SZ: 14 | RS (EC) | ANN with late fusion | Connectivity features | N/A | N/A | 70-15-15 train/val/test split, 50 times | + | 99.84% | - |
| Hassan et al., 2023 [82] | RepOD  HC: 14  SZ: 14 | RS (EC) | Hybrid CNN + ML (LR, SVM, RF, GB) | Raw EEG | Channel selection via individual-channel performance | + | 10-fold CV, LOSO-CV (14-fold) | +/- | 98.05% (EB)  90.22% (SB) | - |
| Khare et al., 2023 [83] | MSU  HC: 39  SZ: 45  RepOD  HC: 14  SZ: 14  Kaggle  HC: 32  SZ: 49 | MSU  RS (EC)  RepOD  RS (EC)  Kaggle  Auditory ERP task | CNN (SchizoNET) | Margenau-Hill time-frequency images (channel-wise) | N/A | N/A | 80-20 train/test split, 5-fold CV, 10-fold CV | + | MSU  98.14%  RepOD  99.95%  Kaggle  97.95% | - |
| Kumar et al., 2023 [84] | MSU  HC: 39  SZ: 45  RepOD  HC: 14  SZ: 14 | RS (EC) | AdaBoost | HLV and SLBP feature from 1-minute epochs | Correlation-based feature selection, channel grouping according to brain region | + | LOO-CV (on the level of 1-minute epochs) | +/- | MSU (SB)  92.85%  RepOD (EB)  99.36% | - |
| Li et al., 2023 [85] | RepOD  HC: 14  SZ: 14 | RS (EC) | LeViT | Image-transformed EEG (spatial feature matrix) | N/A | N/A | 10-fold CV, 14-fold, subject-based CV (12-1-1 train/val/test split) | +/- | 98.99% (EB)  85.04% (SB) | - |
| Luján el al., 2023 [86] | Private  HC: 320  SZ: 312 | UNC | ANN-RBF | Raw EEG | N/A | N/A | 70-30 train/test + external validation | UNK | 93.40% | - |
| Parija et al., 2023 [87] | RepOD  HC: 14  SZ: 14  Kaggle  HC: 32  SZ: 49  MSU  HC: 39  SZ: 45 | RepOD  RS (EC)  Kaggle  Auditory ERP task  MSU  RS(EC) | Minimum variance multikernel random vector functional link network | Raw EEG, DSEM-ELM Autoencoder for feature extraction | Unsupervised AE features | UNK | 60-40 train/test split | UNK | RepOD  99.99%  Kaggle  95.01%  MSU  96.65% | - |
| Sahu et al., 2023 [88] | RepOD  HC: 14  SZ: 14  Kaggle  HC: 14  SZ: 14 | RepOD  RS (EC)  Kaggle  Auditory ERP task | CNN | Scalogram (CWT) from overlapping EEG segments | N/A | N/A | 10-fold CV  (8-1-1 train/val/test split) | + | RepOD  99%  Kaggle  96% | - |
| Sharma et al., 2023 [89] | RepOD  HC: 14  SZ: 14 | RS (EC) | CNN-TCN | Raw EEG, statistical, spectral, wavelet, coherence features | N/A | N/A | 10-fold CV both subject-wise and epoch-wise | +/- | 98.89% (EB)  95.89% (SB) | - |
| Shen et al., 2023 [90] | MSU  HC: 39  SZ: 45 | RS (EC) | CNN | Connectivity from CWT scalograms | Various sliding window sizes (2-, 5-, 10-, 30-second) | N/A | 5-fold CV, subject-based groups | - | 97.74% (best) | - |
| Siuly et al., 2023 [91] | Kaggle  HC: 32  SZ: 49 | Auditory ERP task | CNN (ResNet) + SVM | Image-based features | 70-10-20 train/val/test training of Resnet to obtain features | + | 10-fold CV | + | 99.23% | - |
| Aksoy et al., 2024 [92] | MSU  HC: 39  SZ: 45 | RS (EC) | Quantum-SVM, KNN, DT, RF, NB, SVM | Time domain, non-linear features after DWT | Channel selection, PCA | UNK | 70-30 train/test split | UNK | 100% (best classifier + features) | - |
| Garip et al., 2024 [93] | RepOD  HC: 14  SZ: 14 | RS (EC) | DT, RF, ET | Chaotic maps | MPA feature selection | UNK | 10-fold CV | UNK | 97.36% | - |
| Saadatinia and Salimi-Badr, 2024 [94] | MSU  HC: 39  SZ: 45  RepOD  HC: 14  SZ: 14 | RS (EC) | CNN (VGG-16, ResNet-50, MobileNet) | Spectrogram images, data augmentation with GAN | N/A | N/A | 80-20 train/test split | + | MSU  96%  RepOD  99.6% | - |
| Shoeibi et al., 2024 [95] | RepOD  HC: 14  SZ: 14 | RS (EC) | Transformer | Raw EEG | N/A | N/A | 10-fold CV | + | 97.62% | - |

Notes

This selection was based on synthesizing the references collected by [96-99] and [83]. From all collected articles, we excluded [100] as it classified high schizotypy against low schizotypy instead and [101] as it classified schizotypy against normal control (thus, not SZ vs. HC). [102] Was excluded as it considered children with- or without learning disability instead of HC and SZ populations. We also excluded [103, 104] as the full text was not available in English for these papers. Furthermore, we excluded [105] as it does not report SZ vs. HC classification accuracy, and [106] as it only reports AUC but not accuracy. Finally, one reference was incorrectly included/reported in one of the reviews [107] and therefore we did not include it in this sample.

### Assessment of DP- and FS-related leakage

We followed the guidelines recently put forward by Young and colleagues [108] to evaluate if leakage was present or not. This assigned papers into one of three (plus one) categories, separately for DP and FS, as detailed below:

- Leakage (+), denoted as “high risk” in [108]: Clear indication or strong probability of information leakage. For DP, this involved cases where EEG data was augmented by segmentation and the data splitting strategy did not ensure that EEG epochs from the same individual do not end up simultaneously in the training and validation/test sets. For FS, this involved cases where the pipeline indicated that a subset of features was selected before splitting the data into train and validation/test sets and the selection criteria were established using the complete dataset.
- No leakage (-), denoted as “low risk” in [108]: No indication of practices that might result in DP- or FS-related leakage. For DP, this involved cases where it was explicitly stated that data splitting was subject-based, only a single sample was considered per participant, or leave-one-subject-out cross-validation was performed. For FS, this involved cases where it was explicitly stated that feature selection was performed inside the loop to avoid information leakage.
- Unknown (UNK), denoted as “moderate risk” in [108]: The available information was insufficient for definitive judgement. For DP, this involved cases where the evaluation/DP scheme was incompletely or ambiguously described, subject-based DP was not explicitly mentioned, but there was no direct evidence or indication (e.g., confusion matrix) of subject-level information leakage. For FS, this involved cases where the pipeline description was incomplete or ambiguous if features were ranked and selected using data other than the training set, inside-the-loop feature selection was not explicitly mentioned, but there was no direct evidence or indication of improper feature selection.
- Not applicable (N/A): Many pipelines did not employ FS at all (e.g., CNN-based approaches using image-converted EEG or raw EEG) and thus FS-related leakage was not applicable.

Please note that while we use short notation as +/-/UNK, since code implementations were only available for three papers, it is most appropriate to interpret these categories as high/low/moderate risk for information leakage, in line with [108]

### Descriptive statistics

Datasets analyzed (14 studies analyzed 2 or more datasets)

- MSU dataset: 21 studies
- RepOD dataset: 43 studies
- Kaggle dataset: 18 studies
- Private dataset: 28 studies

Data partition

- Subject-based: 34
- Epoch-based: 41
- Both: 7
- Unknown/Unclear: 13

Feature selection

- No feature selection: 46
- Train-only/inside-the-loop: 3
- Full dataset/outside-the-loop: 34
- Both: 2
- Unknown/Unclear: 10

Publications containing both FS- and DP-related leakage: 14 (not including unknown/unclear cases).

Publications containing no FS- or DP-related leakage: 16 (not including unknown/unclear cases).

The total number of publications per calendar year in the considered sample of 95 studies is shown in **Supplementary Figure S1**, which clearly shows a rapidly growing trend (not considering the year 2024, which was only partially covered by the considered systematic reviews [96-99]).

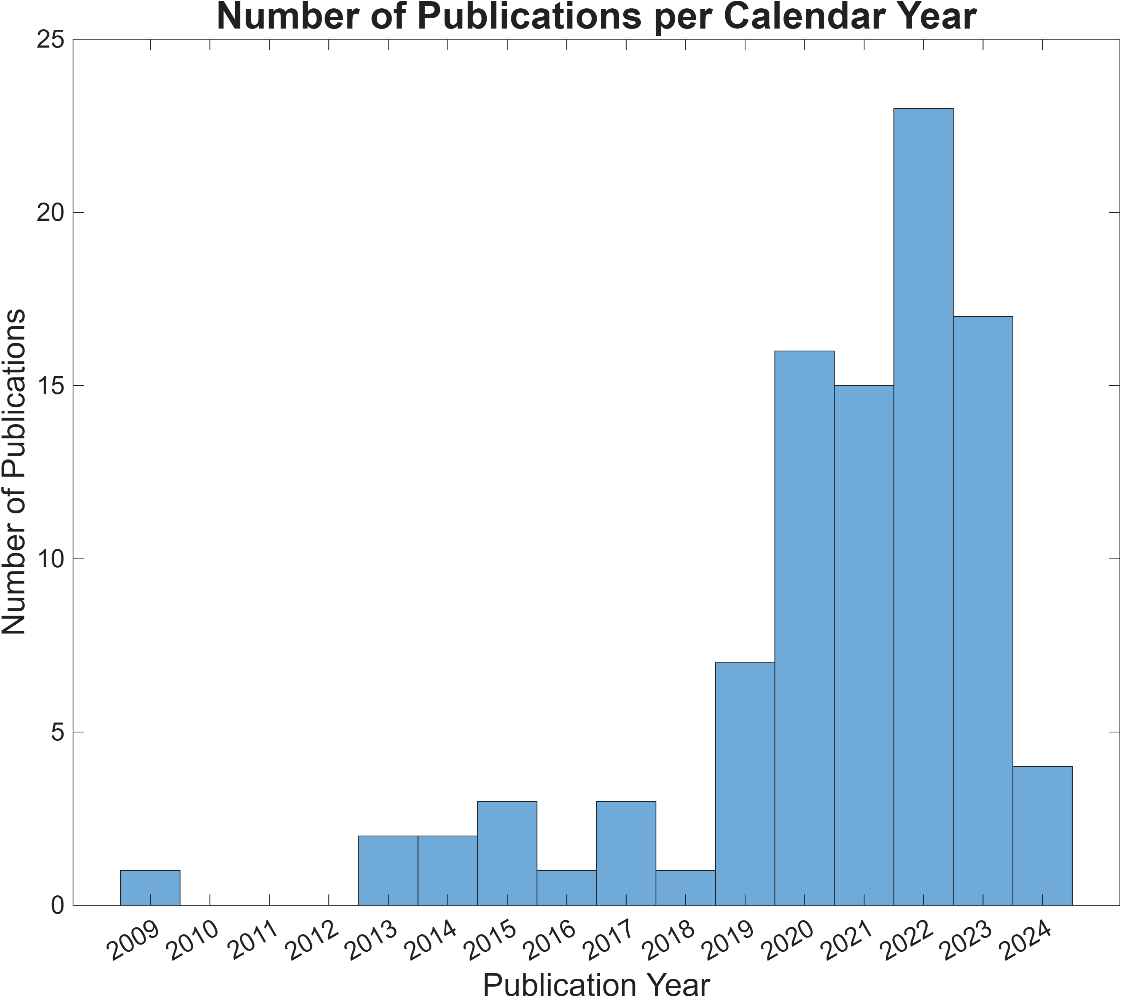

**Supplementary Figure S1.** Number of publications per calendar year in the considered sample of 95 studies.

### Performance differences in subject-based vs. epoch-based pipelines in previous literature

**Supplementary Table S2** collects classification accuracies from six publications that executed classification pipelines both using subject- and epoch-based DP. In these reported classification accuracies, a paired t-test indicates that the average drop of 10.98±4.83% is statistically significant ($t_{6}=6.0107$, $p={9.56}^{-4}$) when switching from epoch-based to subject-based DP. In addition, Kumar et al., 2023 [84] analyzed the RepOD dataset using an epoch-based DP pipeline (99.36%) while the MSU dataset using a subject-based DP pipeline (92.85%), indicating a similar trend.

**Supplementary Table S2.** Table comparing studies evaluating classification performance in subject- and epoch-based data partition (DP) schemes.

| **Paper** | **Dataset** | **Epoch-based DP** | **Subject-based DP** | **Difference** |
| --- | --- | --- | --- | --- |
| Oh et al., 2019 [19] | RepOD | 98.07% | 81.26% | 16.81% |
| Aydemir et al., 2022 [54] | RepOD | 99.91% | 84.33% | 15.58% |
| Sharma and Joshi, 2022 [69] | RepOD  MSU | 99.90%  99.50% | 90.11%  89.6% | 9.79%  9.90% |
| Hassan et al., 2023 [82] | RepOD | 98.05% | 90.22% | 7.83% |
| Li et al., 2023 [85] | RepOD | 98.99% | 85.04% | 13.95% |
| Sharma et al., 2023 [89] | RepOD | 98.89% | 95.89% | 3.00% |

### Feature extraction from EEG data

The extracted features can be sorted into five broader categories: *i*) power spectral features, *ii*) time-frequency features, *iii*) time domain features, *iv*) non-linear features and *v*) connectivity features. We provide the analysis specifics for each of these below.

i) Power spectral features. In every 10-second epoch, a spectrogram was obtained for every channel using the spectrogram() Matlab function. In this, window size was set to 1 second with Hanning-windowing and 75% overlap (0.25-second step size), and the frequency resolution was set to 0.25 Hz. The obtained spectrogram $S(f,t)$ was converted to dB according to $S_{dB}\left( f,t \right)=20\times\log_{10} (\left| S\left( f,t \right) \right|)$. Spectral power was averaged over five canonical frequency bands (delta: 2-4 Hz, theta: 4-7 Hz, alpha: 7-12 Hz, beta: 12-30 Hz and gamma: 30-45 Hz) to obtain time-resolved band-limited power (BLP) estimates. Then, each frequency band was characterized by its mean BLP over time, yielding 5 features per channel (i.e., one per frequency band).
ii) Time-frequency features. These features were intended to capture how spectral properties vary over time. To this end, the same spectrograms were utilized as in *i*), only the standard deviation of BLP was computed for each frequency band, yielding 5 time-frequency features per channel.
iii) Time domain features. Complexity of brain activity was characterized via two distinct methods: Hjorth analysis [109] and fractal time series analysis [110]. Hjorth analysis derives three metrics of EEG dynamics: *Activity* (A), *Mobility* (M) and *Complexity* (C). Namely, given an EEG time series $x_{t}$, Activity is derived as the variance of the signal $A=m_{0}=var(x_{t})$, where $m_{0}$ represents the zeroth spectral moment of $x_{t}$. Mobility and Complexity can be derived from the second ($m_{2}=\frac{dx_{t}}{dt}$) and fourth ($m_{4}=\frac{d^{2}x_{t}}{{dt}^{2}}$) spectral moments such as $M=\sqrt{\frac{m_{2}}{m_{0}}}$ and $C=\sqrt{\frac{m_{4}}{m_{2}}}$. $M$ and $C$ were computed from standardized EEG time series; however, as this would render $m_{0}=1$ in all cases, $A$ was instead replaced by EEG signal variance before standardization. For estimating the fractal scaling exponent, the detrended fluctuation analysis (DFA) method was utilized [111]. In that, the scaling function $S(s)$ was obtained by divided standardized EEG segments into non-overlapping windows of scale $s$, then computing the fluctuation around the local trend in each window and averaging the outcomes [112]. The scaling exponent was then derived via ordinary least squares regression of log-transformed $S\left( s \right)$ over log-transformed $s$. In this analysis, scale was set to correspond to 1/32, 1/16, 1/8, 1/4, 1/2 and 1-second windows. For each epoch these analyses produced 4 features per channel.
iv) Non-linear features. Permutation Entropy [113] was used to characterize the non-linear complexity and information content of EEG signals. Permutation Entropy utilizes the time-delay embedding technique put forward by Takens [114] by converting EEG data into state space vectors using embedding dimension $m$ and time delay $L$. Given signals with explicit frequency band priors, $m$ and $L$ can be set according to simple signal processing principles [115]. The utilized values for $m$ and $L$ are provided in **Supplementary Table S3**. Then, in each state space vector, amplitude values are replaced by their respective ordinal ranks within the vector, converting it into one of the $m!$ possible permutations of order $m$. Once the signal is thus converted into a sequence of symbols (permutations $\pi$) – where the relative frequency $p(\pi)$ of a given symbol $\pi$ can be easily estimated –, Permutation Entropy of order $m$ is obtained via $PE\left( m \right)=-\sum p\left( \pi\right)\log p(\pi)$ where the sum runs over all $m!$ possible permutations [113]. Furthermore, since the maximum possible value of $PE\left( m \right)$ is known to be $\log m!$, it can be normalized to the $[0, 1]$ range where 0 and 1 denote perfectly regular and white noise processes. As $PE(m)$ was computed on signals filtered in five frequency bands, this analysis provided 5 features per channel, per epoch.
v) Connectivity features. In this study, the Phase-Lag Index (PLI) was utilized as functional connectivity estimator [116]. First, in each case an adjacency matrix was obtained by computing PLI between all possible channel pairs. Elements of the main diagonal was set to zero, excluding self-connections. Since weak connectivity estimates likely represent spurious connections, we employed a connection density-based thresholding scheme to eliminate these [117]. In that, given a threshold density $D$, only the strongest connections are kept while the rest are set to zero so that the connection density in the resulting network is equal to $D$. This procedure was carried out using threshold densities ranging from 15% to 50% in 5% increments. Finally, following the approach in [33], networks thresholded at different values of $D$ were averaged, and each channel (i.e., node of network) was characterized by its node degree i.e., the sum of all connection weights of that given node. This procedure was carried out in all five frequency bands, yielding 5 features per channel, per epoch.

**Supplementary Table S3.** Parameter settings for Permutation Entropy for different frequency bands.

| **Dataset** | | **Delta (2-4 Hz)** | **Theta (4-7 Hz)** | **Alpha (7-12 Hz)** | **Beta (12-30 Hz)** | **Gamma (30-45 Hz)** |
| --- | --- | --- | --- | --- | --- | --- |
| MSU (128 Hz) | $m$ | 7 | 7 | 7 | 9 | 6 |
|  | $L$ | 11 | 7 | 4 | 2 | 1 |
| RepOD and SU-SZ  (250 Hz) | $m$ | 7 | 7 | 7 | 9 | 6 |
|  | $L$ | 21 | 12 | 7 | 3 | 2 |

### Ablation study using ensemble-based classifiers: the effect of leaky feature selection

Classification performances for the three datasets considered in our study (MSU [118], RepOD [119] and SU-SZ [120]) under various leakage scenarios are presented in **Supplementary Table S4**. Since sample sizes were substantially larger than the number of extracted features due to data augmentation (segmentation), we employed a less radical feature selection scheme, in which either 50% of features were retained, or those with a positive weight assigned to by the ReliefF algorithm [121], whichever was smaller. Notably, in this setup we observed no significant effect of leaky FS (i.e., Leaky DP+FS vs. Leaky DP, or Leaky FS vs. Leakage-free, $p>0.05$ in all cases via permutation testing).

**Supplementary Table S4.** Classification performances using boosted ensemble classifier and feature selection under various leakage scenarios. DP: data partition; FS: feature selection.

| **Dataset** | **Leaky DP+FS** | **Leaky DP** | **Leaky FS** | **Leakage-free** |
| --- | --- | --- | --- | --- |
| MSU | 95.62 ± 4.69% | 95.43 ± 4.31% | 82.75 ± 10.08% | 82.27 ± 7.91% |
| RepOD | 97.62 ± 1.14% | 97.47 ± 1.07% | 68.40 ± 12.24% | 64.73 ± 13.58% |
| SU-SZ | 94.68 ± 3.54% | 94.42 ± 3.24% | 64.80 ± 8.55% | 64.91 ± 8.96% |

### Performance Summary

**Supplementary Table S5.** Performance summary through eight standard metrics. Values represent pooled performance over cross-validation runs. bACC: balanced accuracy; MCC: Matthew’s correlation coefficient; ROC-AUC receiver operator characteristic area under the curve.

| **Model** | **Dataset** | **Leakage** | **bACC (%)** | **Specificity (%)** | **Sensitivity (%)** | **Precision (%)** | **F1 score (%)** | **MCC** | **Cohen’s** $\boldsymbol{\kappa}$ | **ROC-AUC** |
| --- | --- | --- | --- | --- | --- | --- | --- | --- | --- | --- |
| CNN | `MSU | Leaky | 85.03 | 80.98 | 89.07 | 84.39 | 86.67 | 0.8524 | 0.8518 | 0.9344 |
|  |  | Leakage-free | 81.00 | 77.56 | 84.44 | 81.28 | 82.83 | 0.8112 | 0.8110 | 0.9002 |
|  | RepOD | Leaky | 86.33 | 84.92 | 87.75 | 85.34 | 86.52 | 0.8635 | 0.8633 | 0.9388 |
|  |  | Leakage-free | 58.06 | 53.27 | 62.85 | 57.36 | 59.98 | 0.5810 | 0.5806 | 0.6099 |
|  | SU-SZ | Leaky | 80.47 | 79.62 | 81.32 | 79.54 | 80.42 | 0.8046 | 0.8046 | 0.8915 |
|  |  | Leakage-free | 67.59 | 68.46 | 66.71 | 67.33 | 67.02 | 0.6759 | 0.6759 | 0.7543 |
| Ensemble | MSU | Leaky | 95.44 | 92.74 | 98.15 | 93.97 | 96.01 | 0.9565 | 0.9560 | 0.9950 |
|  |  | Leakage-free | 81.88 | 78.21 | 85.56 | 81.91 | 83.70 | 0.8203 | 0.8199 | 0.8962 |
|  | RepOD | Leaky | 97.62 | 97.52 | 97.72 | 97.52 | 97.62 | 0.9762 | 0.9762 | 0.9977 |
|  |  | Leakage-free | 64.73 | 57.74 | 71.73 | 62.92 | 67.04 | 0.6488 | 0.6473 | 0.7142 |
|  | SU-SZ | Leaky | 94.68 | 94.62 | 94.74 | 94.49 | 94.61 | 0.9467 | 0.9467 | 0.9881 |
|  |  | Leakage-free | 65.10 | 72.31 | 57.89 | 67.07 | 62.15 | 0.6527 | 0.6513 | 0.7300 |
| SVM | MSU | Leaky | 90.94 | 86.32 | 95.56 | 87.48 | 91.34 | 0.9112 | 0.9094 | 0.9571 |
|  |  | Leakage-free | 80.77 | 73.62 | 87.92 | 76.92 | 82.05 | 0.8109 | 0.8077 | 0.8724 |
|  | RepOD | Leaky | 63.78 | 58.67 | 68.88 | 62.50 | 65.53 | 0.6385 | 0.6378 | 0.7148 |
|  |  | Leakage-free | 56.38 | 54.59 | 58.16 | 56.16 | 57.14 | 0.5638 | 0.5638 | 0.5843 |
|  | SU-SZ | Leaky | 81.44 | 76.05 | 86.84 | 78.38 | 82.39 | 0.8163 | 0.8144 | 0.8464 |
|  |  | Leakage-free | 72.84 | 69.03 | 76.65 | 71.22 | 73.84 | 0.7291 | 0.7284 | 0.7914 |
| EEG foundation model | MSU | Leaky | 96.40 | 95.94 | 96.85 | 96.49 | 96.67 | 0.9641 | 0.9641 | 0.9964 |
|  |  | Leakage-free | 83.20 | 79.91 | 86.48 | 83.24 | 84.83 | 0.8333 | 0.8330 | 0.9136 |
|  | RepOD | Leaky | 98.26 | 98.45 | 98.07 | 98.47 | 98.27 | 0.9826 | 0.9826 | 0.9979 |
|  |  | Leakage-free | 72.91 | 58.84 | 86.99 | 68.18 | 76.45 | 0.7390 | 0.7296 | 0.7855 |
|  | SU-SZ | Leaky | 88.81 | 87.98 | 89.64 | 87.98 | 88.80 | 0.8881 | 0.8880 | 0.9531 |
|  |  | Leakage-free | 59.20 | 58.78 | 59.62 | 58.68 | 59.15 | 0.5920 | 0.5920 | 0.6323 |

To further illustrate performance in the various evaluation pipelines, confusion matrices are presented below on **Supplementary Figures** **S2**-**S5**. Note that these matrices aggregate outcomes from all CV runs for a given evaluation pipeline.

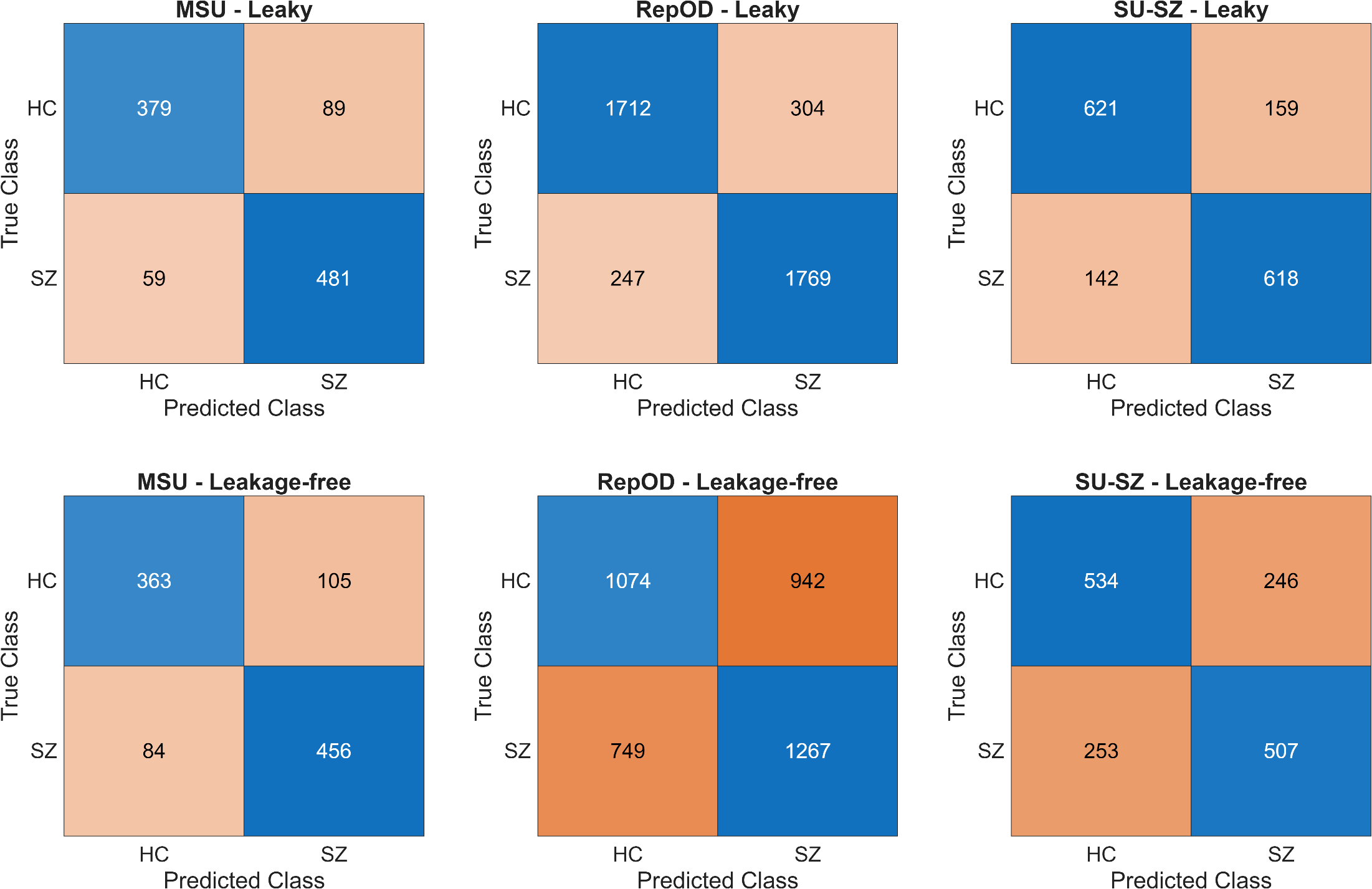

**Supplementary Figure S2.** Aggregated confusion matrices from leaky (upper row) and leakage-free (lower row) evaluations of the CNN-based pipeline in the three datasets.

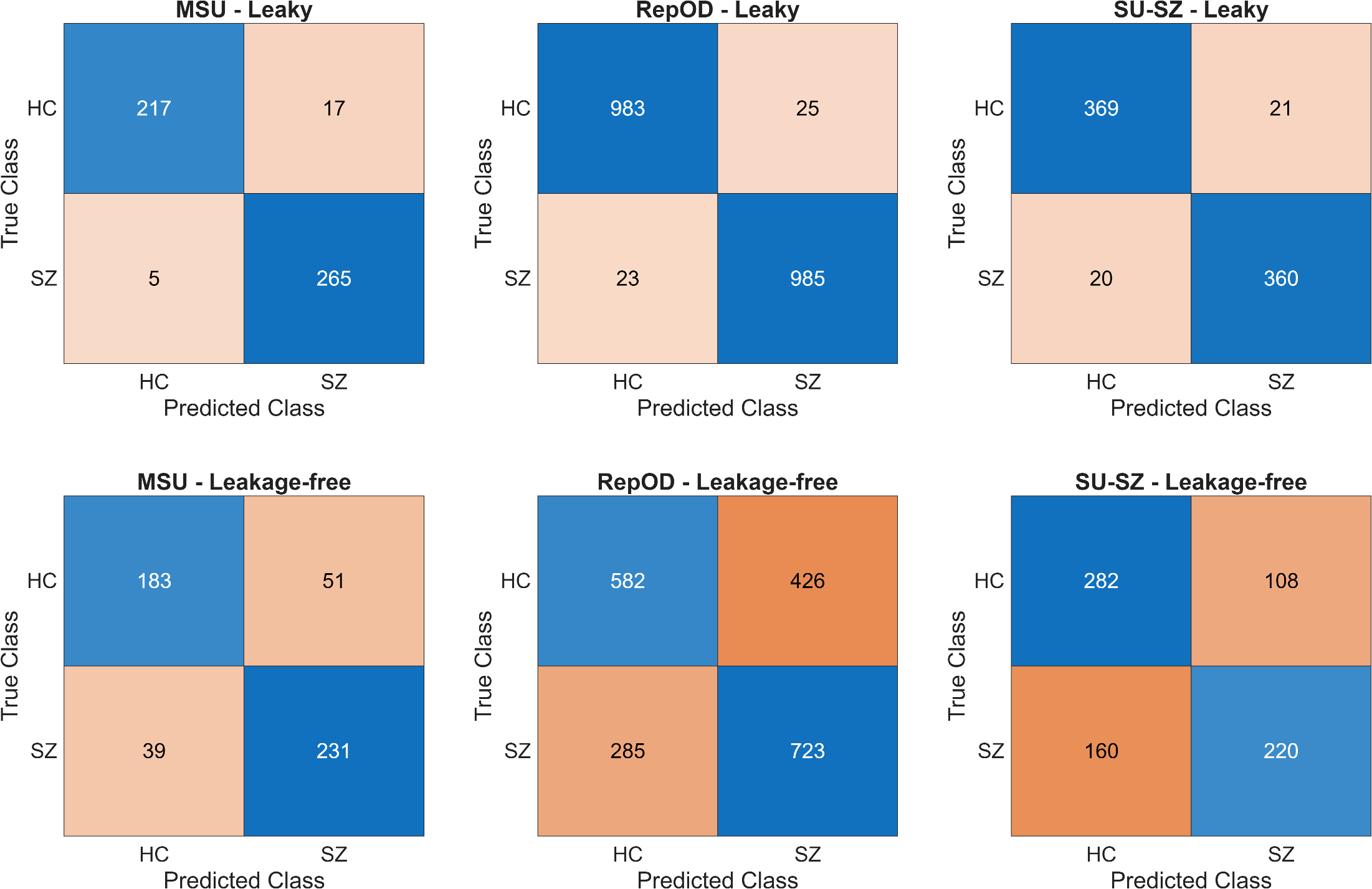

**Supplementary Figure S3.** Aggregated confusion matrices from leaky (upper row) and leakage-free (lower row) evaluations of the ensemble-based pipeline in the three datasets.

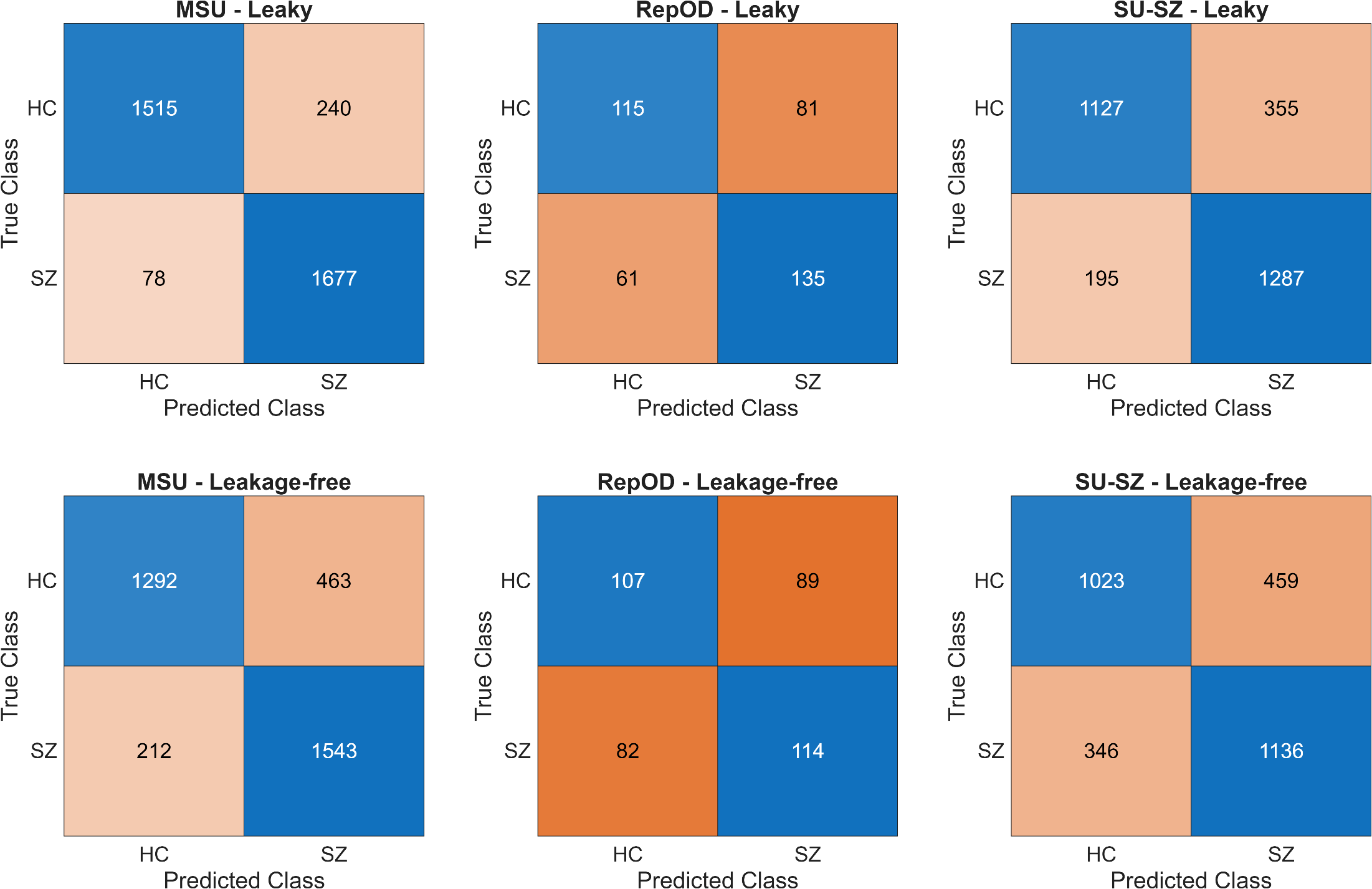

**Supplementary Figure S4.** Aggregated confusion matrices from leaky (upper row) and leakage-free (lower row) evaluations of the subject-based SVM pipeline in the three datasets.

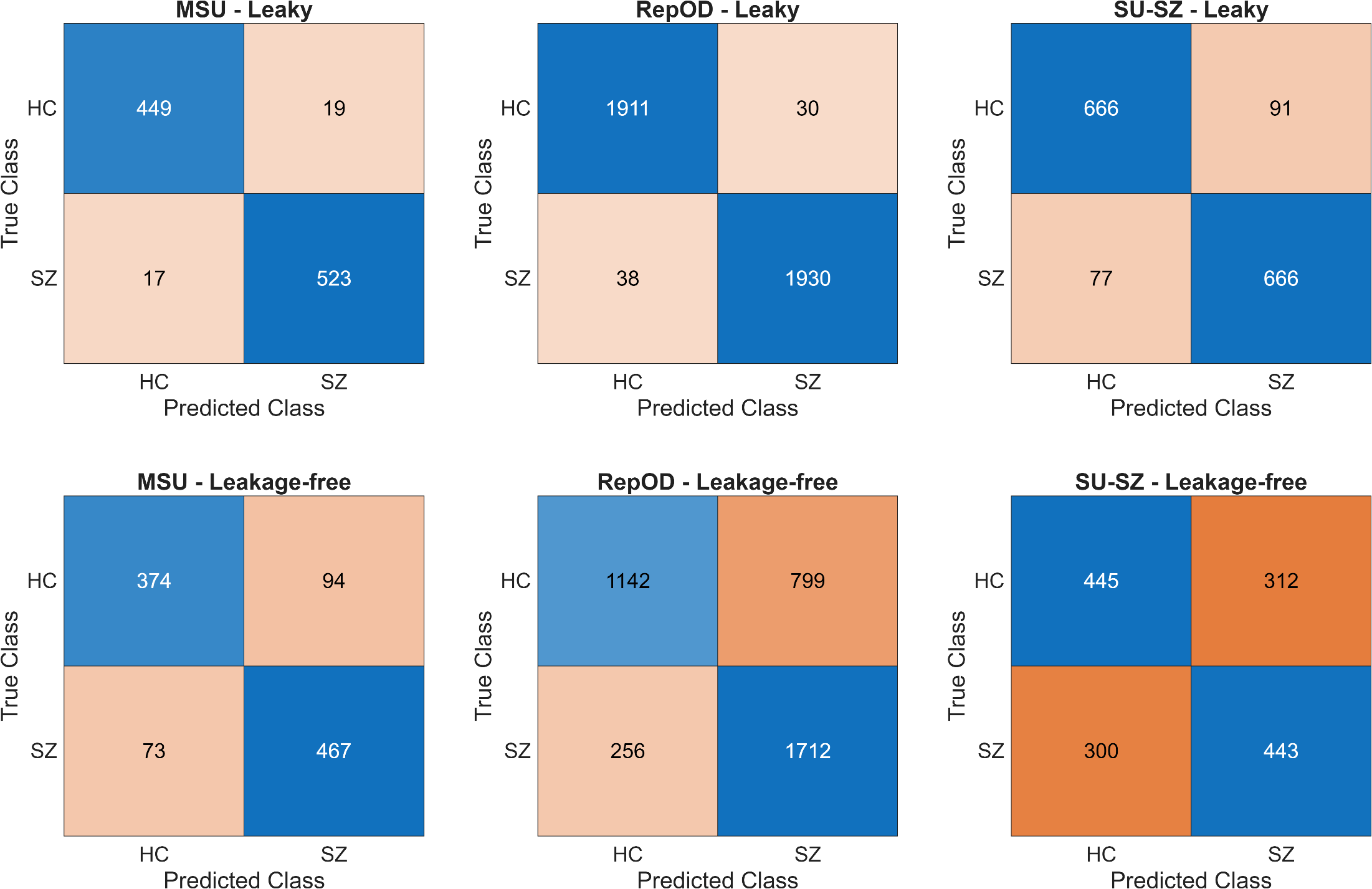

**Supplementary Figure S5.** Aggregated confusion matrices from leaky (upper row) and leakage-free (lower row) evaluations of the EEG foundation model pipeline in the three datasets.

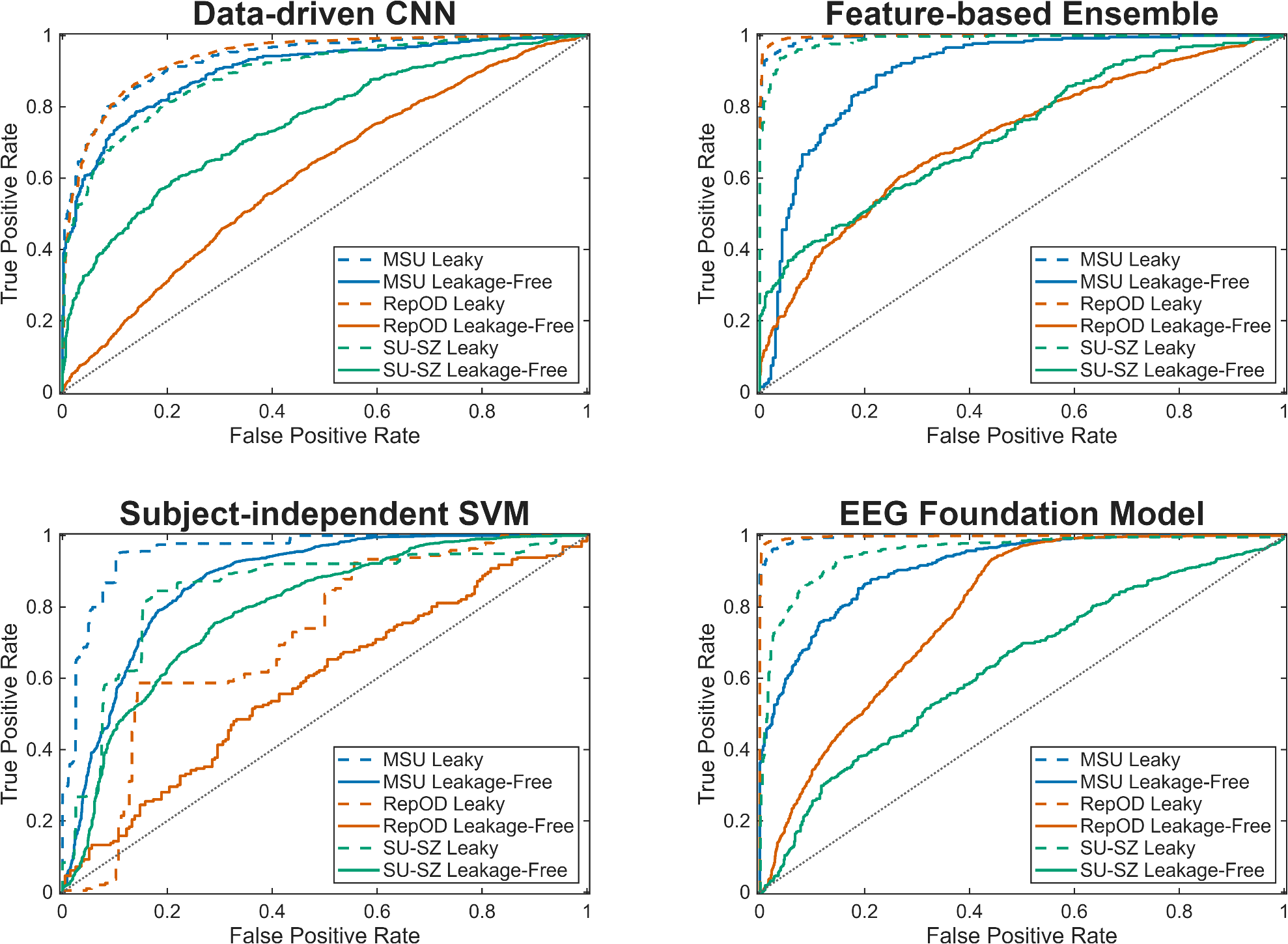

**Supplementary Figure S6.** Receiver operator characteristic (ROC) curves for each evaluation scenario. In each case, the ROC curve was derived after aggregating outcomes from all cross-validation iterations.

### Scoping Review

We used the following search term: "schizophrenia" AND ("EEG" OR "Electroencephalography") AND ("machine learning" OR "deep learning") AND ("classification" OR "detection" OR "diagnosis"). This search term was utilized to collect articles from the following databases: EBSCO-PsycInfo, Embase, IEEE-Xplore, PubMed and Web of Science Core Collection. A total of 362 references were collected with this process. We utilized the Rayyan tool [122] to synthesize and screen these references. Removing duplicates resulted in a final number of 219 papers; the title and abstract of these were screened for consideration. We employed the following screening criteria:

Inclusion criteria:

- **Inclusion Criteria 1:** Population: Studies performing binary classification of patients with schizophrenia against healthy controls using EEG data. Patients in the SZ group must have a diagnosis according to ICD-10 or DSM-5 (schizophrenia including all subtypes).
- **Inclusion Criteria 2:** Concept: ML or DL models/approaches that are evaluated in a standard framework (e.g., training + testing, cross-validation, etc.) and report standard performance measures.
- **Inclusion Criteria 3:** Context: Peer-reviewed journal articles (original research articles), conference proceedings, preprints. All included studies must report original data analysis and outcomes.
- **Inclusion Criteria 4:** Time Frame: Publication/acceptance date from 01 Jan 2024 until 01 Nov 2025.
- **Inclusion Criteria 5:** Language: English.

Exclusion criteria:

- **Exclusion Criteria 1:** Studies not involving a SZ patient population and a healthy control population
- **Exclusion Criteria 2:** Studies not using EEG, or studies using additional modalities to EEG (e.g., fMRI)
- **Exclusion Criteria 3:** Studies not reporting standard performance metrics.
- **Exclusion Criteria 4:** Studies only utilizing conventional statistical methods for hypothesis testing, but no ML/DL for prediction.
- **Exclusion Criteria 5:** Reviews, opinion pieces, letters, abstracts only.
- **Exclusion Criteria 6:** Non-English publications.

Screening according to these criteria yielded 125 publications for full-text screening. Since performing a full scoping or systematic review is beyond the scope of our current study, we did not proceed with full-text screening and assessing the presence of various leakage scenarios in these articles. Nevertheless, this preliminary screening analysis indicates that the number of relevant publications grows rapidly and such an updated synthesis of the literature is called for.

### References

1.	Sabeti, M., S. Katebi, and R. Boostani, *Entropy and complexity measures for EEG signal classification of schizophrenic and control participants.* Artificial intelligence in medicine, 2009. **47**(3): p. 263-274.

2.	Hiesh, M.-H., et al. *Classification of schizophrenia using genetic algorithm-support vector machine (ga-svm)*. in *2013 35th Annual International Conference of the IEEE Engineering in Medicine and Biology Society (EMBC)*. 2013. IEEE.

3.	Neuhaus, A.H., et al., *Single-subject classification of schizophrenia using event-related potentials obtained during auditory and visual oddball paradigms.* European archives of psychiatry and clinical neuroscience, 2013. **263**(3): p. 241-247.

4.	Parvinnia, E., et al., *Classification of EEG Signals using adaptive weighted distance nearest neighbor algorithm.* Journal of King Saud University-Computer and Information Sciences, 2014. **26**(1): p. 1-6.

5.	Sui, J., et al. *Combination of FMRI-SMRI-EEG data improves discrimination of schizophrenia patients by ensemble feature selection*. in *2014 36th Annual International Conference of the IEEE Engineering in Medicine and Biology Society*. 2014. IEEE.

6.	Dvey-Aharon, Z., et al., *Schizophrenia detection and classification by advanced analysis of EEG recordings using a single electrode approach.* PloS one, 2015. **10**(4): p. e0123033.

7.	Ravan, M., et al., *A machine learning approach using auditory odd-ball responses to investigate the effect of Clozapine therapy.* Clinical Neurophysiology, 2015. **126**(4): p. 721-730.

8.	Santos-Mayo, L., L.M. San-José-Revuelta, and J.I. Arribas, *A computer-aided diagnosis system with EEG based on the P3b wave during an auditory odd-ball task in schizophrenia.* IEEE transactions on biomedical engineering, 2016. **64**(2): p. 395-407.

9.	Shim, M., et al., *Machine-learning-based diagnosis of schizophrenia using combined sensor-level and source-level EEG features.* Schizophrenia research, 2016. **176**(2-3): p. 314-319.

10.	Chu, L., et al., *Individual recognition in schizophrenia using deep learning methods with random forest and voting classifiers: Insights from resting state EEG streams.* arXiv preprint arXiv:1707.03467, 2017.

11.	Liu, H., et al., *A data driven approach for resting-state EEG signal classification of schizophrenia with control participants using random matrix theory.* arXiv preprint arXiv:1712.05289, 2017.

12.	Piryatinska, A., B. Darkhovsky, and A. Kaplan, *Binary classification of multichannel-EEG records based on the ϵ-complexity of continuous vector functions.* Computer methods and programs in biomedicine, 2017. **152**: p. 131-139.

13.	Alimardani, F., et al., *Classification of bipolar disorder and schizophrenia using steady-state visual evoked potential based features.* IEEE Access, 2018. **6**: p. 40379-40388.

14.	Buettner, R., et al., *High-performance exclusion of schizophrenia using a novel machine learning method on EEG data*. 2019, Bogotá, Colombia.

15.	Devia, C., et al., *EEG classification during scene free-viewing for schizophrenia detection.* IEEE Transactions on Neural Systems and Rehabilitation Engineering, 2019. **27**(6): p. 1193-1199.

16.	Jahmunah, V., et al., *Automated detection of schizophrenia using nonlinear signal processing methods.* Artificial Intelligence in Medicine, 2019. **100**.

17.	Li, F., et al., *Differentiation of schizophrenia by combining the spatial EEG brain network patterns of rest and task P300.* IEEE transactions on neural systems and rehabilitation engineering, 2019. **27**(4): p. 594-602.

18.	Naira, C.A.T. and C. Jos, *Classification of people who suffer schizophrenia and healthy people by EEG signals using deep learning.* International Journal of Advanced Computer Science and Applications, 2019. **10**(10).

19.	Oh, S.L., et al., *Deep Convolutional Neural Network Model for Automated Diagnosis of Schizophrenia Using EEG Signals.* Applied Sciences-Basel, 2019. **9**(14).

20.	Phang, C.-R., et al. *Classification of EEG-based effective brain connectivity in schizophrenia using deep neural networks*. in *2019 9th International IEEE/EMBS Conference on Neural Engineering (NER)*. 2019. IEEE.

21.	Ahmedt-Aristizabal, D., et al., *Identification of children at risk of schizophrenia via deep learning and EEG responses.* IEEE journal of biomedical and health informatics, 2020. **25**(1): p. 69-76.

22.	Aslan, Z. and M. Akin, *Automatic Detection of Schizophrenia by Applying Deep Learning over Spectrogram Images of EEG Signals.* Traitement du Signal, 2020. **37**(2).

23.	Buettner, R., et al., *Development of a machine learning based algorithm to accurately detect schizophrenia based on one-minute EEG recordings.* HICSS-53 Proceedings, IEEE, 2020.

24.	Calhas, D., E. Romero, and R. Henriques, *On the use of pairwise distance learning for brain signal classification with limited observations.* Artificial intelligence in medicine, 2020. **105**: p. 101852.

25.	Goshvarpour, A. and A. Goshvarpour, *Schizophrenia diagnosis using innovative EEG feature-level fusion schemes.* Physical and Engineering Sciences in Medicine, 2020. **43**(1): p. 227-238.

26.	Khare, S.K., et al., *Classification of schizophrenia patients through empirical wavelet transformation using electroencephalogram signals*, in *Modelling and analysis of active biopotential signals in healthcare, Volume 1*. 2020, IOP Publishing Bristol, UK. p. 1-1-1-26.

27.	Kim, J.-Y., H.S. Lee, and S.-H. Lee, *EEG source network for the diagnosis of schizophrenia and the identification of subtypes based on symptom severity—A machine learning approach.* Journal of Clinical Medicine, 2020. **9**(12): p. 3934.

28.	Krishnan, P.T., et al., *Schizophrenia detection using MultivariateEmpirical Mode Decomposition and entropy measures from multichannel EEG signal.* Biocybernetics and Biomedical Engineering, 2020. **40**(3): p. 1124-1139.

29.	Nikhil Chandran, A., K. Sreekumar, and D. Subha, *EEG-based automated detection of schizophrenia using long short-term memory (LSTM) network*, in *Advances in Machine Learning and Computational Intelligence: Proceedings of ICMLCI 2019*. 2020, Springer. p. 229-236.

30.	Phang, C.R., et al., *A Multi-Domain Connectome Convolutional Neural Network for Identifying Schizophrenia From EEG Connectivity Patterns.* Ieee Journal of Biomedical and Health Informatics, 2020. **24**(5): p. 1333-1343.

31.	Prabhakar, S.K., H. Rajaguru, and S.-W. Lee, *A framework for schizophrenia EEG signal classification with nature inspired optimization algorithms.* IEEE Access, 2020. **8**: p. 39875-39897.

32.	Prabhakar, S.K., H. Rajaguru, and S.-H. Kim, *Schizophrenia EEG signal classification based on swarm intelligence computing.* Computational Intelligence and Neuroscience, 2020. **2020**(1): p. 8853835.

33.	Racz, F.S., et al., *Multifractal and Entropy-Based Analysis of Delta Band Neural Activity Reveals Altered Functional Connectivity Dynamics in Schizophrenia.* Frontiers in Systems Neuroscience, 2020. **14**.

34.	Shalbaf, A., S. Bagherzadeh, and A. Maghsoudi, *Transfer learning with deep convolutional neural network for automated detection of schizophrenia from EEG signals.* Physical and Engineering Sciences in Medicine, 2020. **43**(4): p. 1229-1239.

35.	Siuly, S., et al., *A computerized method for automatic detection of schizophrenia using EEG signals.* IEEE Transactions on Neural Systems and Rehabilitation Engineering, 2020. **28**(11): p. 2390-2400.

36.	Zhang, L. *EEG signals feature extraction and artificial neural networks classification for the diagnosis of schizophrenia*. in *2020 IEEE 19th International Conference on Cognitive Informatics & Cognitive Computing (ICCI* CC)*. 2020. IEEE.

37.	Azizi, S., D.B. Hier, and D.C. Wunsch. *Schizophrenia classification using resting state EEG functional connectivity: source level outperforms sensor level*. in *2021 43rd Annual International Conference of the IEEE Engineering in Medicine & Biology Society (EMBC)*. 2021. IEEE.

38.	Chang, Q., et al., *Classification of first-episode schizophrenia, chronic schizophrenia and healthy control based on brain network of mismatch negativity by graph neural network.* IEEE Transactions on Neural Systems and Rehabilitation Engineering, 2021. **29**: p. 1784-1794.

39.	Ciprian, C., et al., *Diagnosing schizophrenia using effective connectivity of resting-state EEG data.* Algorithms, 2021. **14**(5): p. 139.

40.	Dimitriadis, S.I., *Reconfiguration of αmplitude driven dominant coupling modes (DoCM) mediated by α-band in adolescents with schizophrenia spectrum disorders.* Progress in Neuro-Psychopharmacology and Biological Psychiatry, 2021. **108**: p. 110073.

41.	Guo, Z., et al. *Deep neural network classification of EEG data in schizophrenia*. in *2021 IEEE 10th Data Driven Control and Learning Systems Conference (DDCLS)*. 2021. IEEE.

42.	Khare, S.K. and V. Bajaj, *A self-learned decomposition and classification model for schizophrenia diagnosis.* Computer Methods and Programs in Biomedicine, 2021. **211**: p. 106450.

43.	Khare, S.K., V. Bajaj, and U.R. Acharya, *SPWVD-CNN for automated detection of schizophrenia patients using EEG signals.* IEEE Transactions on Instrumentation and Measurement, 2021. **70**: p. 1-9.

44.	Khodabakhsh, A., H. Arabi, and H. Zaidi. *U-net based estimation of functional connectivity from time series multi-channel eeg from schizophrenia patients*. in *2021 IEEE Nuclear Science Symposium and Medical Imaging Conference (NSS/MIC)*. 2021. IEEE.

45.	Kim, K., et al., *EEG microstate features for schizophrenia classification.* PloS one, 2021. **16**(5): p. e0251842.

46.	Masychev, K., et al., *Advanced signal processing methods for characterization of schizophrenia.* IEEE Transactions on Biomedical Engineering, 2021. **68**(4): p. 1123-1130.

47.	Najafzadeh, H., et al., *Automatic classification of schizophrenia patients using resting-state EEG signals.* Physical and Engineering Sciences in Medicine, 2021. **44**(3): p. 855-870.

48.	Rajesh, K.N. and T.S. Kumar. *Schizophrenia detection in adolescents from EEG signals using symmetrically weighted local binary patterns*. in *2021 43rd Annual International Conference of the IEEE Engineering in Medicine & Biology Society (EMBC)*. 2021. IEEE.

49.	Shoeibi, A., et al., *Automatic diagnosis of schizophrenia in EEG signals using CNN-LSTM models.* Frontiers in neuroinformatics, 2021. **15**: p. 777977.

50.	Singh, K., S. Singh, and J. Malhotra, *Spectral features based convolutional neural network for accurate and prompt identification of schizophrenic patients.* Proceedings of the Institution of Mechanical Engineers, Part H: Journal of Engineering in Medicine, 2021. **235**(2): p. 167-184.

51.	Sun, J., et al., *A hybrid deep neural network for classification of schizophrenia using EEG Data.* Scientific Reports, 2021. **11**(1): p. 4706.

52.	Aksöz, A., et al., *Analysis and classification of schizophrenia using event related potential signals.* Computer Science, 2022: p. 32-36.

53.	Aslan, Z. and M. Akin, *A deep learning approach in automated detection of schizophrenia using scalogram images of EEG signals.* Physical and Engineering Sciences in Medicine, 2022. **45**(1): p. 83-96.

54.	Aydemir, E., et al. *CGP17Pat: automated schizophrenia detection based on a cyclic group of prime order patterns using EEG signals*. in *Healthcare*. 2022. MDPI.

55.	Bagherzadeh, S., M.S. Shahabi, and A. Shalbaf, *Detection of schizophrenia using hybrid of deep learning and brain effective connectivity image from electroencephalogram signal.* Computers in Biology and Medicine, 2022. **146**: p. 105570.

56.	Barros, C., et al., *From sound perception to automatic detection of schizophrenia: an EEG-based deep learning approach.* Frontiers in Psychiatry, 2022. **12**: p. 813460.

57.	Ellis, C.A., et al. *Examining effects of schizophrenia on EEG with explainable deep learning models*. in *2022 IEEE 22nd International Conference on Bioinformatics and Bioengineering (BIBE)*. 2022. IEEE.

58.	Ilakiyaselvan, N., A.N. Khan, and A. Shahina, *Reconstructed phase space portraits for detecting brain diseases using deep learning.* Biomedical Signal Processing and Control, 2022. **71**: p. 103278.

59.	Jindal, K., et al., *Bi-LSTM-deep CNN for schizophrenia detection using MSST-spectral images of EEG signals*, in *Artificial Intelligence-Based Brain-Computer Interface*. 2022, Elsevier. p. 145-162.

60.	Keihani, A., et al., *Bayesian optimization of machine learning classification of resting-state EEG microstates in schizophrenia: a proof-of-concept preliminary study based on secondary analysis.* Brain sciences, 2022. **12**(11): p. 1497.

61.	Khare, S.K. and V. Bajaj, *A hybrid decision support system for automatic detection of Schizophrenia using EEG signals.* Computers in biology and medicine, 2022. **141**: p. 105028.

62.	Ko, D.-W. and J.-J. Yang, *EEG-Based schizophrenia diagnosis through time series image conversion and deep learning.* Electronics, 2022. **11**(14): p. 2265.

63.	Lillo, E., M. Mora, and B. Lucero, *Automated diagnosis of schizophrenia using EEG microstates and Deep Convolutional Neural Network.* Expert Systems with Applications, 2022. **209**: p. 118236.

64.	Luján, M.Á., et al., *Mental disorder diagnosis from EEG signals employing automated leaning procedures based on radial basis functions.* Journal of Medical and Biological Engineering, 2022. **42**(6): p. 853-859.

65.	Nsugbe, E., et al. *Intelligence combiner: a combination of deep learning and handcrafted features for an adolescent psychosis prediction using EEG signals*. in *2022 IEEE International Workshop on Metrology for Industry 4.0 & IoT (MetroInd4. 0&IoT)*. 2022. IEEE.

66.	Prabhakar, S.K., et al., *A fusion-based technique with hybrid swarm algorithm and deep learning for biosignal classification.* Frontiers in Human Neuroscience, 2022. **16**: p. 895761.

67.	Saeedi, M., A. Saeedi, and P. Mohammadi, *Schizophrenia diagnosis via FFT and wavelet convolutional neural networks utilizing EEG signals.* 2022.

68.	Santos Febles, E., et al., *Machine learning techniques for the diagnosis of schizophrenia based on event-related potentials.* Frontiers in Neuroinformatics, 2022. **16**: p. 893788.

69.	Sharma, G. and A.M. Joshi, *SzHNN: A novel and scalable deep convolution hybrid neural network framework for schizophrenia detection using multichannel EEG.* IEEE Transactions on Instrumentation and Measurement, 2022. **71**: p. 1-9.

70.	Siuly, S., et al., *SchizoGoogLeNet: The GoogLeNet‐Based Deep Feature Extraction Design for Automatic Detection of Schizophrenia.* Computational Intelligence and Neuroscience, 2022. **2022**(1): p. 1992596.

71.	Sobahi, N., et al., *A new signal to image mapping procedure and convolutional neural networks for efficient schizophrenia detection in EEG recordings.* IEEE Sensors Journal, 2022. **22**(8): p. 7913-7919.

72.	Supakar, R., P. Satvaya, and P. Chakrabarti, *A deep learning based model using RNN-LSTM for the detection of schizophrenia from EEG data.* Computers in Biology and Medicine, 2022. **151**: p. 106225.

73.	Wang, Z., et al., *Automated rest EEG-based diagnosis of depression and schizophrenia using a deep convolutional neural network.* IEEE Access, 2022. **10**: p. 104472-104485.

74.	Wu, Y., et al. *Schizophrenia detection based on EEG using recurrent auto-encoder framework*. in *International Conference on Neural Information Processing*. 2022. Springer.

75.	Agarwal, M. and A. Singhal, *Fusion of pattern-based and statistical features for Schizophrenia detection from EEG signals.* Medical Engineering & Physics, 2023. **112**: p. 103949.

76.	Baygin, M., et al., *CCPNet136: automated detection of schizophrenia using carbon chain pattern and iterative TQWT technique with EEG signals.* Physiological Measurement, 2023. **44**(3): p. 035008.

77.	Bretones, C.S., et al., *Automatic identification of schizophrenia employing EEG records analyzed with deep learning algorithms.* Schizophrenia Research, 2023. **261**: p. 36-46.

78.	De Miras, J.R., et al., *Schizophrenia classification using machine learning on resting state EEG signal.* Biomedical Signal Processing and Control, 2023. **79**: p. 104233.

79.	Divya, V., et al., *Signal Conducting System with Effective Optimization Using Deep Learning for Schizophrenia Classification.* Comput. Syst. Sci. Eng., 2023. **45**(2): p. 1869-1886.

80.	Göker, H., *1D-convolutional neural network approach and feature extraction methods for automatic detection of schizophrenia.* Signal, Image and Video Processing, 2023. **17**(5): p. 2627-2636.

81.	Grover, N., et al., *Schizo-Net: A novel Schizophrenia Diagnosis framework using late fusion multimodal deep learning on Electroencephalogram-based Brain connectivity indices.* IEEE Transactions on Neural Systems and Rehabilitation Engineering, 2023. **31**: p. 464-473.

82.	Hassan, F., S.F. Hussain, and S.M. Qaisar, *Fusion of multivariate EEG signals for schizophrenia detection using CNN and machine learning techniques.* Information Fusion, 2023. **92**: p. 466-478.

83.	Khare, S.K., V. Bajaj, and U.R. Acharya, *SchizoNET: a robust and accurate Margenau–Hill time-frequency distribution based deep neural network model for schizophrenia detection using EEG signals.* Physiological Measurement, 2023. **44**(3): p. 035005.

84.	Kumar, T.S., et al., *Automated Schizophrenia detection using local descriptors with EEG signals.* Engineering Applications of Artificial Intelligence, 2023. **117**: p. 105602.

85.	Li, B., et al., *Automatic detection of schizophrenia based on spatial–temporal feature mapping and LeViT with EEG signals.* Expert Systems with Applications, 2023. **224**: p. 119969.

86.	Luján, M.Á., et al., *Accurate neural network classification model for schizophrenia disease based on electroencephalogram data.* International Journal of Machine Learning and Cybernetics, 2023. **14**(3): p. 861-872.

87.	Parija, S., et al., *Autoencoder-based improved deep learning approach for schizophrenic EEG signal classification.* Pattern Analysis and Applications, 2023. **26**(2): p. 403-435.

88.	Sahu, G., et al., *SCZ-SCAN: An automated Schizophrenia detection system from electroencephalogram signals.* Biomedical Signal Processing and Control, 2023. **86**: p. 105206.

89.	Sharma, G., et al., *A smart healthcare framework for accurate detection of schizophrenia using multichannel EEG.* IEEE Transactions on Instrumentation and Measurement, 2023. **72**: p. 1-9.

90.	Shen, M., et al., *Automatic identification of schizophrenia based on EEG signals using dynamic functional connectivity analysis and 3D convolutional neural network.* Comput Biol Med, 2023. **160**: p. 107022.

91.	Siuly, S., et al., *Exploring deep residual network based features for automatic schizophrenia detection from EEG.* Physical and Engineering Sciences in Medicine, 2023. **46**(2): p. 561-574.

92.	Aksoy, G., et al., *Quantum machine-based decision support system for the detection of schizophrenia from EEG records.* Journal of medical systems, 2024. **48**(1): p. 29.

93.	Garip, Z., et al., *Chaotic marine predator optimization algorithm for feature selection in schizophrenia classification using EEG signals.* Cluster Computing, 2024. **27**(8): p. 11277-11297.

94.	Saadatinia, M. and A. Salimi-Badr, *An explainable deep learning-based method for schizophrenia diagnosis using generative data-augmentation.* IEEE Access, 2024.

95.	Shoeibi, A., et al. *Early diagnosis of schizophrenia in EEG signals using one dimensional transformer model*. in *International work-conference on the interplay between natural and artificial computation*. 2024. Springer.

96.	Ranjan, R., B.C. Sahana, and A.K. Bhandari, *Deep learning models for diagnosis of schizophrenia using EEG signals: emerging trends, challenges, and prospects.* Archives of Computational Methods in Engineering, 2024. **31**(4): p. 2345-2384.

97.	Saha, A., et al., *Schizophrenia detection and classification: A systematic review of the last decade.* Diagnostics, 2024. **14**(23): p. 2698.

98.	Rahul, J., et al., *A systematic review of EEG based automated schizophrenia classification through machine learning and deep learning.* Frontiers in Human Neuroscience, 2024. **18**: p. 1347082.

99.	Uyanik, H., et al., *Automated Detection of Neurological and Mental Health Disorders Using EEG Signals and Artificial Intelligence: A Systematic Review.* Wiley Interdisciplinary Reviews: Data Mining and Knowledge Discovery, 2025. **15**(1): p. e70002.

100.	Zandbagleh, A., et al., *Classification of low and high schizotypy levels via evaluation of brain connectivity.* International Journal of Neural Systems, 2022. **32**(04): p. 2250013.

101.	Jeong, J.W., et al., *Classifying schizotypy using an audiovisual emotion perception test and scalp electroencephalography.* Frontiers in human neuroscience, 2017. **11**: p. 450.

102.	Seshadri, N.G., et al., *EEG based classification of children with learning disabilities using shallow and deep neural network.* Biomedical Signal Processing and Control, 2023. **82**: p. 104553.

103.	Du, X., et al., *Research on electroencephalogram specifics in patients with schizophrenia under cognitive load.* Sheng wu yi xue Gong Cheng xue za zhi= Journal of Biomedical Engineering= Shengwu Yixue Gongchengxue Zazhi, 2020. **37**(1): p. 45-53.

104.	Lai, H., et al., *Resting-state electroencephalogram classification of patients with schizophrenia or depression.* Sheng wu yi xue Gong Cheng xue za zhi= Journal of Biomedical Engineering= Shengwu Yixue Gongchengxue Zazhi, 2019. **36**(6): p. 916-923.

105.	Guo, G., et al., *Method for persistent topological features extraction of schizophrenia patients’ electroencephalography signal based on persistent homology.* Frontiers in Computational Neuroscience, 2022. **16**: p. 1024205.

106.	Vázquez, M.A., A. Maghsoudi, and I.P. Mariño, *An interpretable machine learning method for the detection of schizophrenia using EEG signals.* Frontiers in Systems Neuroscience, 2021. **15**: p. 652662.

107.	Bao, X., et al., *A novel ultrasound robot with force/torque measurement and control for safe and efficient scanning.* IEEE transactions on instrumentation and measurement, 2023. **72**: p. 1-12.

108.	Young, V.M., et al., *Data Leakage in Deep Learning for Alzheimer’s Disease Diagnosis: A Scoping Review of Methodological Rigor and Performance Inflation.* Diagnostics, 2025. **15**(18): p. 2348.

109.	Hjorth, B., *EEG analysis based on time domain properties.* Electroencephalography and clinical neurophysiology, 1970. **29**(3): p. 306-310.

110.	Eke, A., et al., *Fractal characterization of complexity in temporal physiological signals.* Physiological measurement, 2002. **23**(1): p. 1-38.

111.	Peng, C.K., et al., *Mosaic Organization of DNA Nucleotides.* Physical Review E, 1994. **49**(2): p. 1685-1689.

112.	Kaposzta, Z., et al., *Real-Time Algorithm for Detrended Cross-Correlation Analysis of Long-Range Coupled Processes.* Frontiers in Physiology, 2022. **13**.

113.	Bandt, C. and B. Pompe, *Permutation entropy: A natural complexity measure for time series.* Physical Review Letters, 2002. **88**(17).

114.	Takens, F., *Detecting strange attractors in turbulence*, in *Dynamical systems and turbulence, Warwick 1980*, D. Rand and L.-S. Young, Editors. 1981, Springer: Berlin. p. 366-381.

115.	Montez, T., et al., *Synchronization likelihood with explicit time-frequency priors.* Neuroimage, 2006. **33**(4): p. 1117-1125.

116.	Stam, C.J., G. Nolte, and A. Daffertshofer, *Phase lag index: Assessment of functional connectivity from multi channel EEG and MEG with diminished bias from common sources.* Human Brain Mapping, 2007. **28**(11): p. 1178-1193.

117.	Rubinov, M. and O. Sporns, *Complex network measures of brain connectivity: Uses and interpretations.* NeuroImage, 2010. **52**(3): p. 1059-1069.

118.	Borisov, S., et al., *Analysis of EEG structural synchrony in adolescents with schizophrenic disorders.* Human Physiology, 2005. **31**(3): p. 255-261.

119.	Olejarczyk, E. and W. Jernajczyk, *Graph-based analysis of brain connectivity in schizophrenia.* PLoS One, 2017. **12**(11): p. e0188629.

120.	Racz, F.S., et al., *Reduced temporal variability of cortical excitation/inhibition ratio in schizophrenia.* Schizophrenia (Heidelb), 2025. **11**(1): p. 20.

121.	Urbanowicz, R.J., et al., *Relief-based feature selection: Introduction and review.* Journal of biomedical informatics, 2018. **85**: p. 189-203.

122.	Ouzzani, M., et al., *Rayyan—a web and mobile app for systematic reviews.* Systematic reviews, 2016. **5**(1): p. 210.
