## Supplementary Document 1 for "Information Leakage and Performance Overestimation in EEG-Based Schizophrenia Detection: Evidence from Literature and Empirical Analyses"

**Checklist for Transparent Reporting of Machine Learning Pipelines in Neuroimaging-based Computer-Aided Diagnostics (ML-NICAD Checklist)**

| **Manuscript checklist** | |
| --- | --- |
| 1. **Title and Abstract** | |
| 1.1. Does the title clearly indicate the use of ML/DL for CAD, the data modality and the target condition? |  |
| 1.2. Does the Abstract summarize the dataset, pipeline overview, evaluation scheme and key results? |  |
| 2. **Introduction/Background** | |
| 2.1. Is the clinical problem and rationale for ML-CAD, including comparison with standard-of-care clinical practices clearly outlined? |  |
| 2.2. Are potential biases/caveats considered? |  |
| 3. **Dataset Description (extend for multiple datasets if applicable)** | |
| 3.1. Dataset ID (if applicable) |  |
| 3.1.1. Dataset availability | Private / public |
| 3.1.1.1. If public: citation/link |  |
| 3.1.2. Sample characteristics |  |
| 3.1.2.1. Sample groups |  |
| 3.1.2.2. Total N subjects (per group) |  |
| 3.1.2.3. Age (mean/SD/range per group) |  |
| 3.1.2.4. Sex (M/F per group) |  |
| 3.1.2.5. Demographics other – please extend (e.g., ethnicity, diagnosis criteria, years in education, medication status, symptom severity, etc.) |  |
| 3.1.2.5.1. … |  |
| 3.1.3. Data Structure |  |
| 3.1.3.1. Modality |  |
| 3.1.3.2. Sampling rate |  |
| 3.1.3.3. Number of simultaneously obtained streams |  |
| 3.1.3.4. Recording length (per subject) |  |
| 3.1.3.5. Condition(s) |  |
| 3.1.3.6. If data augmentation: is multi-sample per subject data explicitly described? | Yes / no / not applicable |
| 4. **Results and Reporting** | |
| 4.1. Main outcomes |  |
| 4.2. Secondary outcomes |  |
| 4.3. Limitations (if applicable) |  |
| 4.4. Benchmarking |  |
| 4.4.1. Computational |  |
| 4.4.2. Clinical (if applicable) |  |
| 5. **Discussion and Interpretation** | |
| 5.1. Clinical relevance (if applicable) |  |
| 5.2. Broader implications |  |
| 6. **Reproducibility and Other Information** | |
| 6.1. Data availability |  |
| 6.2. Code availability |  |
| 6.3. Disclosures |  |
| 6.4. Checklist compliance/self-assessment |  |

| **Evaluation checklist** | |
| --- | --- |
| 1. **Model Specifics** | |
| 1.1. Model ID |  |
| 1.2. Date of execution |  |
| 1.3. Hardware environment |  |
| 1.4. Software environment |  |
| 1.5. Execution time |  |
| 1.6. Dataset ID |  |
| 1.7. Study groups/response variable |  |
| 1.8. Number of subjects (per group/condition) |  |
| 2. **Data Pre-processing** | |
| 2.1. Steps in order – please extend |  |
| 2.1.1. … |  |
| 2.2. Software/tools |  |
| 2.3. Handling of missing/imbalanced data |  |
| 2.3.1. Method |  |
| 2.3.2. Before/after partitioning? |  |
| 2.4. Data augmentation |  |
| 2.4.1. Method |  |
| 2.4.2. Justification |  |
| 2.5. Output format (per subject) |  |
| 3. **Feature Extraction/Engineering** | |
| 3.1. Feature details – please extend (feature type, unit of FE, number of features per unit, etc.) |  |
| 3.1.1. … |  |
| 3.2. Dimensionality reduction | Yes / no / not applicable |
| 3.2.1. Method |  |
| 3.2.2. Parameters |  |
| 3.2.3. Timing (i.e., before/after data partitioning) |  |
| 3.3. Normalization |  |
| 3.3.1. Method |  |
| 3.3.2. Parameters |  |
| 3.3.3. Timing |  |
| 3.4. Aggregation (if applicable) |  |
| 3.5. Output format (per subject) |  |
| 4. **Model Architecture and Training** | |
| 4.1. Model type |  |
| 4.2. Implementation |  |
| 4.3. Input format |  |
| 4.4. Key parameters |  |
| 4.5. Training procedure |  |
| 4.6. Normalization + timing (before/after data partitioning) |  |
| 5. **Hyperparameter Tuning (if applicable, otherwise skip to 8.)** | |
| 5.1. Method |  |
| 5.2. Parameters tuned |  |
| 5.3. Timing/scheme |  |
| 5.4. Leakage confirmation |  |
| 6. **Data Partitioning (DP) and Cross-Validation (CV, if applicable)** | |
| 6.1. Full dataset size |  |
| 6.2. Scheme and ratio |  |
| 6.3. Hierarchical handling (i.e., group-based CV) |  |
| 6.4. Stratification |  |
| 6.5. Repetitions |  |
| 6.6. For multi-dataset: cross-dataset/pooled-dataset validation? |  |
| 6.7. Leakage confirmation |  |
| 7. **Feature Selection** | |
| 7.1. Method |  |
| 7.2. Parameters |  |
| 7.3. Timing |  |
| 7.4. Leakage confirmation |  |
| 8. **Evaluation** | |
| 8.1. Metrics |  |
| 8.1.1. Primary |  |
| 8.1.2. Secondary |  |
| 8.2. Aggregation (if applicable) |  |
| 8.3. Statistical testing |  |
| 8.3.1. Chance-level |  |
| 8.3.2. Comparative |  |
| 8.4. Generalization (external data) |  |
| 8.5. Leakage confirmation |  |

**Notes:**

We acknowledge that the proposed checklist might not accommodate all classification/regression pipelines or strategies; extend with additional information wherever necessary. The current version is proposed primarily for classification scenarios.
