## Supplementary Document 2 for "Information Leakage and Performance Overestimation in EEG-Based Schizophrenia Detection: Evidence from Literature and Empirical Analyses"

**Checklist for Transparent Reporting of Machine Learning Pipelines in Neuroimaging-based Computer-Aided Diagnostics (ML-NICAD Checklist)**

### General Information

| **Manuscript checklist** | |
| --- | --- |
| 1. **Title and Abstract** | |
| 1.1. Does the title clearly indicate the use of ML/DL for CAD, the data modality and the target condition? | Yes |
| 1.2. Does the Abstract summarize the dataset, pipeline overview, evaluation scheme and key results? | Yes (minimal summary) |
| 2. **Introduction/Background** | |
| 2.1. Is the clinical problem and rationale for ML-CAD, including comparison with standard-of-care clinical practices clearly outlined? | Yes |
| 2.2. Are potential biases/caveats considered? | Yes |
| 3. **Dataset Description (extend for multiple datasets if applicable)** | |
| 3.1. Dataset 1 | MSU dataset |
| 3.1.1. Dataset availability | Private / **public** |
| 3.1.1.1. If public: citation/link | <http://brain.bio.msu.ru/eeg_schizophrenia.htm> |
| 3.1.2. Sample characteristics |  |
| 3.1.2.1. Sample groups | Healthy Control (HC), Schizophrenia (SZ) |
| 3.1.2.2. Total N subjects (per group) | HC: 39  SZ: 45 |
| 3.1.2.3. Age (mean/SD/range per group) | HC: 12.25 [11-13.75]  SZ: 12.25 [10.67-14] |
| 3.1.2.4. Sex (M/F per group) | N/A |
| 3.1.2.5. Demographics other – please extend (e.g., ethnicity, diagnosis criteria, years in education, medication status, symptom severity, etc.) | N/A |
| 3.1.3. Data Structure |  |
| 3.1.3.1. Modality | EEG |
| 3.1.3.2. Sampling rate | 128 Hz |
| 3.1.3.3. Number of simultaneously obtained streams | 16 channels |
| 3.1.3.4. Recording length (per subject) | 1 minute |
| 3.1.3.5. Condition(s) | Resting-state (eyes-closed) |
| 3.1.3.6. If data augmentation: is multi-sample per subject data explicitly described? | **Yes** / no / not applicable |
| 3.2. Dataset 2 | RepOD dataset |
| 3.2.1. Dataset availability | Private / **public** |
| 3.2.1.1. If public: citation/link | <https://doi.org/10.18150/repod.0107441> |
| 3.2.2. Sample characteristics |  |
| 3.2.2.1. Sample groups | Healthy Control (HC), Schizophrenia (SZ) |
| 3.2.2.2. Total N subjects (per group) | HC: 14  SZ: 14 |
| 3.2.2.3. Age (mean/SD/range per group) | HC: 27.75 (3.16)  SZ: 28.10 (3.72) |
| 3.2.2.4. Sex (M/F per group) | HC: 7F/7M  SZ: 7F/7M |
| 3.2.2.5. Demographics other – please extend (e.g., ethnicity, diagnosis criteria, years in education, medication status, symptom severity, etc.) | Medication washout for at least 2 years prior to assessment |
| 3.2.3. Data Structure |  |
| 3.2.3.1. Modality | EEG |
| 3.2.3.2. Sampling rate | 250 Hz |
| 3.2.3.3. Number of simultaneously obtained streams | 19 channels |
| 3.2.3.4. Recording length (per subject) | >12 minutes |
| 3.2.3.5. Condition(s) | Resting-state (eyes-closed) |
| 3.2.3.6. If data augmentation: is multi-sample per subject data explicitly described? | **Yes** / no / not applicable |
| 3.3. Dataset 3 | SU-SZ dataset |
| 3.3.1. Dataset availability | Private / **public** |
| 3.3.1.1. If public: citation/link | <https://doi.org/10.5281/zenodo.14808295> |
| 3.3.2. Sample characteristics |  |
| 3.3.2.1. Sample groups | Healthy Control (HC), Schizophrenia (SZ) |
| 3.3.2.2. Total N subjects (per group) | HC: 39  SZ: 38 |
| 3.3.2.3. Age (mean/SD/range per group) | HC: 32.8 (9.5)  SZ: 34.3 (10.7) |
| 3.3.2.4. Sex (M/F per group) | HC: 15F/24M  SZ: 17F/21M |
| 3.3.2.5. Demographics other – please extend (e.g., ethnicity, diagnosis criteria, years in education, medication status, symptom severity, etc.) | Patients were ON medication at the time of recording. Further information (medication, years in education, disease duration) available at data repository. |
| 3.3.3. Data Structure |  |
| 3.3.3.1. Modality | EEG |
| 3.3.3.2. Sampling rate | 1000 Hz |
| 3.3.3.3. Number of simultaneously obtained streams | 64 channels |
| 3.3.3.4. Recording length (per subject) | >100 seconds |
| 3.3.3.5. Condition(s) | Resting-state (eyes-closed) |
| 3.3.3.6. If data augmentation: is multi-sample per subject data explicitly described? | **Yes** / no / not applicable |
| 4. **Results and Reporting** | |
| 4.1. Main outcomes | Classification accuracy |
| 4.2. Secondary outcomes | Balanced accuracy, sensitivity, specificity, precision, F1-score, MCC, Cohen’s Kappa, ROC-AUC |
| 4.3. Limitations (if applicable) | Small sample size, no hyperparameter tuning, no cross-dataset validation |
| 4.4. Benchmarking |  |
| 4.4.1. Computational | Not considered |
| 4.4.2. Clinical (if applicable) | Not considered |
| 5. **Discussion and Interpretation** | |
| 5.1. Clinical relevance (if applicable) | No direct clinical utility considered, assessment of realistic expected performance in CADs for schizophrenia detection |
| 5.2. Broader implications | Recommending best practices, potential generalizability to other medical conditions (e.g., major depressive disorder, bipolar disorder) |
| 6. **Reproducibility and Other Information** | |
| 6.1. Data availability | Included |
| 6.2. Code availability | https://github.com/samuelracz/ schizophrenia_EEG_detection_leakage |
| 6.3. Disclosures | No conflicts of interest, no funding to declare |
| 6.4. Checklist compliance/self-assessment | All relevant fields completed except: 4.4 |

### Pipeline 1: fine-tuned ResNet18 with image-transformed EEG features

| **Evaluation checklist** | |
| --- | --- |
| 1. **Model Specifics** | |
| 1.1. Model IDs | MSU-CNN-Leaky, MSU-CNN-Subj  RepOD-CNN-Leaky, RepOD-CNN-Subj  SU-SZ-CNN-Leaky, RepOD-CNN-Subj |
| 1.2. Date of execution | MSU: 11/21/2025  RepOD: 11/21/2025  SU-SZ: 11/21/2025 |
| 1.3. Hardware environment | 12^th^ Gen Intel® CoreTM i9-12900Kx24  NVIDIA GeForce RTX 4090  Physical memory: 64 Gb |
| 1.4. Software environment | MATLAB R2025a |
| 1.5. Execution time | MSU: ~10min  RepOD: ~18min  SU-SZ: ~14min |
| 1.6. Dataset IDs | MSU  RepOD  SU-SZ |
| 1.7. Study groups/response variable | MSU: HC vs. SZ  RepOD: HC vs. SZ  SU-SZ: HC vs. SZ |
| 1.8. Number of subjects (per group/condition) | MSU: HC=39, SZ=45  RepOD: HC=14, SZ=14  SU-SZ: HC=39, SZ=38 |
| 2. **Data Pre-processing** | |
| 2.1. Steps in order – please extend |  |
| 2.1.1. Channel selection – **only for SU-SZ** | Out of 64 channels, only the 19 corresponding to standard 10-20 are retained |
| 2.1.2. Band-pass filtering | 1-45 Hz |
| 2.1.3. Notch filtering | 50 Hz (CleanLine) |
| 2.1.4. Downsampling – **only for SU-SZ** | 1000 Hz data downsampled to 250 Hz |
| 2.1.5. ICA-based artifact removal (automated) | Multiple Artifact Rejection Algorithm, default parameters |
| 2.1.6. Re-referencing | Common average reference |
| 2.1.7. Normalization | Standardization (z-scoring) per channel |
| 2.2. Software/tools | Matlab 2025a, EEGLAB |
| 2.3. Handling of missing/imbalanced data |  |
| 2.3.1. Method | N/A |
| 2.3.2. Before/after partitioning? | N/A |
| 2.4. Data augmentation | **Yes** / no / not applicable |
| 2.4.1. Method | Segmentation into 5-second non-overlapping epochs |
| 2.4.2. Justification | Increase sample size |
| 2.5. Output format (per subject) | MSU: 640x16x12  RepOD: 1250x19x144  SU-SZ: 1250x19x20  (*time* *points* x *channels* x *epochs*) |
| 3. **Feature Extraction/Engineering** | |
| 3.1. Feature details – please extend (feature type, unit of FE, number of features per unit, etc.) |  |
| 3.1.1. Time-frequency decomposition for each channel | Three different time-frequency decomposition schemes (see below) |
| 3.1.1.1. Spectrogram map | Matlab spectrogram()  Window=1*fs  noverlap=0.9*fs  f=1:0.25:45  fs=128 or 250  transformed to dB |
| 3.1.1.2. Continuous wavelet transform | Matlab cwt()  wname=’morse’  1-45 Hz regime isolated, log-transformed |
| 3.1.1.3. Smoothed pseudo Wigner-Ville Distribution | Matlab wvd()  “smoothedPseudo”  1-45 Hz regime isolated, log-transformed |
| 3.1.2. Data aggregation | Channel-wise maps organized into grid according to scalp localization to form a single image per TF decomposition technique. Missing locations filled with average of surrounding electrodes. |
| 3.1.2.1. Spectrogram output format (per epoch) | MSU: 708x200  RepOD/SU-SZ: 885x205 |
| 3.1.2.2. CWT output format (per epoch) | MSU: 216x3200  RepOD/SU-SZ: 275x6250 |
| 3.1.2.3. WVD output format (per epoch) | MSU: 1760x6400  RepOD/SU-SZ: 2200x12500 |
| 3.1.3. Scaling values | Each TF matrix (spectrogram, CWT, WVD) is scaled between [0, 255] for ResNet18 input |
| 3.1.4. Reshaping | Matlab imresize()  Each TF matrix (spectrogram, CWT, WVD) is reshaped to 224x224 for ResNet18 input |
| 3.1.5. Image construction (1 image per epoch) | The three TF maps are concatenated to form a tri-color image of size 224x224x3 |
| 3.2. Dimensionality reduction | Yes / **no** / not applicable |
| 3.2.1. Method | N/A |
| 3.2.2. Parameters | N/A |
| 3.2.3. Timing (i.e., before/after data partitioning) | N/A |
| 3.3. Normalization |  |
| 3.3.1. Method | Min-max scaling the three time-frequency maps to [0, 1] in each image |
| 3.3.2. Parameters | N/A |
| 3.3.3. Timing | Independently for each image |
| 3.4. Aggregation (if applicable) | none |
| 3.5. Output format (per subject) | MSU: (224x224x3)x12  RepOD: (224x224x3)x144  SU-SZ: (224x224x3)x20 |
| 4. **Model Architecture and Training** | |
| 4.1. Model type | CNN (ResNet18) |
| 4.2. Implementation | Matlab imagePretrainedNetwork()  First 10 convolutional layers frozen. An additional DropOut layer with p=0.4 is introduced before the fully connected layer. Binary prediction obtained via SoftMax layer. |
| 4.3. Input format | 224x224x3 |
| 4.4. Key parameters | solverName=’adam’  InitialLearnRate=1e-5  MaxEpochs=25  ValidationPatience=3  L2Regularization=0.005  Shuffle=’every-epoch’  GradientThresholdMethod=’l2norm’  GradientThreshold=1  MiniBatchSize=64  ValidationFrequency=30  Verbose=false  ExecutionEnvironment=’GPU’ |
| 4.5. Training procedure | Matlab trainNetwork() |
| 4.6. Normalization + timing (before/after data partitioning) | Images already normalized before imputation |
| 5. **Hyperparameter Tuning (if applicable, otherwise skip to 6.)** | |
| 5.1. Method | N/A |
| 5.2. Parameters tuned | N/A |
| 5.3. Timing/scheme | N/A |
| 5.4. Leakage confirmation | N/A |
| 6. **Data Partitioning (DP) and Cross-Validation (CV, if applicable)** | |
| 6.1. Full dataset size | MSU: (224x224x3)x1008  RepOD: (224x224x3)x4032  SU-SZ: (224x224x3)x1540 |
| 6.2. Scheme and ratio | **Leaky pipelines**: stratified 10-fold CV  (7-fold CV for RepOD)  **Subj pipelines:** subject-based stratified 10-fold CV (7-fold CV for RepOD) |
| 6.3. Hierarchical handling (i.e., group-based CV) | Subject-based data partitioning in **Subj** **pipelines** |
| 6.4. Stratification | Stratified: preserving the proportion of HC and SZ examples (**Leaky pipelines**) or subjects (**Subj pipelines**) in train/test sets |
| 6.5. Repetitions | CV schemes executed only once |
| 6.6. For multi-dataset: cross-dataset/pooled-dataset validation? | N/A |
| 6.7. Leakage confirmation | Contrasted subject-based and epoch-based (leaky) evaluation schemes |
| 7. **Feature Selection (if applicable, otherwise skip to 8.)** | |
| 7.1. Method | N/A |
| 7.2. Parameters | N/A |
| 7.3. Timing | N/A |
| 7.4. Leakage confirmation | N/A |
| 8. **Evaluation** | |
| 8.1. Metrics |  |
| 8.1.1. Primary | Accuracy (averaged over CV runs) |
| 8.1.2. Secondary | Balanced accuracy, sensitivity, specificity, precision, F1-score, MMC, Cohen’s Kappa, ROC-AUC, computed from aggregated CV outcomes |
| 8.2. Aggregation (if applicable) | N/A |
| 8.3. Statistical testing |  |
| 8.3.1. Chance-level | Not assessed |
| 8.3.2. Comparative | Leaky- vs. leakage-free DP pipelines contrasted via permutation testing (N=10000) |
| 8.4. Generalization (external data) | Not assessed |
| 8.5. Leakage confirmation | Epoch-based pipeline accuracy higher than subject-based pipeline accuracy |

### Pipeline 2: Boosted ensemble of decision trees with hand crafted features

| **Evaluation checklist** | |
| --- | --- |
| 1. **Model Specifics** | |
| 1.1. Model IDs | MSU-Ensemble-Leaky-Full, MSU-Ensemble-ablFS, MSU-Ensemble-ablDP, MSU-Ensemble-Subj  RepOD-Ensemble-Leaky-Full, RepOD-Ensemble-ablFS, RepOD-Ensemble-ablDP, RepOD-Ensemble-Subj  SU-SZ-Ensemble-Leaky-Full, SU-SZ-Ensemble-ablFS, SU-SZ-Ensemble-ablDP, SU-SZ-Ensemble-Subj |
| 1.2. Date of execution | MSU: 11/21/2025  RepOD: 11/21/2025  SU-SZ: 11/23/2025 |
| 1.3. Hardware environment | 12^th^ Gen Intel® CoreTM i9-12900Kx24  NVIDIA GeForce RTX 4090  Physical memory: 64 Gb |
| 1.4. Software environment | MATLAB R2025a |
| 1.5. Execution time | <5min for all |
| 1.6. Dataset IDs | MSU  RepOD  SU-SZ |
| 1.7. Study groups/response variable | MSU: HC vs. SZ  RepOD: HC vs. SZ  SU-SZ: HC vs. SZ |
| 1.8. Number of subjects (per group/condition) | MSU: HC=39, SZ=45  RepOD: HC=14, SZ=14  SU-SZ: HC=39, SZ=38 |
| 2. **Data Pre-processing** | |
| 2.1. Steps in order |  |
| 2.1.1. Channel selection – **only for SU-SZ** | Out of 64 channels, only the 19 corresponding to standard 10-20 are retained |
| 2.1.2. Band-pass filtering | 1-45 Hz |
| 2.1.3. Notch filtering | 50 Hz (CleanLine) |
| 2.1.4. Downsampling – **only for SU-SZ** | 1000 Hz data downsampled to 250 Hz |
| 2.1.5. ICA-based artifact removal (automated) | Multiple Artifact Rejection Algorithm, default parameters |
| 2.1.6. Re-referencing | Common average reference |
| 2.2. Software/tools | Matlab 2025a, EEGLAB |
| 2.3. Handling of missing/imbalanced data |  |
| 2.3.1. Method | N/A |
| 2.3.2. Before/after partitioning? | N/A |
| 2.4. Data augmentation | **Yes** / no / not applicable |
| 2.4.1. Method | Segmentation into 10-second non-overlapping epochs |
| 2.4.2. Justification | Increase sample size |
| 2.5. Output format (per subject) | MSU: 1280x16x6  RepOD: 2500x19x72  SU-SZ: 2500x19x10  (*time* *points* x *channels* x *epochs*) |
| 3. **Feature Extraction/Engineering** | |
| 3.1. Feature details |  |
| 3.1.1. Mean spectral power in five frequency bands (δ: 2-4 Hz, θ: 4-7 Hz, α: 7-12 Hz, β: 12-30 Hz, γ: 30-45 Hz) – 5 features/channel/epoch | Spectrogram obtained via spectrogram(), transformed to dB, then averaged over 5 frequency bands and averaged over time |
| 3.1.2. Variance of spectral power in five frequency bands (δ: 2-4 Hz, θ: 4-7 Hz, α: 7-12 Hz, β: 12-30 Hz, γ: 30-45 Hz) – 5 features/channel/epoch | Spectrogram obtained via spectrogram(), transformed to dB, then averaged over 5 frequency bands and standard deviation computed over time |
| 3.1.3. Hjorth parameters from broadband (1-45 Hz) EEG – 3 features/channel/epoch | Activity, Motility and Complexity. Activity is computed before EEG data standardization. |
| 3.1.4. Detrended Fluctuation Analysis scaling exponent from broadband (1-45 Hz) EEG – 1 feature/channel/epoch | Scaling function is obtained at scales 4, 8, 16, 32, 64 and 128 data points (MSU) or 8, 16, 32, 64, 128, 256 data points (RepOD, SU-SZ) |
| 3.1.5. Permutation Entropy in five frequency bands (δ: 2-4 Hz, θ: 4-7 Hz, α: 7-12 Hz, β: 12-30 Hz, γ: 30-45 Hz) – 5 features/channel/epoch | Embedding dimension and lag set to match frequency range and sampling rate |
| 3.1.6. Weighted node degree in five frequency bands (δ: 2-4 Hz, θ: 4-7 Hz, α: 7-12 Hz, β: 12-30 Hz, γ: 30-45 Hz) – 5 features/channel/epoch | Connectivity matrices are reconstructed using Phase-Lag Index, then density thresholded between 15-50% and averaged |
| 3.2. Dimensionality reduction | Yes / **no** / not applicable |
| 3.2.1. Method | N/A |
| 3.2.2. Parameters | N/A |
| 3.2.3. Timing (i.e., before/after data partitioning) | N/A |
| 3.3. Normalization |  |
| 3.3.1. Method | N/A |
| 3.3.2. Parameters | N/A |
| 3.3.3. Timing | N/A |
| 3.4. Aggregation (if applicable) | none |
| 3.5. Output format (per subject) | MSU: 6x384  RepOD: 72x456  SU-SZ: 10x456 |
| 4. **Model Architecture and Training** | |
| 4.1. Model type | Boosted ensemble of decision trees |
| 4.2. Implementation | Matlab templateTree()  MaxNumSplits=5  Surrogate=’on’ |
| 4.3. Input format | MSU: 1x386  RepOD: 1x456  SU-SZ: 1x456 |
| 4.4. Key parameters | method=’GentleBoost’  numlearningcycles=100  learnrate=0.1 |
| 4.5. Training procedure | Matlab fitensemble() |
| 4.6. Normalization + timing (before/after data partitioning) | Standardization (z-scoring)  **ablFS** and **Subj pipelines**: inside-the-loop, standardizing test set using mean and variance obtained from training set  **Leaky-Full** and **ablDP pipelines**: outside-the-loop using full dataset |
| 5. **Hyperparameter Tuning (if applicable, otherwise skip to 6.)** | |
| 5.1. Method | N/A |
| 5.2. Parameters tuned | N/A |
| 5.3. Timing/scheme | N/A |
| 5.4. Leakage confirmation | N/A |
| 6. **Data Partitioning (DP) and Cross-Validation (CV, if applicable)** | |
| 6.1. Full dataset size | MSU: 504x384  RepOD: 2016x456  SU-SZ: 770x456 |
| 6.2. Scheme and ratio | **Leaky-Full** and **ablFS pipelines**: stratified 10-fold CV (7-fold for RepOD)  **ablDP** and **Subj pipelines:** subject-based stratified 10-fold CV (7-fold for RepOD) |
| 6.3. Hierarchical handling (i.e., group-based CV) | Subject-based data partitioning in **ablDP** and **Subj** **pipelines** |
| 6.4. Stratification | Stratified: preserving the proportion of HC and SZ examples (**Leaky-full** and **ablFS pipelines**) or subjects (**ablDP** and **Subj pipelines**) in train/test sets |
| 6.5. Repetitions | CV schemes executed only once |
| 6.6. For multi-dataset: cross-dataset/pooled-dataset validation? | N/A |
| 6.7. Leakage confirmation | Subject-based and epoch-based DP pipelines contrasted via permutation testing |
| 7. **Feature Selection (if applicable, otherwise skip to 8.)** | |
| 7.1. Method | ReliefF |
| 7.2. Parameters | k=10  Either top 50% of features or all features with positive weights retained, whichever is smaller |
| 7.3. Timing | **Leaky-Full** and **ablDP pipelines**: Outside-the-loop  **ablFS** and **Subj pipelines**: Inside-the-loop |
| 7.4. Leakage confirmation | Inside-the-loop and outside-the-loop FS pipelines contrasted via permutation testing |
| 8. **Evaluation** | |
| 8.1. Metrics |  |
| 8.1.1. Primary | Accuracy (averaged over CV runs) |
| 8.1.2. Secondary | Balanced accuracy, sensitivity, specificity, precision, F1-score, MMC, Cohen’s Kappa, ROC-AUC, computed from aggregated CV outcomes |
| 8.2. Aggregation (if applicable) | N/A |
| 8.3. Statistical testing |  |
| 8.3.1. Chance-level | Not assessed |
| 8.3.2. Comparative | Leaky-Full, ablFS, ablDP, Subj pipelines contrasted via permutation testing (N=10000) |
| 8.4. Generalization (external data) | Not assessed |
| 8.5. Leakage confirmation | Leaky DP accuracy significantly higher than leakage-free DP accuracy  No leakage effect from FS on accuracy |

### Pipeline 3: Subject-level Support Vector Machine with hand crafted features

| **Evaluation checklist** | |
| --- | --- |
| 1. **Model Specifics** | |
| 1.1. Model IDs | MSU-CNN-Leaky, MSU-CNN-Subj  RepOD-CNN-Leaky, RepOD-CNN-Subj  SU-SZ-CNN-Leaky, RepOD-CNN-Subj |
| 1.2. Date of execution | MSU: 11/21/2025  RepOD: 11/21/2025  SU-SZ: 11/21/2025 |
| 1.3. Hardware environment | 12^th^ Gen Intel® CoreTM i9-12900Kx24  NVIDIA GeForce RTX 4090  Physical memory: 64 Gb |
| 1.4. Software environment | MATLAB R2025a |
| 1.5. Execution time | <1min for all |
| 1.6. Dataset IDs | MSU  RepOD  SU-SZ |
| 1.7. Study groups/response variable | MSU: HC vs. SZ  RepOD: HC vs. SZ  SU-SZ: HC vs. SZ |
| 1.8. Number of subjects (per group/condition) | MSU: HC=39, SZ=45  RepOD: HC=14, SZ=14  SU-SZ: HC=39, SZ=38 |
| 2. **Data Pre-processing** | |
| 2.1. Steps in order |  |
| 2.1.1. Channel selection – **only for SU-SZ** | Out of 64 channels, only the 19 corresponding to standard 10-20 are retained |
| 2.1.2. Band-pass filtering | 1-45 Hz |
| 2.1.3. Notch filtering | 50 Hz (CleanLine) |
| 2.1.4. Downsampling – **only for SU-SZ** | 1000 Hz data downsampled to 250 Hz |
| 2.1.5. ICA-based artifact removal (automated) | Multiple Artifact Rejection Algorithm, default parameters |
| 2.1.6. Re-referencing | Common average reference |
| 2.2. Software/tools | Matlab 2025a, EEGLAB |
| 2.3. Handling of missing/imbalanced data |  |
| 2.3.1. Method | N/A |
| 2.3.2. Before/after partitioning? | N/A |
| 2.4. Data augmentation | **Yes** / no / not applicable |
| 2.4.1. Method | Segmentation into 10-second non-overlapping epochs |
| 2.4.2. Justification | Increase sample size |
| 2.5. Output format (per subject) | MSU: 1280x16x6  RepOD: 2500x19x72  SU-SZ: 2500x19x10  (*time* *points* x *channels* x *epochs*) |
| 3. **Feature Extraction/Engineering** | |
| 3.1. Feature details |  |
| 3.1.1. Mean spectral power in five frequency bands (δ: 2-4 Hz, θ: 4-7 Hz, α: 7-12 Hz, β: 12-30 Hz, γ: 30-45 Hz) – 5 features/channel/epoch | Spectrogram obtained via spectrogram(), transformed to dB, then averaged over 5 frequency bands and averaged over time |
| 3.1.2. Variance of spectral power in five frequency bands (δ: 2-4 Hz, θ: 4-7 Hz, α: 7-12 Hz, β: 12-30 Hz, γ: 30-45 Hz) – 5 features/channel/epoch | Spectrogram obtained via spectrogram(), transformed to dB, then averaged over 5 frequency bands and standard deviation computed over time |
| 3.1.3. Hjorth parameters from broadband (1-45 Hz) EEG – 3 features/channel/epoch | Activity, Motility and Complexity. Activity is computed before EEG data standardization. |
| 3.1.4. Detrended Fluctuation Analysis scaling exponent from broadband (1-45 Hz) EEG – 1 feature/channel/epoch | Scaling function is obtained at scales 4, 8, 16, 32, 64 and 128 data points (MSU) or 8, 16, 32, 64, 128, 256 data points (RepOD, SU-SZ) |
| 3.1.5. Permutation Entropy in five frequency bands (δ: 2-4 Hz, θ: 4-7 Hz, α: 7-12 Hz, β: 12-30 Hz, γ: 30-45 Hz) – 5 features/channel/epoch | Embedding dimension and lag set to match frequency range and sampling rate |
| 3.1.6. Weighted node degree in five frequency bands (δ: 2-4 Hz, θ: 4-7 Hz, α: 7-12 Hz, β: 12-30 Hz, γ: 30-45 Hz) – 5 features/channel/epoch | Connectivity matrices are reconstructed using Phase-Lag Index, then density thresholded between 15-50% and averaged |
| 3.2. Dimensionality reduction | Yes / **no** / not applicable |
| 3.2.1. Method | N/A |
| 3.2.2. Parameters | N/A |
| 3.2.3. Timing (i.e., before/after data partitioning) | N/A |
| 3.3. Normalization |  |
| 3.3.1. Method | N/A |
| 3.3.2. Parameters | N/A |
| 3.3.3. Timing | N/A |
| 3.4. Aggregation (if applicable) | Subject-level averaging over epochs: values for each feature are averaged over all epochs, yielding a single feature vector per subject |
| 3.5. Output format (per subject) | MSU: 1x384  RepOD: 1x456  SU-SZ: 1x456 |
| 4. **Model Architecture and Training** | |
| 4.1. Model type | Support vector machine |
| 4.2. Implementation | Matlab csvm |
| 4.3. Input format | MSU: 1x386  RepOD: 1x456  SU-SZ: 1x456 |
| 4.4. Key parameters | ‘Standardize’=false  ‘KernelFunction’=’RBF’  ‘KernelScale’=’auto’ |
| 4.5. Training procedure | Matlab fitcsvm() |
| 4.6. Normalization + timing (before/after data partitioning) | Standardization (z-scoring)  **ablFS** and **Subj pipelines**: inside-the-loop, standardizing test set using mean and variance obtained from training set  **Leaky-Full** and **ablDP pipelines**: outside-the-loop using full dataset |
| 5. **Hyperparameter Tuning (if applicable, otherwise skip to 6.)** | |
| 5.1. Method | N/A |
| 5.2. Parameters tuned | N/A |
| 5.3. Timing/scheme | N/A |
| 5.4. Leakage confirmation | N/A |
| 6. **Data Partitioning (DP) and Cross-Validation (CV, if applicable)** | |
| 6.1. Full dataset size | MSU: 88x384  RepOD: 28x456  SU-SZ: 77x456 |
| 6.2. Scheme and ratio | Extensive leave-one-subject-out CV: all possible HC-SZ subject pairs are evaluated as test set, with all remaining data as training set.  MSU: 1755 CV iterations  RepOD: 196 CV iterations  SU-SZ: 1482 CV iterations |
| 6.3. Hierarchical handling (i.e., group-based CV) | N/A |
| 6.4. Stratification | Singe HC-SZ pairs as test set in all CV iterations |
| 6.5. Repetitions | CV schemes executed only once |
| 6.6. For multi-dataset: cross-dataset/pooled-dataset validation? | N/A |
| 6.7. Leakage confirmation | N/A |
| 7. **Feature Selection (if applicable, otherwise skip to 8.)** | |
| 7.1. Method | Matlab relieff()  Matlab fscmrmr() |
| 7.2. Parameters | ReliefF: k=10  All features with positive weights retained  MRMR performed on reduced feature set, only 10 top ranking features retained |
| 7.3. Timing | **Leaky-Full** and **ablDP pipelines**: Outside-the-loop (ReliefF on full dataset)  **ablFS** and **Subj pipelines**: Inside-the-loop (ReliefF on training set only) |
| 7.4. Leakage confirmation | Inside-the-loop and outside-the-loop FS pipelines contrasted via permutation testing |
| 8. **Evaluation** | |
| 8.1. Metrics |  |
| 8.1.1. Primary | Accuracy (averaged over CV runs) |
| 8.1.2. Secondary | Balanced accuracy, sensitivity, specificity, precision, F1-score, MMC, Cohen’s Kappa, ROC-AUC, computed from aggregated CV outcomes |
| 8.2. Aggregation (if applicable) | N/A |
| 8.3. Statistical testing |  |
| 8.3.1. Chance-level | Not assessed |
| 8.3.2. Comparative | Leaky- vs. leakage-free DP pipelines contrasted via permutation testing (N=10000) |
| 8.4. Generalization (external data) | Not assessed |
| 8.5. Leakage confirmation | Leaky FS accuracy significantly higher than leakage-free FS accuracy |

### Pipeline 4: EEG Foundation Model (REVE) feature vectors with linear Support Vector Machine classifier

| **Evaluation checklist** | |
| --- | --- |
| 1. **Model Specifics** | |
| 1.1. Model IDs | MSU-FM-Leaky, MSU-FM-Subj  RepOD-FM-Leaky, RepOD-FM-Subj  SU-SZ-FM-Leaky, RepOD-FM-Subj |
| 1.2. Date of execution | MSU: 04/29/2026  RepOD: 04/29/2026  SU-SZ: 04/29/2026 |
| 1.3. Hardware environment | 12^th^ Gen Intel® CoreTM i9-12900Kx24  NVIDIA GeForce RTX 4090  Physical memory: 64 Gb |
| 1.4. Software environment | MATLAB R2025a (data preparation, SVM classification)  Python 3.11.15 (for REVE feature extraction)  Packages: scipy, numpy, torch, transformers, os, tqdm, glob, mat73 |
| 1.5. Execution time | MSU: ~3min  RepOD: ~6min  SU-SZ: ~3min |
| 1.6. Dataset IDs | MSU  RepOD  SU-SZ |
| 1.7. Study groups/response variable | MSU: HC vs. SZ  RepOD: HC vs. SZ  SU-SZ: HC vs. SZ |
| 1.8. Number of subjects (per group/condition) | MSU: HC=39, SZ=45  RepOD: HC=14, SZ=14  SU-SZ: HC=39, SZ=38 |
| 2. **Data Pre-processing** | |
| 2.1. Steps in order – please extend |  |
| 2.1.1. Channel selection – **only for SU-SZ** | Out of 64 channels, only the 19 corresponding to standard 10-20 are retained |
| 2.1.2. Band-pass filtering | 1-45 Hz |
| 2.1.3. Notch filtering | 50 Hz (CleanLine) |
| 2.1.4. Downsampling – **only for SU-SZ** | 1000 Hz data downsampled to 250 Hz |
| 2.1.5. ICA-based artifact removal (automated) | Multiple Artifact Rejection Algorithm, default parameters |
| 2.1.6. Re-referencing | Common average reference |
| 2.1.7. Normalization | Standardization (z-scoring) per channel |
| 2.1.8. Resampling | MSU: upsampling to 200 Hz  RepOD, SU-SZ: downsampling to 200 Hz |
| 2.1.9. Data conversion | Conversion to float32 |
| 2.1.10. Normalization | Standardization (z-scoring) per channel |
| 2.2. Software/tools | Matlab 2025a, EEGLAB |
| 2.3. Handling of missing/imbalanced data |  |
| 2.3.1. Method | N/A |
| 2.3.2. Before/after partitioning? | N/A |
| 2.4. Data augmentation | **Yes** / no / not applicable |
| 2.4.1. Method | Segmentation into 5-second non-overlapping epochs |
| 2.4.2. Justification | Increase sample size |
| 2.5. Output format (per subject) | MSU: 12x16x1000  RepOD: 144x19x1000  SU-SZ: 20x19x1000  (*epochs* x *channels* x *time points*) |
| 3. **Feature Extraction/Engineering** | |
| 3.1. Feature details – please extend (feature type, unit of FE, number of features per unit, etc.) |  |
| 3.1.1. Removing epochs with outliers | EEG epochs with any value outside the ±15 SD range are removed. Average number of epohcs:  MSU: *n=*12 ± 0  RepOD: *n=*139.61 ± 3.29  SU-SZ: *n=*19.48 ± 0.75 |
| 3.1.2. Obtaining feature vectors via REVE base model | model = AutoModel.from_pretrained ("brain-bzh/reve-base", trust_remote_code=True)  model.eval() |
| 3.1.3. Data aggregation | Original embeddings of size 5x512 were mean-aggregated to produce single vectors of size 1x512 per EEG epoch |
| 3.2. Dimensionality reduction | Yes / **no** / not applicable |
| 3.2.1. Method | N/A |
| 3.2.2. Parameters | N/A |
| 3.2.3. Timing (i.e., before/after data partitioning) | N/A |
| 3.3. Normalization |  |
| 3.3.1. Method | N/A |
| 3.3.2. Parameters | N/A |
| 3.3.3. Timing | N/A |
| 3.4. Aggregation (if applicable) | none |
| 3.5. Output format (per subject) | MSU: n_epochs x 512  RepOD: n_epochs x 512  SU-SZ: n_epochs x 512 |
| 4. **Model Architecture and Training** | |
| 4.1. Model type | Support vector machine |
| 4.2. Implementation | Matlab csvm |
| 4.3. Input format | MSU: 1x512  RepOD: 1x512  SU-SZ: 1x512 |
| 4.4. Key parameters | ‘Standardize’=true  ‘KernelFunction’=linear |
| 4.5. Training procedure | Matlab fitcsvm() |
| 4.6. Normalization + timing (before/after data partitioning) | Internally with fitcsvm() |
| 5. **Hyperparameter Tuning (if applicable, otherwise skip to 6.)** | |
| 5.1. Method | N/A |
| 5.2. Parameters tuned | N/A |
| 5.3. Timing/scheme | N/A |
| 5.4. Leakage confirmation | N/A |
| 6. **Data Partitioning (DP) and Cross-Validation (CV, if applicable)** | |
| 6.1. Full dataset size | MSU: 1008x512  RepOD: 3909x512  SU-SZ: 1500x512 |
| 6.2. Scheme and ratio | **Leaky pipelines**: stratified 10-fold CV  (7-fold CV for RepOD)  **Subj pipelines:** subject-based stratified 10-fold CV (7-fold CV for RepOD) |
| 6.3. Hierarchical handling (i.e., group-based CV) | Subject-based data partitioning in **Subj** **pipelines** |
| 6.4. Stratification | Stratified: preserving the proportion of HC and SZ examples (**Leaky pipelines**) or subjects (**Subj pipelines**) in train/test sets |
| 6.5. Repetitions | CV schemes executed only once |
| 6.6. For multi-dataset: cross-dataset/pooled-dataset validation? | N/A |
| 6.7. Leakage confirmation | Contrasted subject-based and epoch-based (leaky) evaluation schemes |
| 7. **Feature Selection (if applicable, otherwise skip to 8.)** | |
| 7.1. Method | N/A |
| 7.2. Parameters | N/A |
| 7.3. Timing | N/A |
| 7.4. Leakage confirmation | N/A |
| 8. **Evaluation** | |
| 8.1. Metrics |  |
| 8.1.1. Primary | Accuracy (averaged over CV runs) |
| 8.1.2. Secondary | Balanced accuracy, sensitivity, specificity, precision, F1-score, MMC, Cohen’s Kappa, ROC-AUC, computed from aggregated CV outcomes |
| 8.2. Aggregation (if applicable) | N/A |
| 8.3. Statistical testing |  |
| 8.3.1. Chance-level | Not assessed |
| 8.3.2. Comparative | Leaky- vs. leakage-free DP pipelines contrasted via permutation testing (N=10000) |
| 8.4. Generalization (external data) | Not assessed |
| 8.5. Leakage confirmation | Epoch-based pipeline accuracy higher than subject-based pipeline accuracy |
